# COVID-19, economic downturn, and long-term trajectories of population mental health: evidence from two nationally representative British birth cohorts at the intersection of gender and socioeconomic position

**DOI:** 10.1101/2025.09.02.25334824

**Authors:** Darío Moreno-Agostino, George B. Ploubidis, Jayati Das-Munshi

## Abstract

**Background:** We examined long-term trajectories of mental (ill-)health in two British generations (‘Baby boomers’ and ‘Generation X’) across the life-course, including the COVID-19 lockdowns and the following cost-of-living increases. We analysed inequalities by generation, gender, socioeconomic position (SEP), and their intersections, and explored the relationship between inflation and mental (ill-)health post-lockdown.

**Methods and Findings:** We used data from the National Child Development Study (NCDS/1958, n=8,215) and the 1970 British Cohort Study (BCS/1970, n=7,789), with repeated measures of psychological distress (Malaise Inventory) between ages 23-64.5 (NCDS/58) and 26-52.5 (BCS/70). We used multilevel growth curve models to study long-term trajectories, and negative binomial regression models to analyse associations with inflation/cost-of-living in the 2021-2023 period. Distress increased during the pandemic but declined post-lockdown (B_spline2_quadratic_NCDS/58_=−0.12 [−0.17, −0.08], *p*<0.001; B_spline2_quadratic_BCS/70_=−0.16 [−0.21, −0.11], *p*<0.001). Women and those from disadvantaged childhood SEPs started their trajectories at significantly (*p*<0.001) higher levels in both NCDS/58 (B_women_=0.72 [0.62, 0.82]; B_manual_=0.24 [0.14, 0.35]; B_rented7&11_=0.34 [0.22, 0.46]) and BCS/70 (B_women_=0.73 [0.62, 0.83]; B_manual_=0.23 [0.12, 0.35]; B_rented5&10_=0.30 [0.15, 0.45]), with even larger inequalities for women from disadvantaged childhood SEPs born in 1958 (intersectional effects). None of these inequalities significantly reduced in the long term. Inflation/cost-of-living was significantly associated with distress, but effects did not vary by gender, concurrent SEP, or their intersection.

**Conclusions:** Despite post-pandemic improvements, persistent inequalities by gender and childhood SEP remain. Considering the existing high levels of socioeconomic adversity in the UK, efforts must be made to reverse these gaps and prevent further inequalities later in life and intergenerational transmission.

## Introduction

The global COVID-19 pandemic came with a deterioration in population mental health, with disproportionate impacts among disadvantaged groups (1, 2). In the UK context, this global shock disrupted the long-term trajectories of mental health, widening pre-existing inequalities such as those by gender (3–5). In many countries, this period was followed by a rapid increase in the cost of living, with global consumer prices reaching levels unseen since the 2008 financial crisis (6). In the UK, similar inflation levels had not been seen since 1990, with the Consumer Prices Index including owner occupiers’ housing costs (CPIH) reaching a peak of 9.6% in October 2022 (7). However, there is no population-based, longitudinal evidence on how long-term trajectories of mental (ill-)health have developed after the COVID-19 lockdowns, a period largely coinciding with large cost-of-living increases, and whether some of the inequalities that widened during the lockdown period have changed.

Economic shocks can impact population mental health, particularly among those socioeconomically disadvantaged (8–10). From a long-term perspective, socioeconomic adversity plays a fundamental and complex role on mental (ill-)health across the life course and even across life courses (e.g., through intergenerational transmission) (11, 12). Even early life socioeconomic disadvantage, over which people have little to no agency, can impact mental health later in life and lead to further socioeconomic adversity (13). Despite being one of the wealthiest economies globally, levels of poverty have remained consistently high in the UK (14). This has led some scholars to propose that ‘poverty pandemic’ may be a more accurate term than the widely used ‘cost-of-living crisis’ (15), as the situation seems “endemic, not a short-term crisis to weather” (16). Studying the trajectories of mental (ill-)health in the population from a life-course perspective can help to understand the potential role of the cost-of-living increases both in the context of other recent (e.g., COVID-19 pandemic) and longer-term (e.g., early life socioeconomic disadvantage) phenomena.

When studying how population mental health levels may have changed after the lockdown period, there are additional sources of complexity to consider. First, understanding whether and how the large (and, during the pandemic, widening (4, 5)) gender inequalities in mental (ill-)health have changed is key to grasp the extent of the action needed to close them. Importantly, the systems of oppression that underlie the gender and socioeconomic gaps in mental health (including sexism, the material impacts of socioeconomic deprivation, and classism) are complex and interlocked, rather than independent from each other. This idea is central to intersectionality theories (17, 18). Importantly, inequalities at specific intersections will remain undetected unless explicitly acknowledged when quantitatively analysing social inequalities (19). Second, there is compelling evidence of a deterioration in multiple health outcomes across generations in the UK, or a ‘generational health drift’ (20, 21).

This includes mental (ill-)health outcomes, with younger generations (particularly those born in 1970s and onwards) experiencing worse mental health levels than older generations at similar or same ages (4, 22–24). People from different generations have lived through different socio-historical contexts in which power and privilege are differently distributed (25). For example, the percentage of women aged 25-54 in paid employment (including self-employment) went from 57% in 1975 to 78% in 2017 (26). And, although it is far from being resolved, the percentual gender pay gap among all employees has slowly reduced over time, from 27.5% in 1997 to 14.4% in 2022 (27). The Equal Pay Act, a key milestone for the legal basis of gender pay equality, was introduced in 1970, with amendments and further legislation being introduced in the 80s, 90s, and most recently consolidated in the Equality Act 2010 (28). By taking place at different times in their lifespan, societal conditions and events like these can have different implications for, in this example, women and men of different generations.

Considering all the above, studying the long-term trajectories of mental (ill-)health from a life-course, cross-generational, and intersectional perspective has multiple advantages. First, it can inform whether the observed increasing trends in psychological distress observed during the COVID-19 pandemic have continued, stopped, or reversed as populations exited the lockdown periods. Second, it can inform how social inequalities observed during the COVID-19 pandemic (e.g., widening gender inequalities) have evolved in the post-lockdown era. Third, it can provide further insights into how the generational decline (or ‘generational health drift’ (20)) in mental health is evolving. And fourth, when coupling this cross-generational approach with a focus on early life determinants of long-term trajectories, it can serve as a measure of societal progress, by informing whether certain social inequalities have meaningfully reduced across generations and in the long term.

Therefore, the two primary aims of this study were #1) to understand how long-term trajectories of mental (ill-)health developed in post-lockdown Britain, a period largely coinciding with cost-of-living increases; and #2) to examine differences in those long-term trajectories by generation, gender, childhood socioeconomic position, and their intersections. As a secondary aim, we also aimed #3) to examine the relationship between inflation and population mental (ill-)health, and whether this relationship varied by gender, socioeconomic position, and their intersections.

## Methods

### Sample

We used data from two British birth cohorts: the 1958 National Child Development Study (NCDS/58) (29) and the 1970 British Cohort Study (BCS/70) (30). These are two nationally representative birth cohort studies following up the lives of people born in Britain in a single week in 1958 and 1970, respectively. Data on the cohort members have been collected since their birth and throughout their life courses. The most recent main survey data collections (*sweeps*) took place between February 2019-April 2024 in NCDS/58 and between January 2020-January 2024. As such, they started before the COVID-19 pandemic, were interrupted during the lockdown period, and resumed after the lockdowns. However, data collection continued during the pandemic as part of the COVID-19 Surveys (31), which collected data from members of these and other cohorts at three time-points during the COVID-19 pandemic: May 2020 (during first national lockdown), September-October 2020 (before second national lockdown), and February-March 2021 (during third national lockdown) (32). A total of 8,215 (NCDS/58) and 7,789 (BCS/70) people took part in the pilots, dress rehearsals, main stage, or mop-up web surveys in the most recent data collection, corresponding to response rates of 73.3% and 65.3%, respectively. The target population for the present study were adults born in Britain in 1958 or 1970, still alive and residing in the UK during the most recent main survey sweep. All procedures involving human subjects/patients were approved by the National Health Service (NHS) Research ethics Committee. All participants provided oral informed consent.

### Measures

#### Main outcome: psychological distress

Psychological distress (a measure of mental ill-health including depressive and anxiety symptomatology) was the main outcome in both the within-person long-term trajectory and between-person post-lockdown analyses. It was measured with the nine-item version of the Malaise Inventory (33). This questionnaire explores whether the respondent ‘often’ experiences a series of general mental ill-health experiences, with “yes/no” response options. Hence, the sum-score ranges between 0 (lowest psychological distress) and 9 (highest psychological distress) at any time-point. The questionnaire was administered at ages 23, 33, 42, 50, 62, 62.5, 63, and 64.5 in NCDS/58 and at ages 26, 29, 34, 42, 46, 50, 50.5, 51, and 52.5 in BCS/70. Previous studies have provided evidence on the appropriateness of a sum-score approach and on the invariance of the resulting measure across time-points, genders, and cohorts (4, 34, 35). Due to the aims of this study and the use of additional newly collected data, we extended this previous evidence by including the most recent data collection time-points and by analysing the measurement invariance at the intersection of gender and socioeconomic position, both within and across cohorts (an often overlooked aspects in quantitative research on intersectional inequalities (36)). Additional details on this approach are available in **eAppendix 1 (Supplementary Material)**.

#### Social identities and positions

We used an inter-categorical approach to intersectional complexity (37), provisionally adopting categorical variables as proxies of the systems of oppression that may lead to inequities in the outcomes under study.

We used sex assigned at birth as a proxy for gender, as information on gender identity was not consistently available from participants. We, however, refer to gender rather than sex inequalities throughout as our position is that differences between women and men in the outcome under study will be more likely due to differences in power and privilege than due to inherent biological characteristics.

We used two alternative childhood socioeconomic position indicators as proxies for social class and classism, harmonised across NCDS/58 and BCS/70: parental social class and childhood housing tenure. A six-category harmonised variable representing the cohort member’s father’s social class at ages 11 (NCDS/58) or 10 (BCS/70) was obtained from the UK Data Service (UKDS) based on the work by Dodgeon et al. (38). This was further dichotomised into manual (including the skilled manual, partly skilled, and unskilled categories) and non-manual (including the skilled non-manual, managerial and technical, and professional categories) for the purposes of this study. A three-category harmonised variable capturing whether the cohort member lived in an accommodation that was rented, owned at one time-point, or owned at both time-points at ages 7&11 (NCDS/58) or 5&10 (BCS/70) was obtained from the UKDS based on the work by Wood et al. (39).

#### Confounders

As ‘time’ was the main exposure in the long-term trajectory analyses, and it may be considered unconfounded in on itself (although related to multiple other processes that unfold with time which may explain changes in the outcome under study), no confounders were adjusted for in these analyses.

### Statistical analysis

#### Within-person long-term trajectory analyses

Multilevel growth curve models (40, 41) were used to understand the change in psychological distress over time, including the period with large cost-of-living increases (aim #1), and differences in these changes by generation, gender, childhood socioeconomic groups, and their intersections (aim #2).

Cohort members whose participation in the most recent main survey sweep took place before the COVID-19 pandemic (1,662 in NCDS/58 and 116 in BCS/70) or within the period spanning the COVID-19 Surveys data collection (44 participants from BCS/70 whose interviews took place between 11 September 2020 and 7 October 2020) were not included in these analyses. Age at the most recent sweep was set at the median weighted age among included cohort members, which was 64.5 in NCDS/58 and 52.5 in BCS/70 (roughly corresponding to the second half of 2022). Therefore, the resulting repeated measures of psychological distress spanned ages 23, 33, 42, 50, 62, 62.5, 63, and 64.5 in NCDS/58 and ages 26, 29, 34, 42, 46, 50, 50.5, 51, and 52.5 in BCS/70.

Models with different growth parameters and random effects (to accommodate individual variation in the included growth parameters) were tested and selected upon the best model fit according to Akaike’s Information Criteria (AIC) and Bayesian Information Criteria (BIC), with lower AIC and BIC indicating better fit. Candidate models were selected based upon the descriptive analysis and visualisation of imputed and weighted life-course data. The selected models were then estimated in each cohort, first overall, then by gender and by each of the childhood socioeconomic position indicators, and finally by the intersection of gender and each of the childhood socioeconomic position indicators, by including the appropriate interaction terms between the growth parameters and the group variables. To aid with the interpretation of the results, marginal predicted levels at each of the time-points (and for each of the groups in the analyses by gender, childhood socioeconomic position, and their intersection) were obtained and visualised.

As a check of the sensitivity of the results to model specification and sample selection, a set of additional overall models were estimated where continuous age was included instead of median age as the main time variable at the latest sweep. These analyses included all participants regardless of whether their most recent main survey sweep interview had taken place before/during the COVID-19 pandemic.

To more specifically characterise any potential change in the gender, socioeconomic, and intersectional gaps over time and across cohorts, an additional set of analyses were conducted comparing the psychological distress levels in the most recent sweep (roughly, as abovementioned, the second half of 2022) with three relevant previous time-points: 1) the earliest time-point in the long-term trajectory (age 23 in NCDS/58 or 26 in BCS/70; 1981 and 1996, respectively), 2) the most recent pre-pandemic assessment (age 50 in NCDS/58 and 46 in BCS/70; 2008 and 2016, respectively), and 3) the point of highest psychological distress during the COVID-19 pandemic (age 62.5 in NCDS/58 or 50.5 in BCS/70; September/October 2020). These analyses were conducted separately for each cohort and, then, pooling together the data from the two cohorts, adding cohort as a main and interaction variable with the existing terms to explore any cohort differences in the change across time-points, gaps, and their potential change over time.

#### Between-person post-lockdown analyses

To provide further insights on the potential shorter-term relationship between inflation and mental (ill-)health both in general and by pre-existing sources of disadvantage (aim #3), a separate group of analyses were conducted focusing on the post-lockdown period and leveraging the time variability in the data collection. Additional details on the rationale for these analyses, as well as the measurement and analytical approach used, are available in **eAppendix 2** (**Supplementary Material**).

#### Missing data

Inverse probability weighting (IPW) and multiple imputation by chained equations (MICE) were used to deal with missing data, both under the assumption that data were missing at random (MAR) after conditioning on observed variables (42).

Non-response weights were derived and used to help restore sample representativeness to the target population (i.e., people born in Britain 1958 or 1970, still alive and residing in the UK at the time of the most recent sweep), using relevant predictors of non-response from across the individuals’ life-courses (43). Full details on the derivation of the weights and their effectiveness to restore representativeness to the target population are available in the cohort studies’ user guides (44, 45).

Multiple Imputation by Chained Equations (MICE) was used to flexibly deal with item-missingness (46). To increase the plausibility of the MAR assumption, we used cohort members’ prior life-course data which has been found to be related to attrition and/or to the missing values themselves as auxiliary variables, in line with previous research (47) and with the Centre for Longitudinal Studies (CLS) missing data strategy (43). The complete list of auxiliary variables included in the imputation models is available in **eAppendix 3 (Supplementary Material)**. Fifty imputed datasets were created, discarding the first 10 iterations of each chain. The analytical models were then conducted over the imputed datasets using Rubin’s rules to pool the estimates and standard errors (42).

All data used in this study are freely available to *bona* fide researchers at the UK Data Service (https://doi.org/10.5255/UKDA-Series-200001 and https://doi.org/10.5255/UKDA-Series-2000032). All code used for the data management, imputation, and analysis is available in the Open Science Framework (https://osf.io/msprq/).

We followed the Strengthening the Reporting of Observational Studies in Epidemiology (STROBE) checklist **(eAppendix 5, Supplementary Material)**.

All analyses were conducted in Stata MP 19.5 (48), except measurement invariance testing, which was conducted in Mplus 8.9 (49).

## Results

The overall sample for this study included 6,553 participants from the NCDS/58 cohort (n=3,277, 50.0% women) and 7,629 participants from the BCS/70 cohort (n=4,015, 52.6% women), after excluding cohort members whose latest main survey sweep took place before the COVID-19 pandemic onset (n=1,662 in NCDS/58 and n=116 in BCS/70) or within the period spanning the COVID-19 Surveys data collection (n=44 in BCS/70, interviewed between September-October 2020). Included participants contributed a median and interquartile range of 7 (5, 8) repeated observations in both cohorts (M_obs,NCDS/58_=6.34, M_obs,BCS/70_=6.56). The percentage of participants in a disadvantaged childhood socioeconomic position was, overall, smaller in BCS/70 (21.9% rented at both time points, 47.4% manual parental social class) compared to NCDS/58 (36.9% and 51.8%, respectively). Similar percentages of missing data were found in the socioeconomic position indicators across both cohorts, although this was slightly higher for the outcome variables in BCS/70 than in NCDS/58. Further descriptive details of the analytical samples, along with the percentage of missing data for the key variables of interest, are available in **Table 1**, and an extended table including a comparison with the overall samples of each cohort study is available in **eAppendix 6** (**Supplementary Material**).

**Table 1.**
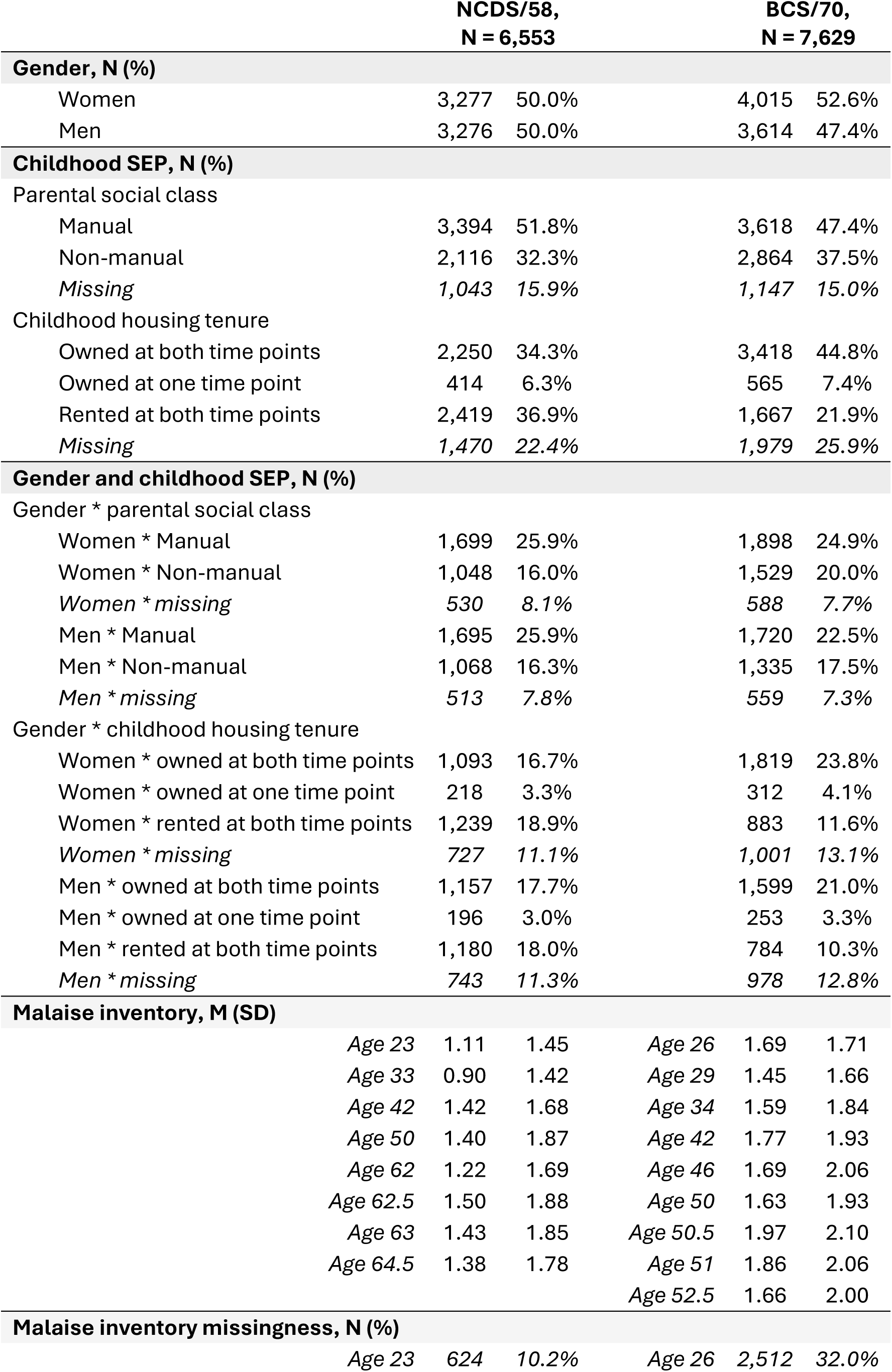
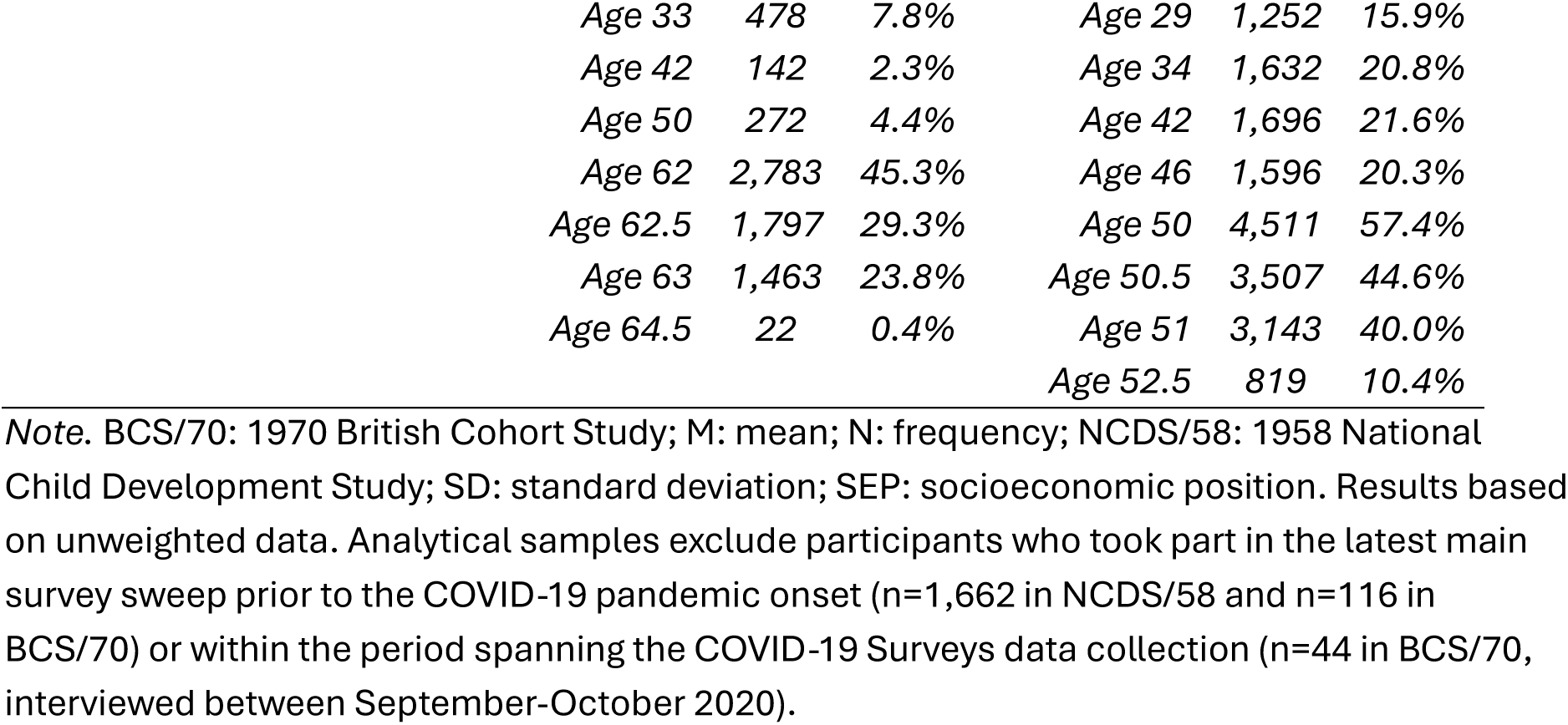
Descriptive information from the analytical samples.

|  | NCDS/58,<br>N = 6,553 |  |  | BCS/70,<br>N = 7,629 |  |  |
| --- | --- | --- | --- | --- | --- | --- |
| Gender, N (%) |  |  |  |  |  |  |
| Women | 3,277 | 50.0% |  | 4,015 | 52.6% |  |
| Men | 3,276 | 50.0% |  | 3,614 | 47.4% |  |
| Childhood SEP, N (%) |  |  |  |  |  |  |
| Parental social class |  |  |  |  |  |  |
| Manual | 3,394 | 51.8% |  | 3,618 | 47.4% |  |
| Non-manual | 2,116 | 32.3% |  | 2,864 | 37.5% |  |
| Missing | 1,043 | 15.9% |  | 1,147 | 15.0% |  |
| Childhood housing tenure |  |  |  |  |  |  |
| Owned at both time points | 2,250 | 34.3% |  | 3,418 | 44.8% |  |
| Owned at one time point | 414 | 6.3% |  | 565 | 7.4% |  |
| Rented at both time points | 2,419 | 36.9% |  | 1,667 | 21.9% |  |
| Missing | 1,470 | 22.4% |  | 1,979 | 25.9% |  |
| Gender and childhood SEP, N (%) |  |  |  |  |  |  |
| Gender * parental social class |  |  |  |  |  |  |
| Women * Manual | 1,699 | 25.9% |  | 1,898 | 24.9% |  |
| Women * Non-manual | 1,048 | 16.0% |  | 1,529 | 20.0% |  |
| Women * missing | 530 | 8.1% |  | 588 | 7.7% |  |
| Men * Manual | 1,695 | 25.9% |  | 1,720 | 22.5% |  |
| Men * Non-manual | 1,068 | 16.3% |  | 1,335 | 17.5% |  |
| Men * missing | 513 | 7.8% |  | 559 | 7.3% |  |
| Gender * childhood housing tenure |  |  |  |  |  |  |
| Women * owned at both time points | 1,093 | 16.7% |  | 1,819 | 23.8% |  |
| Women * owned at one time point | 218 | 3.3% |  | 312 | 4.1% |  |
| Women * rented at both time points | 1,239 | 18.9% |  | 883 | 11.6% |  |
| Women * missing | 727 | 11.1% |  | 1,001 | 13.1% |  |
| Men * owned at both time points | 1,157 | 17.7% |  | 1,599 | 21.0% |  |
| Men * owned at one time point | 196 | 3.0% |  | 253 | 3.3% |  |
| Men * rented at both time points | 1,180 | 18.0% |  | 784 | 10.3% |  |
| Men * missing | 743 | 11.3% |  | 978 | 12.8% |  |
| Malaise inventory, M (SD) |  |  |  |  |  |  |
|  | Age 23 | 1.11 | 1.45 | Age 26 | 1.69 | 1.71 |
|  | Age 33 | 0.90 | 1.42 | Age 29 | 1.45 | 1.66 |
|  | Age 42 | 1.42 | 1.68 | Age 34 | 1.59 | 1.84 |
|  | Age 50 | 1.40 | 1.87 | Age 42 | 1.77 | 1.93 |
|  | Age 62 | 1.22 | 1.69 | Age 46 | 1.69 | 2.06 |
|  | Age 62.5 | 1.50 | 1.88 | Age 50 | 1.63 | 1.93 |
|  | Age 63 | 1.43 | 1.85 | Age 50.5 | 1.97 | 2.10 |
|  | Age 64.5 | 1.38 | 1.78 | Age 51 | 1.86 | 2.06 |
|  |  |  |  | Age 52.5 | 1.66 | 2.00 |
| Malaise inventory missingness, N (%) |  |  |  |  |  |  |
|  | Age 23 | 624 | 10.2% | Age 26 | 2,512 | 32.0% |

|  |  |  |  |  |  |
| --- | --- | --- | --- | --- | --- |
| *Age 33* | *478* | *7.8%* | *Age 29* | *1,252* | *15.9%* |
| *Age 42* | *142* | *2.3%* | *Age 34* | *1,632* | *20.8%* |
| *Age 50* | *272* | *4.4%* | *Age 42* | *1,696* | *21.6%* |
| *Age 62* | *2,783* | *45.3%* | *Age 46* | *1,596* | *20.3%* |
| *Age 62.5* | *1,797* | *29.3%* | *Age 50* | *4,511* | *57.4%* |
| *Age 63* | *1,463* | *23.8%* | *Age 50.5* | *3,507* | *44.6%* |
| *Age 64.5* | *22* | *0.4%* | *Age 51* | *3,143* | *40.0%* |
|  |  |  | *Age 52.5* | *819* | *10.4%* |

*Note.* BCS/70: 1970 British Cohort Study; M: mean; N: frequency; NCDS/58: 1958 National Child Development Study; SD: standard deviation; SEP: socioeconomic position. Results based on unweighted data. Analytical samples exclude participants who took part in the latest main survey sweep prior to the COVID-19 pandemic onset (n=1,662 in NCDS/58 and n=116 in BCS/70) or within the period spanning the COVID-19 Surveys data collection (n=44 in BCS/70, interviewed between September-October 2020).

### Long-term trajectory analyses

Evidence supporting the measurement invariance of the nine-item Malaise Inventory was found at the level needed to ensure valid comparisons of the psychological distress levels across time-points, genders, childhood socioeconomic position groups, and their intersections, as well as across birth cohorts. Further details on the approach and its results are available in **eAppendix 1 (Supplementary Material)**.

The model comparison strategy supported the use of a piecewise model with a cubic spline and a quadratic spline, with a knot at the latest pre-pandemic assessment, as well as random intercepts and slopes for the linear terms within each of the splines. Further details on the model selection approach and its results are available in **eAppendix 7 (Supplementary Material)**.

The overall long-term trajectory models in both birth cohorts showed that, after an initial increase in distress during the pandemic (mid-2020 to mid-2021) (B_spline2_linear_NCDS/58_=0.33 [0.21, 0.44], *p*<0.001; B_spline2_linear_BCS/70_=0.37 [0.24, 0.50], *p*<0.001), levels decreased towards the post-lockdown period (towards the second half of 2022) (B_spline2_quadratic_NCDS/58_=−0.12 [−0.17, −0.08], *p*<0.001; B_spline2_quadratic_BCS/70_=−0.16 [−0.21, −0.11], *p*<0.001), suggesting a bounce back to pre-pandemic levels (**Figure 1**). Similar results were found in the models with interaction terms. We found large inequalities at the first time-point in the long-term trajectory (age 23 in NCDS/58, age 26 in BCS/70), with women having significantly higher levels of distress than men (B_women_NCDS/58_=0.72 [0.62, 0.82], *p*<0.001; B_women_BCS/70_=0.73 [0.62, 0.83], *p*<0.001) (**Figure 2**), and those with parents from a non-manual social class (B_non-manual_NCDS/58_=−0.24 [−0.35, −0.14], *p*<0.001; B_non-manual_BCS/70_=−0.23 [−0.35, −0.12], *p*<0.001) (**Figure 3**) or living in an owned property during childhood (B_owned7&11_NCDS/58_=−0.34 [−0.46, −0.22], *p*<0.001; B_owned5&10_BCS/70_=−0.30 [−0.45, −0.15], *p*<0.001) (**Figure 4**) having significantly lower levels of distress than those with parents from a manual social class or living in a rented property at ages 7&11 (NCDS/58) or 5&10 (BCS/70).

**Figure 1.**
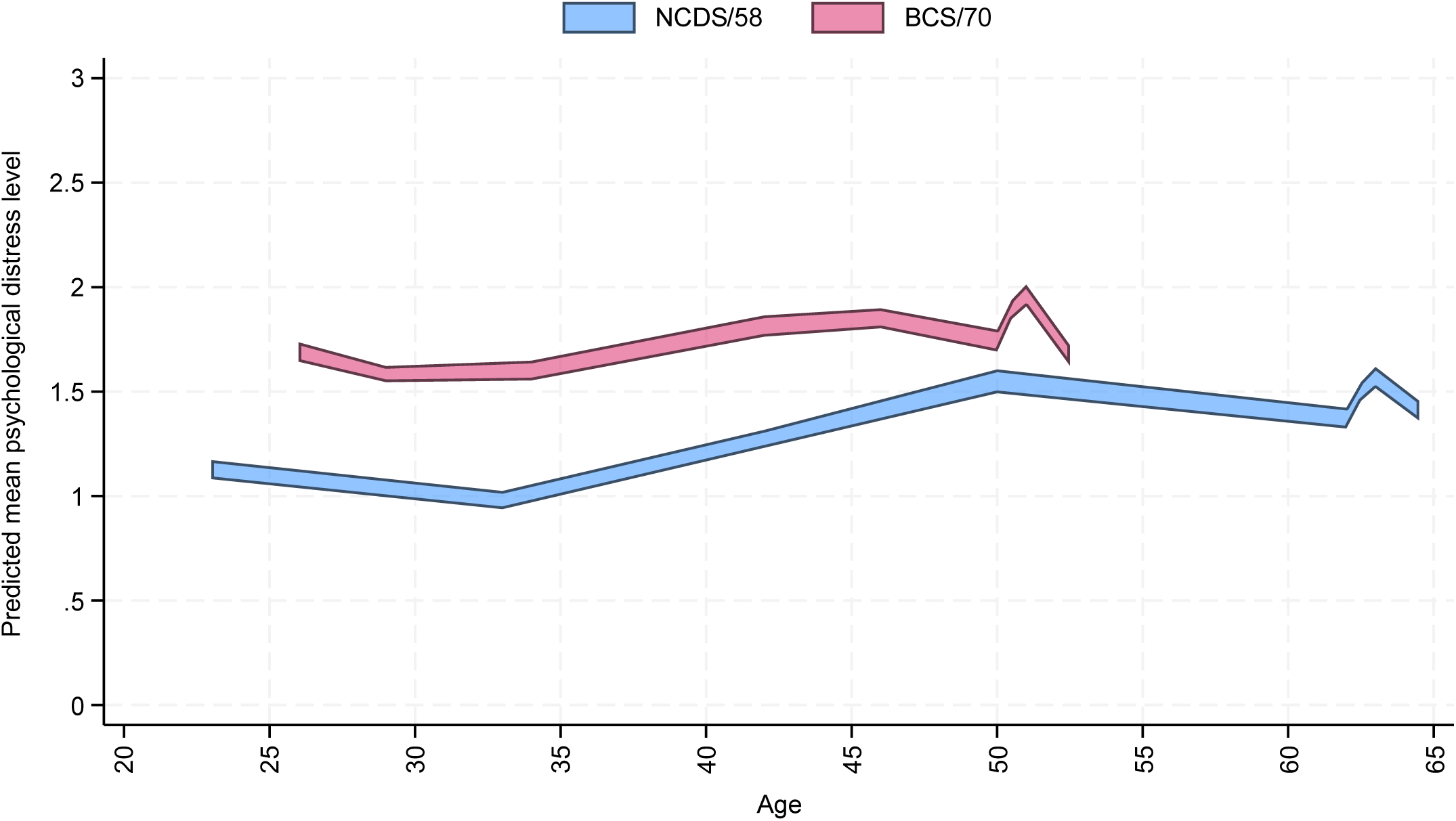
Overall long-term trajectories of psychological distress across NCDS/58 (n=6,553) and BCS/70 (n=7,629) *Note.* 95% confidence intervals for the marginal predicted mean psychological distress levels from the multilevel growth curve models. Results based on weighted and imputed data.

**Figure 2.**
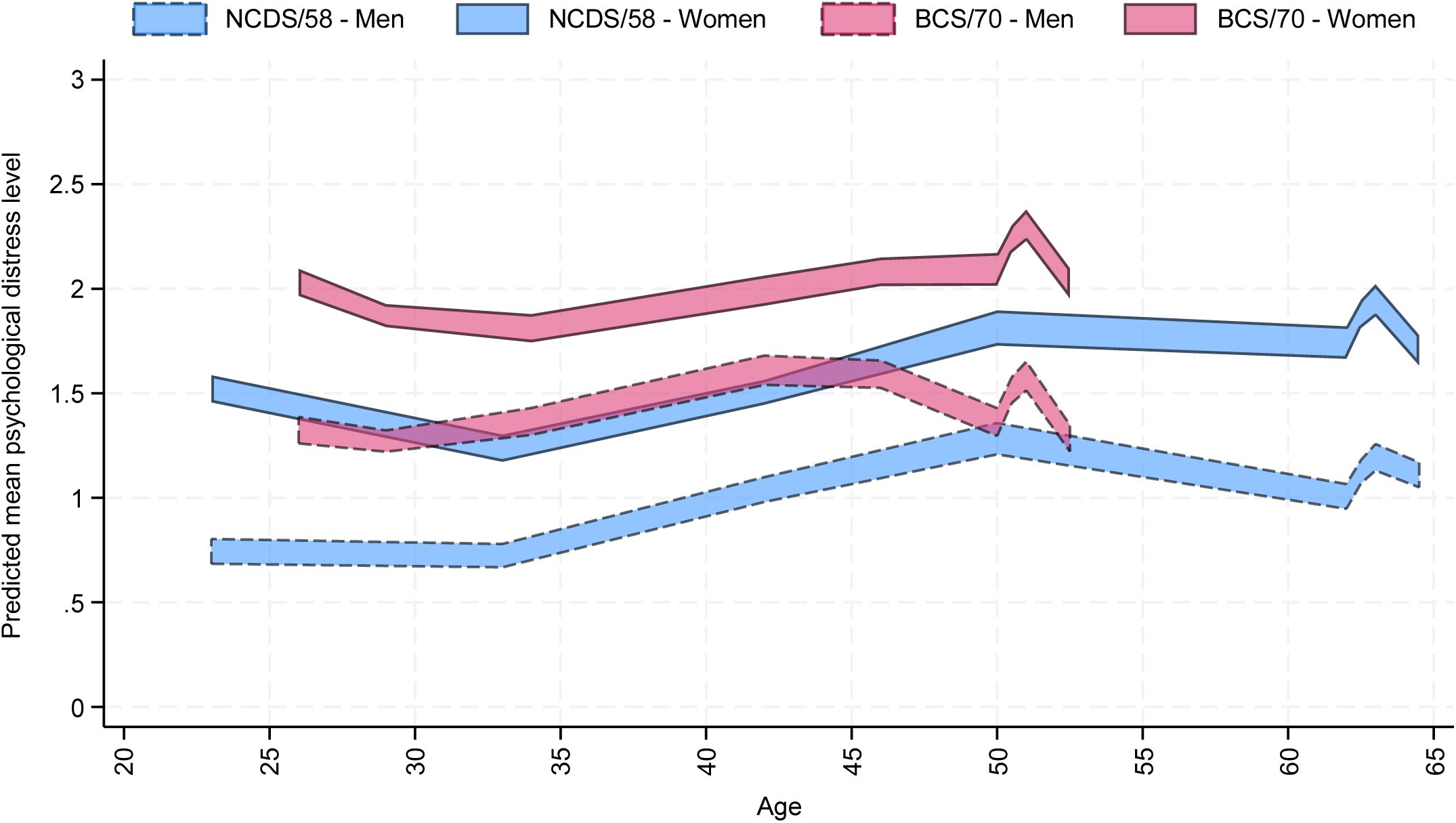
Long-term trajectories of psychological distress by gender across NCDS/58 (n=6,553) and BCS/70 (n=7,629) *Note.* 95% confidence intervals for the marginal predicted mean psychological distress levels from the multilevel growth curve models. Results based on weighted and imputed data.

**Figure 3.**
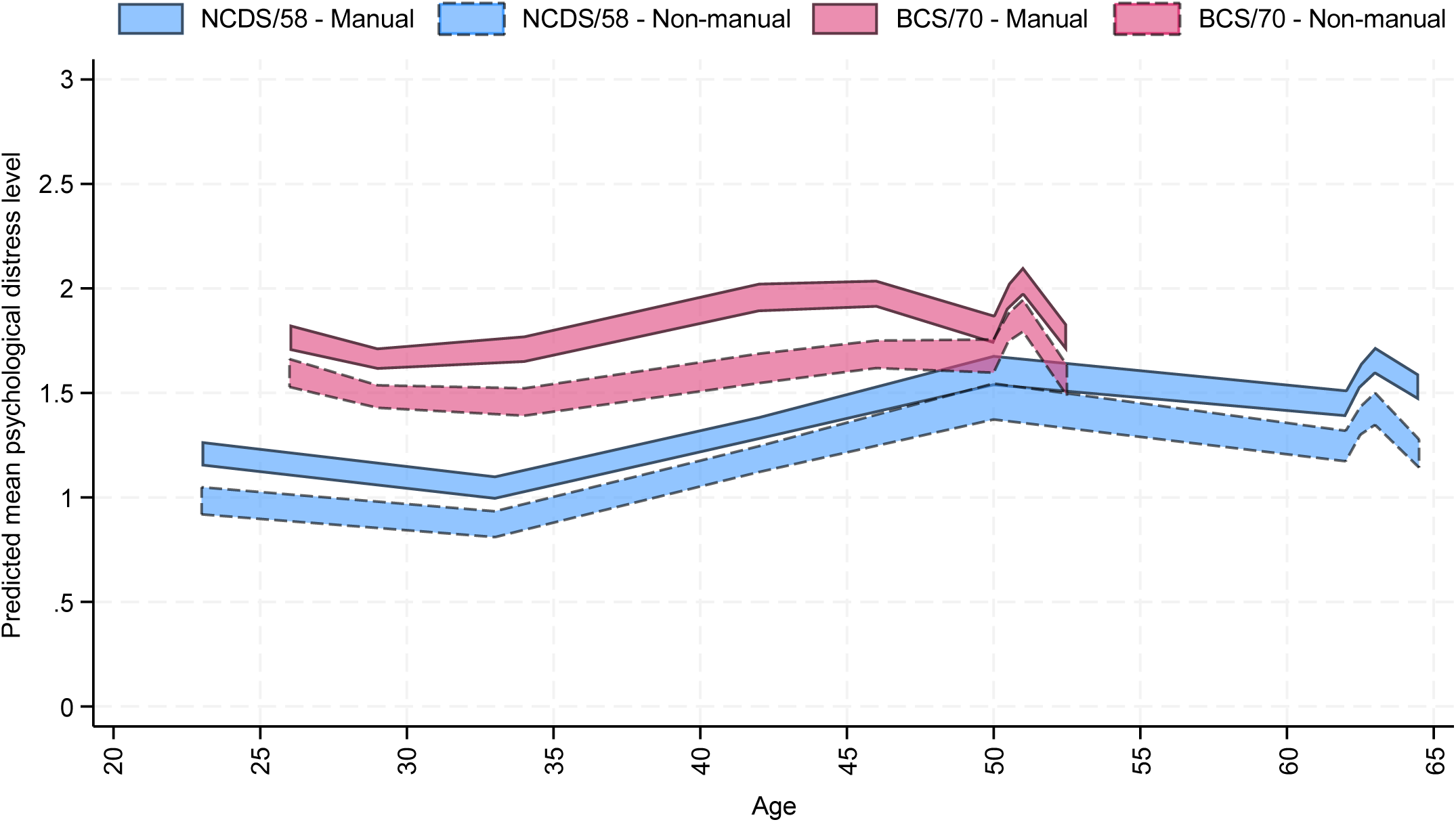
Long-term trajectories of psychological distress by parental social class during childhood (age 10/11) across NCDS/58 (n=6,553) and BCS/70 (n=7,629) *Note.* 95% confidence intervals for the marginal predicted mean psychological distress levels from the multilevel growth curve models. Results based on weighted and imputed data.

**Figure 4.**
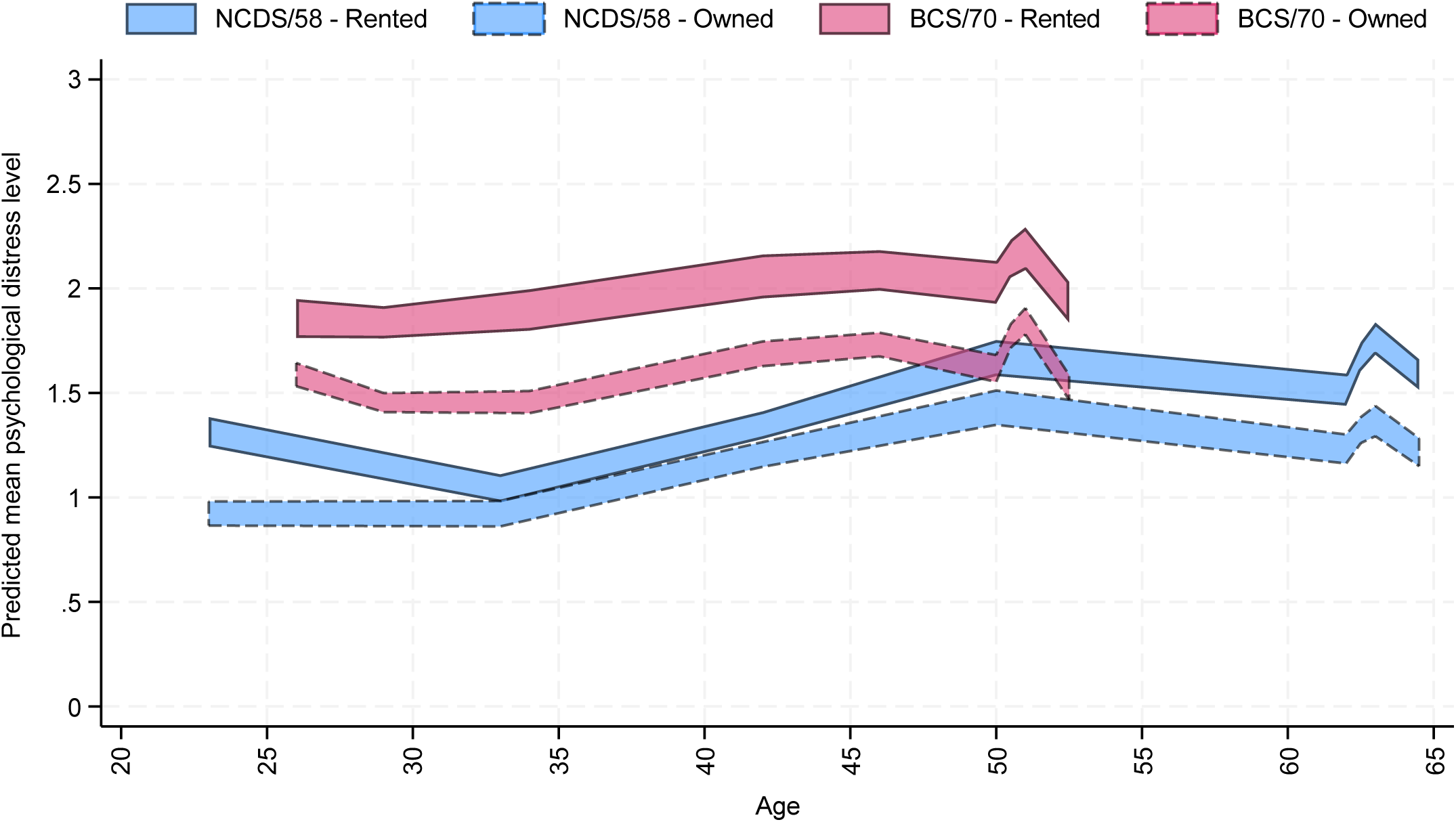
Long-term trajectories of psychological distress by childhood housing tenure (ages 5&10/7&11) across NCDS/58 (n=6,553) and BCS/70 (n=7,629) *Note.* 95% confidence intervals for the marginal predicted mean psychological distress levels from the multilevel growth curve models. Results based on weighted and imputed data.

In NCDS/58, inequalities in the starting points were also found at the intersection of gender and childhood housing tenure (B_women*owned7&11_NCDS/58_=−0.27 [−0.49, −0.05], *p*=0.014) (**Figure 5**) and, to a lesser extent, at the intersection of gender and childhood parental social class (B_women*non-manual_NCDS/58_=−0.21 [−0.42, 0.00], *p*=0.048) (**Figure 6**), although in this latter case they were almost non-statistically significant at the 95%CI, showing larger childhood socioeconomic position inequalities in women than in men. Evidence suggestive of inequalities at those intersections in the starting points was not found in BCS/70.

**Figure 5.**
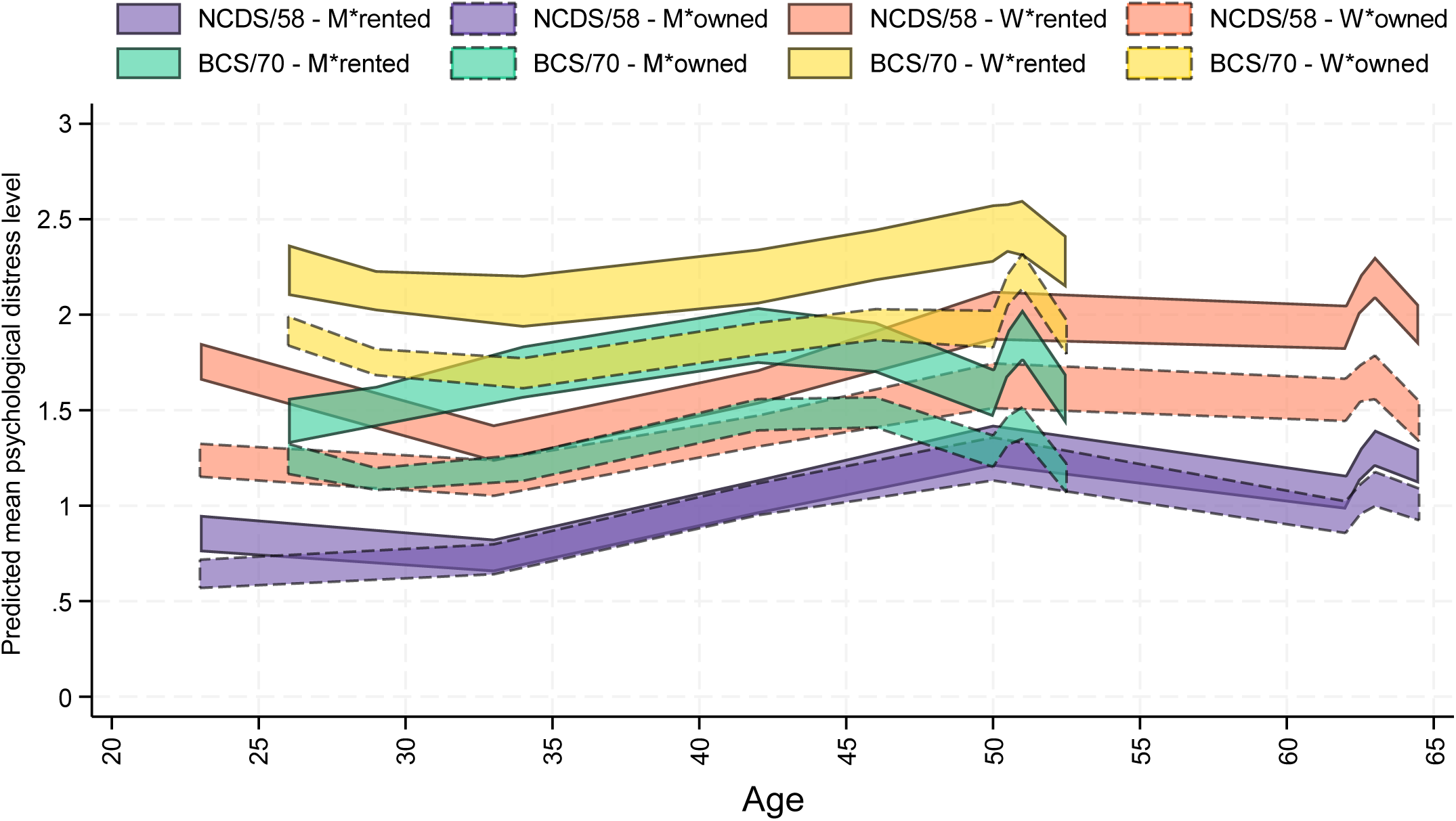
Long-term trajectories of psychological distress by gender and childhood housing tenure (ages 5&10/7&11) across NCDS/58 (n=6,553) and BCS/70 (n=7,629) *Note.* M: men; W: women. 95% confidence intervals for the marginal predicted mean psychological distress levels from the multilevel growth curve models. Results based on weighted and imputed data.

**Figure 6.**
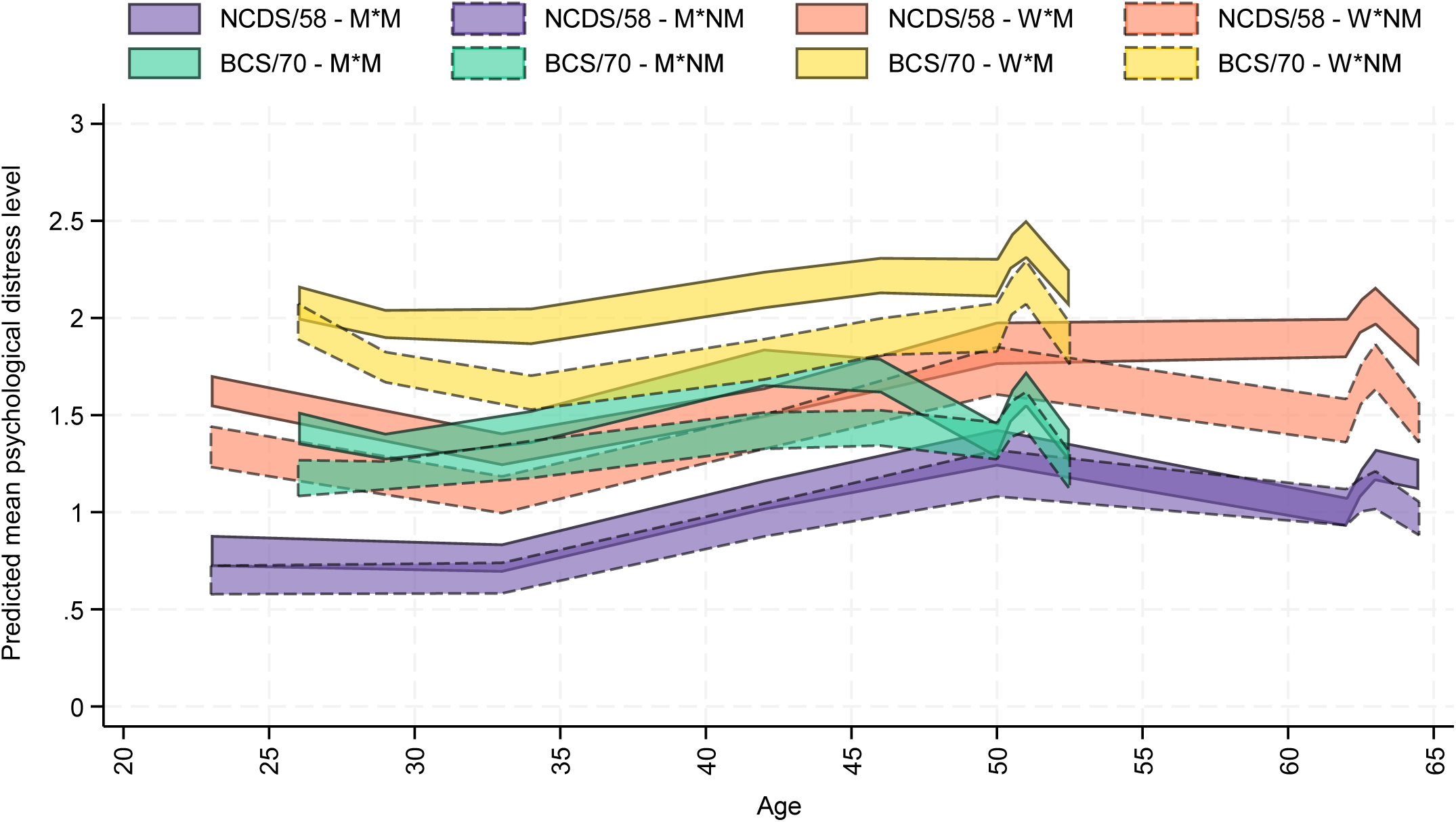
Long-term trajectories of psychological distress by gender and parental social class during childhood (age 10/11) across NCDS/58 (n=6,553) and BCS/70 (n=7,629) *Note.* M*: men; *M: manual parental social class; *NM: non-manual parental social class; W*: women. 95% confidence intervals for the marginal predicted mean psychological distress levels from the multilevel growth curve models. Results based on weighted and imputed data.

In NCDS/58, a larger linear decrease among women (B_spline1_linear*women_NCDS/58_=−0.04 [−0.07, −0.01], *p*=0.020) in the first segment of the long-term trajectory (up to the COVID-19 pandemic onset) was reflected in the gender gap decreasing towards the age of 30 (**Figure 2**). We did not find evidence of significant differences in the change over time by childhood socioeconomic position or their intersection with gender in any of the two cohorts.

The coefficients from the long-term trajectory models (multilevel growth curve models), are available in **eAppendix 8 (Supplementary Material)**.

Models estimated with continuous age and with the extended sample provided very similar results. Estimates and plots of the marginal mean predicted levels from these models are available in **eAppendix 9** and **eAppendi× 10 (Supplementary Material)**, respectively.

### Differences across key time-points

**Table 2** shows the estimates and 95% CIs from the analyses comparing the psychological distress levels and gaps between the most recent survey sweeps in both birth cohorts and three key time-points.

**Table 2.** Estimates and 95% confidence intervals (CIs) from analyses comparing psychological distress levels between most recent main sweeps and earliest time-point, most recent pre-pandemic assessment, or point of highest psychological distress during COVID-19 pandemic.

|  | NCDS/58 (n=6,553) |  |  |  |  |  |
| --- | --- | --- | --- | --- | --- | --- |
|  | First (age 23)<br>vs last (age 64.5) |  | Pre-pandemic (age 50)<br>vs last (age 64.5) |  | Pandemic (age 62.5)<br>vs last (age 64.5) |  |
|  | B (95% CI) | p | B (95% CI) | p | B (95% CI) | p |
| **Overall** |  |  |  |  |  |  |
| Time | 0.35 (0.26, 0.45) | <0.001 | 0.01 (-0.07, 0.09) | 0.791 | -0.16 (-0.23, -0.08) | <0.001 |
| **Gender (ref.: Men)** |  |  |  |  |  |  |
| Time | 0.41 (0.28, 0.54) | <0.001 | 0.00 (-0.13, 0.12) | 0.974 | -0.06 (-0.17, 0.04) | 0.248 |
| Women | 0.72 (0.61, 0.83) | <0.001 | 0.58 (0.45, 0.70) | <0.001 | 0.79 (0.67, 0.91) | <0.001 |
| Time * Women | -0.12 (-0.30, 0.06) | 0.208 | 0.03 (-0.15, 0.21) | 0.765 | -0.19 (-0.35, -0.04) | 0.016 |
| **Parental social class (ref.: Manual)** |  |  |  |  |  |  |
| Time | 0.38 (0.25, 0.50) | <0.001 | 0.08 (-0.04, 0.20) | 0.179 | -0.13 (-0.23, -0.02) | 0.019 |
| Non-manual | -0.24 (-0.35, -0.12) | <0.001 | -0.10 (-0.23, 0.04) | 0.162 | -0.21 (-0.34, -0.08) | 0.002 |
| Time * Non-manual | -0.07 (-0.26, 0.13) | 0.505 | -0.20 (-0.39, -0.01) | 0.039 | -0.08 (-0.24, 0.07) | 0.287 |
| **Gender (ref.: Men) and parental social class (ref.: Manual)** |  |  |  |  |  |  |
| Time | 0.45 (0.28, 0.63) | <0.001 | 0.07 (-0.11, 0.24) | 0.452 | -0.01 (-0.15, 0.13) | 0.882 |
| Women | 0.81 (0.66, 0.95) | <0.001 | 0.62 (0.45, 0.79) | <0.001 | 0.90 (0.73, 1.06) | <0.001 |
| Non-manual | -0.11 (-0.27, 0.05) | 0.175 | -0.03 (-0.22, 0.16) | 0.734 | -0.06 (-0.23, 0.12) | 0.532 |
| Time * Women | -0.16 (-0.41, 0.09) | 0.210 | 0.03 (-0.22, 0.27) | 0.823 | -0.24 (-0.45, -0.03) | 0.026 |
| Time * Non-manual | -0.13 (-0.41, 0.16) | 0.377 | -0.20 (-0.47, 0.08) | 0.162 | -0.15 (-0.37, 0.07) | 0.176 |
| Women * Non-manual | -0.25 (-0.50, -0.01) | 0.044 | -0.12 (-0.39, 0.15) | 0.397 | -0.29 (-0.56, -0.02) | 0.034 |
| Time * Women * Non-manual | 0.13 (-0.31, 0.56) | 0.563 | 0.00 (-0.38, 0.38) | 0.983 | 0.14 (-0.20, 0.47) | 0.417 |
| **Childhood housing tenure (ref.: Rented at both time-points)** |  |  |  |  |  |  |
| Time | 0.35 (0.22, 0.49) | <0.001 | 0.05 (-0.07, 0.18) | 0.401 | -0.17 (-0.28, -0.05) | 0.007 |
| Owned at both time-points | -0.35 (-0.47, -0.23) | <0.001 | -0.24 (-0.38, -0.10) | 0.001 | -0.35 (-0.50, -0.21) | <0.001 |
| Time * Owned at both time-points | 0.00 (-0.19, 0.20) | 0.977 | -0.10 (-0.30, 0.09) | 0.294 | 0.00 (-0.17, 0.16) | 0.961 |
| **Gender (ref.: Men) and childhood housing tenure (ref.: Rented at both time-points)** |  |  |  |  |  |  |
| Time | 0.36 (0.18, 0.53) | <0.001 | 0.02 (-0.18, 0.22) | 0.846 | -0.07 (-0.23, 0.08) | 0.363 |
| Women | 0.83 (0.66, 0.99) | <0.001 | 0.76 (0.55, 0.96) | <0.001 | 0.99 (0.79, 1.20) | <0.001 |
| Owned at both time-points | -0.19 (-0.34, -0.05) | 0.007 | -0.06 (-0.24, 0.13) | 0.545 | -0.16 (-0.34, 0.03) | 0.102 |
| Time * Women | 0.00 (-0.27, 0.26) | 0.980 | 0.07 (-0.21, 0.34) | 0.627 | -0.18 (-0.43, 0.06) | 0.144 |
| Time * Owned at both time-points | 0.03 (-0.21, 0.28) | 0.781 | -0.10 (-0.36, 0.16) | 0.463 | -0.02 (-0.24, 0.20) | 0.882 |
| Women * Owned at both time-points | -0.27 (-0.50, -0.04) | 0.021 | -0.33 (-0.60, -0.06) | 0.017 | -0.35 (-0.64, -0.07) | 0.016 |
| Time * Women * Owned at both time-points | -0.07 (-0.44, 0.31) | 0.733 | -0.01 (-0.38, 0.36) | 0.968 | 0.01 (-0.32, 0.35) | 0.932 |
| **BCS/70 (n=7,629)** |  |  |  |  |  |  |
|  | **First (age 26)**<br>**vs last (age 52.5)** |  | **Pre-pandemic (age 46)**<br>**vs last (age 52.5)** |  | **Pandemic (age 50.5)**<br>**vs last (age 52.5)** |  |
|  | **B (95% CI)** | **p** | **B (95% CI)** | **p** | **B (95% CI)** | **p** |
| **Overall** |  |  |  |  |  |  |
| Time | 0.02 (-0.07, 0.11) | 0.644 | -0.08 (-0.15, -0.01) | 0.022 | -0.33 (-0.41, -0.26) | <0.001 |
| **Gender (ref.: Men)** |  |  |  |  |  |  |
| Time | 0.00 (-0.12, 0.11) | 0.975 | -0.22 (-0.32, -0.13) | <0.001 | -0.33 (-0.44, -0.22) | <0.001 |
| Women | 0.73 (0.63, 0.84) | <0.001 | 0.48 (0.36, 0.59) | <0.001 | 0.75 (0.63, 0.87) | <0.001 |
| Time * Women | 0.04 (-0.12, 0.21) | 0.600 | 0.28 (0.13, 0.42) | <0.001 | 0.00 (-0.15, 0.15) | 0.984 |
| **Parental social class (ref.: Manual)** |  |  |  |  |  |  |
| Time | 0.02 (-0.11, 0.14) | 0.783 | -0.08 (-0.17, 0.02) | 0.105 | -0.34 (-0.46, -0.22) | <0.001 |
| Non-manual | -0.27 (-0.40, -0.15) | <0.001 | -0.24 (-0.37, -0.11) | <0.001 | -0.26 (-0.41, -0.12) | <0.001 |
| Time * Non-manual | 0.01 (-0.19, 0.20) | 0.939 | -0.01 (-0.16, 0.14) | 0.891 | 0.02 (-0.16, 0.19) | 0.862 |
| **Gender (ref.: Men) and parental social class (ref.: Manual)** |  |  |  |  |  |  |
| Time | -0.05 (-0.22, 0.12) | 0.566 | -0.22 (-0.36, -0.08) | 0.002 | -0.34 (-0.51, -0.17) | <0.001 |
| Women | 0.73 (0.56, 0.89) | <0.001 | 0.55 (0.38, 0.72) | <0.001 | 0.84 (0.65, 1.02) | <0.001 |
| Non-manual | -0.30 (-0.48, -0.11) | 0.002 | -0.15 (-0.33, 0.02) | 0.089 | -0.17 (-0.38, 0.04) | 0.107 |
| Time * Women | 0.14 (-0.11, 0.39) | 0.280 | 0.29 (0.09, 0.49) | 0.005 | 0.00 (-0.22, 0.22) | 0.993 |
| Time * Non-manual | 0.12 (-0.16, 0.40) | 0.382 | 0.00 (-0.22, 0.21) | 0.981 | 0.02 (-0.24, 0.27) | 0.901 |
| Women * Non-manual | 0.03 (-0.24, 0.31) | 0.814 | -0.17 (-0.41, 0.08) | 0.187 | -0.19 (-0.48, 0.09) | 0.180 |
| Time * Women * Non-manual | -0.23 (-0.65, 0.19) | 0.279 | -0.03 (-0.34, 0.28) | 0.860 | 0.00 (-0.37, 0.36) | 0.991 |
| **Childhood housing tenure (ref.: Rented at both time-points)** |  |  |  |  |  |  |
| Time | 0.08 (-0.11, 0.27) | 0.399 | 0.00 (-0.16, 0.16) | 0.960 | -0.32 (-0.48, -0.16) | <0.001 |
| Owned at both time-points | -0.31 (-0.47, -0.15) | <0.001 | -0.28 (-0.45, -0.12) | 0.001 | -0.37 (-0.54, -0.19) | <0.001 |
| Time * Owned at both time-points | -0.15 (-0.36, 0.07) | 0.177 | -0.13 (-0.32, 0.05) | 0.164 | -0.04 (-0.23, 0.15) | 0.680 |
| **Gender (ref.: Men) and childhood housing tenure (ref.: Rented at both time-points)** |  |  |  |  |  |  |
| Time | 0.07 (-0.18, 0.32) | 0.592 | -0.18 (-0.38, 0.03) | 0.100 | -0.41 (-0.63, -0.19) | <0.001 |
| Women | 0.86 (0.61, 1.12) | <0.001 | 0.54 (0.28, 0.80) | <0.001 | 0.70 (0.43, 0.98) | <0.001 |
| Owned at both time-points | -0.25 (-0.45, -0.04) | 0.020 | -0.24 (-0.46, -0.02) | 0.031 | -0.39 (-0.64, -0.15) | 0.001 |
| Time * Women | 0.02 (-0.34, 0.39) | 0.892 | 0.34 (0.04, 0.64) | 0.024 | 0.17 (-0.14, 0.48) | 0.273 |
| Time * Owned at both time-points | -0.12 (-0.41, 0.16) | 0.397 | -0.08 (-0.32, 0.17) | 0.533 | 0.08 (-0.19, 0.34) | 0.568 |
| Women * Owned at both time-points | -0.12 (-0.42, 0.18) | 0.419 | -0.09 (-0.39, 0.22) | 0.588 | 0.05 (-0.28, 0.39) | 0.751 |
| Time * Women * Owned at both time-points | -0.05 (-0.46, 0.37) | 0.831 | -0.10 (-0.45, 0.25) | 0.571 | -0.22 (-0.59, 0.15) | 0.235 |

*Note.* BCS/70: 1970 British Cohort Study; B: coefficient; CI: confidence interval; NCDS/58: 1958 National Child Development Study; p: significance level. Results based on weighted and imputed data. Analyses including childhood housing tenure exclude cohort members who lived in a rented or owned house at only one of the time-points; sample size in these analyses is $n_{\text{NCDS/58}}=4,703$ and $n_{\text{BCS/70}}=5,106$ .

Differences between the most recent pre-pandemic assessments and the latest sweeps (ages 50/64.5 in NCDS/58 and 46/52.5 in BCS/70) were not significant in NCDS/58 (B_prepandemic_time_NCDS/58_=0.01 [−0.07, 0.09], *p*=0.791) and marginally significant and negative in BCS/70 (B_prepandemic_time_BCS/70_=−0.08 [−0.15, −0.01], *p*=0.022), in line with the notion of a “bounce back” to (or even slight improvement compared to) pre-pandemic levels.

In addition to the above-reported inequalities at earliest time-points (akin to the starting points in the long-term trajectory analyses, age 23 in NCDS/58 and 26 in BCS/70) by gender, parental social class during childhood, and childhood housing tenure (and their intersections in NCDS/58), we found similar gaps at the height of the COVID-19 pandemic (age 62.5 in NCDS/58 and 50.5 in BCS/70). Similar results were found in the comparisons with the pre-pandemic time-point, although in this case the gap by parental social class during childhood was not significant in NCDS/58 (B_non-manual_NCDS/58_=−0.10 [−0.23, 0.04], *p*=0.162), widening again towards the post-pandemic period (B_prepandemic_time*non-manual_NCDS/58_=−0.20 [−0.39, −0.01], *p*=0.039). In NCDS/58 there was a larger decrease in psychological distress between the pandemic and latest time-point among women than men (B_pandemic_time*women_NCDS/58_=−0.19 [−0.35, −0.04], *p*=0.016). In BCS/70, however, we found evidence suggestive of a widening gender gap when comparing the pre-pandemic and most recent assessments (B_prepandemic_time*women_BCS/70_=0.28 [0.13, 0.42], *p<*0.001).

The pooled analyses with the additional cohort terms (main effects and interactions) showed that, in line with **Figure 1**, overall levels of psychological distress were significantly higher in BCS/70 than NCDS/58 in the three time comparisons. The cohort gap was significantly smaller in the most recent sweep when compared to the earliest time-point and to the gap during the pandemic (B_first_time*BCS/70_=−0.33 [−0.45, −0.21], *p*<0.001; B_pandemic_time*BCS/70_=−0.18 [−0.29, −0.07], *p*=0.002), but not significantly different when using the most recent pre-pandemic assessment as the comparison time-point (B_prepandemic_time*BCS/70_=−0.09 [−0.20, −0.01], *p*=0.087). The full results from the pooled analyses are available in **eAppendix 11 (Supplementary Material)**.

### Between-person post-lockdown analyses

We found a significant relationship between inflation and psychological distress in the post-lockdown period, and we did not find evidence that this relationship significantly varied by gender, (concurrent) socioeconomic position, or their intersection. Detailed results from these analyses are available in **eAppendix 12** and **eAppendix 13 (Supplementary Material)**.

## Discussion

In this study, we used data from two long-standing birth cohorts, representative of the British population born in 1958 and 1970, with three aims: first, to understand how long-term trajectories of mental (ill-)health evolved during the post-lockdown period, which overlapped with large cost-of-living increases; second, to examine differences in the trajectories by generation, gender, childhood socioeconomic position, and their intersections; and, third, to explore differences in mental (ill-)health by cost-of-living levels. Regarding the first and second aims, we found that, after a period of increased distress coinciding with the COVID-19 pandemic, levels reduced towards the post-lockdown period. These post-lockdown reductions were larger among the younger cohort (born in 1970 vs 1958) and among women from the 1958 cohort than men. Across most of the adult lifespan of both cohorts (ages 23-64.5 in NCDS/58 and 26-52.5 in BCS/70, spanning 41.5 and 26.5 years, respectively), women and those from a disadvantaged childhood socioeconomic position had, on average, higher levels of distress than men and those from an advantaged childhood socioeconomic position. We also found evidence suggesting that the gaps across childhood socioeconomic position were larger in women from the older cohort (NCDS/58). In most cases, we did not find evidence of different changes over time by gender, childhood socioeconomic position, or their intersection. The only evidence we found suggesting a reduction of these gaps was for women from the older cohort (NCDS/58) between the height of the pandemic and the latest data collection around 2022. In turn, partial evidence suggested a widening in the gender gaps in the younger cohort (BCS/70) between the pre- and post-pandemic periods, as well as a temporary reduction in the gaps by parental social class in childhood at age 50 in the older cohort (NCDS/58), just to widen again towards the most recent (post-pandemic) sweep. Finally, regarding the third aim, we found that inflation was associated with the incidence of psychological distress symptoms, but we did not find evidence suggesting that this relationship substantially varied by gender, concurrent socioeconomic position, or their intersections.

Taken altogether, these results offer a mixed view of the population mental health in post-lockdown Britain. The two generations of adults under study seem to be ‘bouncing back’ from the high levels of psychological distress experienced during the pandemic (4). However, our results also suggest that the improvement is smaller in the older cohort (NCDS/58). This is consistent with emerging evidence on depressive symptomatology in the pandemic aftermath among English adults aged 50 and older, with larger representation of adults aged 60-74 (50). However, it would not be expected based on previous cross-generational evidence including older (1946) cohorts at similar ages (4, 34) or from evidence on mental health around retirement age in Britain (51), both of which would have suggested an improvement in mental health around this age. Indeed, although the younger cohort (BCS/70, ‘gen-X’) had higher levels of psychological distress than the older cohort (NCDS/58, ‘baby boomers’) throughout the lifespan, in line with previous cross-generational evidence (4, 34), our study suggests that those gaps may be decreasing towards older ages. It is possible that this is a result of experiencing multiple population-wide shocks (e.g., COVID-19 pandemic and cost-of-living increases) at a particularly sensitive period (retirement age). However, this is uncharted territory in a scholarship otherwise suggesting a decline in multiple health (including mental health) outcomes in younger generations (20, 21), so further continued monitoring is needed.

We also found evidence suggesting a narrowing gender gap in psychological distress in the NCDS/58 cohort between the height of the pandemic and the post-lockdown period. On the one hand, it is possible that psychological distress levels for men in their 60s are declining or not improving as fast as for women in recent years. Considering the challenging economic situation and the crucial transition into retirement, this could be consistent with previous evidence suggesting that, although women’s mental health tends to be poorer at all time-points, men’s mental health can deteriorate more during economic crises (52) or with multiple inflation hardships (53), in line with sexist conceptions of the man as the ‘breadwinner’. This could also be consistent with the finding that these and other (but not all) sexist conceptions and attitudes have declined in Britain over the last few decades (54), which could reflect in them having a larger influence over older cohorts. On the other hand, however, the gender gaps in the most recent NCDS/58 data collection were not different when compared to the ones earlier in life (ages 23 and 50). This suggests that these ‘improvements’ may be due to the fact that, since the pandemic came with a widening of the pre-existing gender inequalities (4), women had a longer way to ‘bounce back’, without really impacting the gender gaps in the long run. Unfortunately, this is consistent with the findings in BCS/70, where the gender gaps in the post-lockdown period were not significantly different than during the height of the pandemic, earlier in life (age 26) or, more dishearteningly, compared to prior to the pandemic, where part of our evidence actually suggests a widening in the gender gaps. A reason for this lack of improvement in the long run may be found in the same report on the change in (some) sexist attitudes, which suggests that behavioural changes have not followed attitudinal ones, with the division of domestic labour, for instance, still resting mostly on women’s shoulders (54).

Like gender inequalities, childhood socioeconomic inequalities persisted throughout the entire long-term trajectories, without consistent evidence of these narrowing. On the one hand, we found that the proportion of people in the relatively disadvantaged childhood socioeconomic position was smaller in the younger generation, meaning that the proportion of individuals in the socioeconomically disadvantaged long-term trajectory has decreased across generations. On the other hand, we provide one of the longest-term empirical studies to date, which suggests early life socioeconomic disadvantage continues to influence mental health inequalities even after more than half a century. As a fundamental cause of health (including mental health) inequalities (12), early life socioeconomic inequalities can persist across the life course (and transmit across life courses) due to complex chains of privilege and disadvantage which start even before birth (55). Our study adds to a growing field supporting the notion that early life prevention of socioeconomic disadvantage will be crucial, in order to reduce life-course inequalities and prevent further intergenerational transmission of health inequalities (11). In light of the growing number of children in poverty in the UK (56), immediate measures to reduce socioeconomic adversity may be needed to prevent these disadvantage cycles to further reproduce into generations currently in their infancy and youth (57). This is also consistent with the results of our analysis focused on the post-lockdown period, which suggests that inflation had a negative relationship with mental health during this period and that most of the inequalities by gender and concurrent socioeconomic position were independent of inflation levels or predating the period with highest inflation levels.

At the intersection of both gender and childhood socioeconomic inequalities, we found that mental health inequalities by childhood socioeconomic position were larger in women than in men, but only in the older (NCDS/58) cohort. We did not find evidence of these intersectional inequalities decreasing over time within that cohort, and we did not find evidence of such inequalities in the younger (BCS/70) cohort. These cohort differences may be, in part, due to increases in educational attainment and, relatedly, labour market participation among women across generations (54), suggesting that these inequalities can be prevented.

### Strengths and limitations

We used data from two long-standing birth cohort studies, representing two generations (‘Baby Boomers’ and ‘Gen X’) of the British population. To the best of our knowledge, this is the longest population-based study of long-term trajectories of mental (ill-)health within the same individuals, with trajectories spanning up to more than four decades. We used a robust approach to the measurement of mental (ill-)health across the life course, ensuring that the measures were equivalent not only over time within the same individuals but also by gender, childhood socioeconomic position, cohort, and their intersections, an aspect that is often overlooked in quantitative research on intersectional inequalities (36). We also used the rich information collected from individuals across their life courses to inform our approach to dealing with missing data (43). By using these approaches, we maximised the plausibility that our results are generalisable to the British population at these ages.

However, the results from our study must be interpreted considering some limitations. First, our findings may not be generalisable to other countries, particularly those with substantially different geopolitical contexts and economic conditions and constraints, and which make up most of the global population, as well as to other sectors within the British adult population, including migrant and displaced communities or ethnically minoritised groups. Second, we used a relatively simple approach to categorising individuals into gender and socioeconomic groups. While we are aware that there is a larger complexity in how these social identities and positions can be approached (and even categorised) (58), we aimed to maximise comparability across the cohorts, minimise the presence of missing data, ensure temporal ordering, and limit model complexity. However, we acknowledge that these conform to an inherently limited set of intersecting identities and positions, as well as variables and categories (e.g., (58)), that are provisionally adopted as proxies of the systems of oppression underlying any of the inequalities we found (37). Third, we may have been limited in terms of statistical power to detect some ‘effects’ (in statistical terms), particularly those involving multiple interactions, which include most intersectional terms. We coupled the analytical approaches with data visualisations to avoid exclusively relying on statistical tests. The visualisations were generally consistent with the observation that social inequalities have not substantially reduced across the life course. It is important to note that the absence of an ‘intersectional effect’ does not imply the absence of intersectional experiences (59). Furthermore, future studies may use other methods (e.g., three-level intersectional MAIHDA models (60)) to either replicate our findings or extend them to additional intersectional strata.

## Conclusions

This study shows that, after a period of increased levels of psychological distress during the early phases of the COVID-19 pandemic, levels of psychological distress in British adults born in 1958 and 1970 have reduced close to pre-pandemic levels. Our study also confirms long-range impacts of childhood socioeconomic disadvantage on adult mental health even after more than half a century, mental health inequalities by gender at all time points and, in the 1958 cohort, inequalities at the intersection of childhood socioeconomic position and gender, that persisted for most of the life course. Although inflation was associated with psychological distress, inequalities by gender and adult socioeconomic position were independent of (or predated high levels of) inflation. This suggests that efforts must be made to reduce gender and socioeconomic inequalities from early in life, coupled with interventions (and further research into optimal interventions) to reverse unjust inequalities in population mental health.

## Data Availability

All data used in this study are freely available to bona fide researchers at the UK Data Service (https://doi.org/10.5255/UKDA-Series-200001 and https://doi.org/10.5255/UKDA-Series-2000032).

https://doi.org/10.5255/UKDA-Series-2000032

https://osf.io/msprq/

## Author contributions

DMA: conceptualisation, data curation, formal analysis, methodology, visualisation, writing – original draft preparation, writing – review & editing.

GBP: conceptualisation, funding acquisition, supervision, writing – review & editing.

JDM: conceptualisation, funding acquisition, supervision, writing – review & editing.

## Acknowledgements

We would like to thank the members of the NCDS/58 and BCS/70 cohorts for generously giving up their time across their life courses, as well as the Centre for Longitudinal Studies team members for collecting, managing, and making these data useable and accessible.

The 1958 National Child Development Study (NCDS/58) and the 1970 British Cohort Study (BCS/70) are supported by the Centre for Longitudinal Studies, Resource Centre 2015-2020 [ES/M001660/1] and 2022 [ES/W013142/1] grants, along a host of other co-funders. The COVID-19 data collections were funded by the UKRI grant Understanding the economic, social and health impacts of COVID-19 using lifetime data: evidence from 5 nationally representative UK cohorts [ES/V012789/1].

The views expressed are those of the authors and not necessarily those of the Wellcome Trust, ESRC, King’s College London, or other funders.

