## Supplementary Material for "COVID-19, economic downturn, and long-term trajectories of population mental health: evidence from two nationally representative British birth cohorts at the intersection of gender and socioeconomic position"

### eAppendix 1. Measurement invariance/equivalence (MI/E) testing approach and results

#### Rationale

One of the key assumptions in a study like ours, in which comparisons were going to be made across time-points within the same individuals, but also across groups within cohorts, and across cohorts, is that that measures of the construct under study (psychological distress) are being measured in an invariant or equivalent way [1, 2].

Previous evidence using data on the same measure (nine-item version of the Malaise Inventory) in the same cohorts (1958 National Child Development Study [NCDS/58] and 1970 British Cohort Study [BCS/70]) has shown that the measure is, indeed, equivalent across time-points within individuals, across genders, and even across cohorts at similar/same ages [3-5]. However, our study includes new data from these birth cohorts and, additionally, deals with comparisons across childhood socioeconomic positions (SEP) and at the intersection of gender and childhood SEP. Therefore, we needed to extend the previous evidence to account for the most recent main survey data collection time-points (or *sweeps*) as well as for equivalence across childhood SEPs (both childhood parental social class and childhood housing tenure) and the gender*childhood SEP intersections. Importantly, quantitative research on intersectional inequalities has typically overlooked the key issue of ensuring that measurement invariance holds across intersectional strata and not only across each of the social identity and position groups that make up the strata [6].

In order to ensure comparisons of psychological distress levels across time-points and groups are not due to differences in the measurement characteristics or parameters, evidence of scalar (also known as 'strong') invariance is typically needed, meaning that item loadings and thresholds are equivalent across time-points and groups [1].

When using a measure like the nine-item version of the Malaise inventory, which includes nine different experiences of mental ill-health, a further, more restrictive level of invariance ('Rasch' invariance) can be helpful. This level of invariance is achieved when the loadings across items within each of the conditions (time-points and groups) are the same. This is helpful because, otherwise, a one-unit increase in the observed Malaise inventory may have different implications on the underlying level of psychological distress depending on what specific indicator/symptom has been endorsed. Therefore, if the measure shows to be invariant at the 'Rasch' level, this provides further evidence of the validity of using the sum-score of the scale, as all indicators have a similarly strong relationship with the underlying construct.

Therefore, in our study, we aimed to empirically test whether the Malaise inventory was invariant at least at the metric level and, ideally, at the 'Rasch' level.

**Analytical approach**

We used a multiple-group confirmatory factor analysis (CFA) approach to estimate different models with increasing levels of constraints [1]. First, 'configural invariance' models were estimated, in which the same factor structure is imposed across conditions without further constraints. These models were considered to have good model fit if their Root Mean Square Error of Approximation (RMSEA) values were below 0.060, and their Comparative Fit Index (CFI) and Tucker-Lewis Index (TLI) values were above 0.950 [7]. If the configural models showed appropriate fit, 'scalar invariance' models were estimated, in which item loadings and thresholds were constrained to be equal across conditions (time-points and groups). Due to increasing constraints, these models are expected to have poorer fit to the data than the configural ones. However, scalar invariance was considered to be achieved if that loss in model fit was smaller than 0.015 for the RMSEA and smaller than 0.010 for the CFI [8, 9]. Finally, 'Rasch invariance' models were estimated, in which all items/indicators were constrained to have the same loadings. The loss in fit compared to the scalar invariance models was assessed using the same criteria.

All models were estimated in Mplus version 8 [10] using a Weighted Least Squares Mean and Variance adjusted (WLSMV) estimator and a Delta parameterisation [11].

**Findings**

We found evidence of the longitudinal invariance of the nine-item Malaise Inventory across genders, childhood SEP indicators, and their intersection in both NCDS/58 and BCS/70, as well as across both birth cohorts.

A 'Rasch' level of invariance (equal loadings and thresholds across groups and time-points as well as same loadings across items) was reached in all cases except for the three-categories childhood housing tenure, where a 'scalar' invariance model (equal loadings and thresholds across groups) was supported by the data, but a more constrained 'Rasch' model resulted in either an invalid solution (in NCDS/58 and BCS/70) or a solution with a large loss in fit (in the model combining both cohorts). A reduced version of the childhood housing tenure variable, focused on the participants living in owned or rented accommodation at both time-points provided a valid and acceptable solution.

Details on the fit indices across the different models and conditions are available in the table below.

| Conditions: time-points * (cohorts) * groups | | Model | Chi-square (df) | RMSEA (90% CI) | CFI | TLI | ΔRMSEA | ΔCFI |
|---|---|---|---|---|---|---|---|---|
| **NCDS/58:** 8 time-points (ages 23, 33, 42, 50, 62, 62.5, 63, 64.5) * groups | Sex/gender (2) | Configural | 1330 (432) | 0.025 (0.024, 0.027) | 0.987 | 0.983 | | |
| | | Scalar | 1932 (537) | 0.028 (0.027, 0.029) | 0.980 | 0.979 | -0.003 | -0.007 |
| | | **Rasch** | **2028 (545)** | **0.029 (0.027, 0.030)** | **0.979** | **0.978** | **-0.001** | **-0.001** |
| | Childhood parental social class (2) | Configural | 1500 (432) | 0.027 (0.026, 0.029) | 0.986 | 0.982 | | |
| | | Scalar | 2083 (537) | 0.030 (0.028, 0.031) | 0.980 | 0.978 | -0.003 | -0.006 |
| | | **Rasch** | **2340 (545)** | **0.032 (0.030, 0.033)** | **0.977** | **0.976** | **-0.002** | **-0.003** |
| | Childhood housing tenure (3) | Configural | 2589 (648) | 0.037 (0.036, 0.039) | 0.975 | 0.967 | | |
| | | **Scalar** | **3268 (809)** | **0.037 (0.036, 0.039)** | **0.969** | **0.967** | **0.000** | **-0.006** |
| | | Rasch | Invalid solution | | | | | |
| | Childhood housing tenure reduced (2) | Configural | 1292 (432) | 0.025 (0.024, 0.027) | 0.989 | 0.985 | | |
| | | Scalar | 1836 (537) | 0.028 (0.027, 0.029) | 0.983 | 0.982 | -0.003 | -0.006 |
| | | **Rasch** | **2074 (545)** | **0.030 (0.029, 0.032)** | **0.980** | **0.979** | **-0.002** | **-0.003** |
| | Sex/gender (2) * childhood parental social class (2) | Configural | 2045 (864) | 0.029 (0.027, 0.030) | 0.984 | 0.979 | | |
| | | Scalar | 2842 (1081) | 0.032 (0.030, 0.033) | 0.977 | 0.975 | -0.003 | -0.007 |
| | | **Rasch** | **2896 (1089)** | **0.032 (0.030, 0.033)** | **0.976** | **0.975** | **0.000** | **-0.001** |
| | Sex/gender (2) * childhood housing tenure reduced (2) | Configural | 1760 (864) | 0.026 (0.024, 0.028) | 0.988 | 0.984 | | |
| | | Scalar | 2508 (1081) | 0.029 (0.028, 0.031) | 0.980 | 0.979 | -0.003 | -0.008 |
| | | **Rasch** | **2560 (1089)** | **0.030 (0.028, 0.031)** | **0.980** | **0.979** | **-0.001** | **0.000** |
| **BCS/70:** 9 time-points (ages 26, 29, 34, 42, 46, 50, 50.5, 51, 52.5) * groups | Sex/gender (2) | Configural | 1603 (486) | 0.028 (0.027, 0.030) | 0.987 | 0.983 | | |
| | | Scalar | 2184 (605) | 0.030 (0.029, 0.032) | 0.982 | 0.981 | -0.002 | -0.005 |
| | | **Rasch** | **2317 (613)** | **0.031 (0.030, 0.033)** | **0.981** | **0.980** | **-0.001** | **-0.001** |
| | Childhood parental social class (2) | Configural | 1760 (486) | 0.030 (0.029, 0.032) | 0.987 | 0.983 | | |
| | | Scalar | 2233 (605) | 0.031 (0.029, 0.032) | 0.983 | 0.982 | -0.001 | -0.004 |
| | | **Rasch** | **2464 (613)** | **0.032 (0.031, 0.034)** | **0.981** | **0.980** | **-0.001** | **-0.002** |
| | Childhood housing tenure (3) | Configural | 1950 (729) | 0.030 (0.028, 0.031) | 0.988 | 0.984 | | |
| | | **Scalar** | **2501 (911)** | **0.030 (0.029, 0.032)** | **0.984** | **0.983** | **0.000** | -0.004 |
| | | Rasch | Invalid solution | | | | | |
| | | Configural | 1621 (486) | 0.030 (0.028, 0.031) | 0.988 | 0.984 | | |

| | | | | | | | | |
|---|---|---|---|---|---|---|---|---|
| | Childhood housing tenure reduced (2) | Scalar | 2115 (605) | 0.031 (0.029, 0.032) | 0.984 | 0.982 | -0.001 | -0.004 |
| | | **Rasch** | **2274 (613)** | **0.032 (0.031, 0.033)** | **0.982** | **0.981** | **-0.001** | **-0.002** |
| | Sex/gender (2) * childhood parental social class (2) | Configural | 2255 (972) | 0.030 (0.029, 0.032) | 0.987 | 0.982 | | |
| | | Scalar | 2984 (1217) | 0.033 (0.031, 0.034) | 0.982 | 0.980 | -0.003 | -0.005 |
| | | **Rasch** | **3015 (1225)** | **0.032 (0.031, 0.033)** | **0.981** | **0.980** | **0.001** | **-0.001** |
| | Sex/gender (2) * childhood housing tenure reduced (2) | Configural | 2109 (972) | 0.030 (0.028, 0.031) | 0.987 | 0.983 | | |
| | | Scalar | 2809(1217) | 0.031 (0.030, 0.033) | 0.982 | 0.981 | -0.001 | -0.005 |
| | | **Rasch** | **2873 (1225)** | **0.032 (0.030, 0.033)** | **0.982** | **0.981** | **-0.001** | **0.000** |
| **NCDS/58 + BCS/70:** 4 time-points (ages 23/26, 33/34, 42, 50) * 2 cohorts * groups | Sex/gender (2) | Configural | 1230 (432) | 0.024 (0.023, 0.026) | 0.986 | 0.982 | | |
| | | Scalar | 1721 (537) | 0.026 (0.025, 0.028) | 0.980 | 0.978 | -0.002 | -0.006 |
| | | **Rasch** | **1785 (545)** | **0.027 (0.025, 0.028)** | **0.979** | **0.978** | **-0.001** | **-0.001** |
| | Childhood parental social class (2) | Configural | 1294 (432) | 0.025 (0.023, 0.027) | 0.987 | 0.982 | | |
| | | Scalar | 1825 (537) | 0.027 (0.026, 0.029) | 0.980 | 0.979 | -0.002 | -0.007 |
| | | **Rasch** | **1986 (545)** | **0.029 (0.027, 0.030)** | **0.978** | **0.977** | **-0.002** | **-0.002** |
| | Childhood housing tenure (3) | Configural | 1481 (648) | 0.025 (0.023, 0.026) | 0.988 | 0.983 | | |
| | | **Scalar** | **2011 (809)** | **0.026 (0.025, 0.028)** | **0.982** | **0.981** | **-0.001** | **-0.006** |
| | | Rasch | 3828 (817) | 0.042 (0.040, 0.043) | 0.975 | 0.973 | -0.016 | -0.007 |
| | Childhood housing tenure reduced (2) | Configural | 1215 (432) | 0.025 (0.023, 0.026) | 0.988 | 0.983 | | |
| | | Scalar | 1707 (537) | 0.027 (0.026, 0.029) | 0.981 | 0.980 | -0.002 | -0.007 |
| | | **Rasch** | **1869 (545)** | **0.029 (0.027, 0.030)** | **0.979** | **0.978** | **-0.002** | **-0.002** |
| | Sex/gender (2) * childhood parental social class (2) | Configural | 1828 (864) | 0.026 (0.025, 0.028) | 0.985 | 0.980 | | |
| | | Scalar | 2521 (1081) | 0.029 (0.027, 0.030) | 0.978 | 0.976 | -0.003 | -0.007 |
| | | **Rasch** | **2555 (1089)** | **0.029 (0.028, 0.031)** | **0.978** | **0.976** | **0.000** | **0.000** |
| | Sex/gender (2) * childhood housing tenure reduced (2) | Configural | 1755 (864) | 0.026 (0.025, 0.028) | 0.986 | 0.981 | | |
| | | Scalar | 2428 (1081) | 0.029 (0.027, 0.031) | 0.978 | 0.977 | -0.003 | -0.008 |
| | | **Rasch** | **2463 (1089)** | **0.029 (0.028, 0.031)** | **0.978** | **0.977** | **0.000** | **0.000** |

*Note*. BCS/70: 1970 British Cohort Study; CFI: Comparative Fit Index; df: degrees of freedom; NCDS/58: 1958 National Child Development Study; RMSEA: Root Mean Square Error of Approximation; TLI: Tucker-Lewis Index; ΔCFI: difference in CFI; ΔRMSEA: difference in RMSEA. Results based on weighted data. Selected models are highlighted in bold.

**eAppendix 2. Rationale and methodological approach for between-person analyses focused on the relationship between inflation and mental (ill-)health**

To supplement the within-person long-term trajectory analyses, we also wished to understand the potential shorter-term impacts of the cost-of-living increases and pre-existing sources of disadvantage. The aim of these analyses was to explore the relationship between inflation and population mental (ill-)health, and –in line with the main analyses– whether this relationship varied by generation, gender, socioeconomic position, and their intersections.

#### Measures

The same outcome measure (psychological distress as measured by the nine-item version of the Malaise Inventory) and the same approach to approaching gender as in the long-term trajectory analyses were used in these analyses.

However, instead of childhood socioeconomic position indicators used in the main analyses, we used two alternative indicators representing the cohort member's *concurrent* socioeconomic position: housing tenure, grouped into owner (outright), owner (with mortgage), and other arrangements; and self-rated financial situation, assessed with the question "How well would you say you personally are managing financially these days?", and grouped into "Living comfortably", "Doing all right", and "Just about getting by or finding it quite/very difficult". Descriptive information on these variables is available in the following table:

| | **NCDS/58, N = 6,140** | | **BCS/70, N = 7,856** | |
|---|---|---|---|---|
| **Gender, N (%)** | | | | |
| Women | 3,277 | 50.0% | 4,015 | 52.6% |
| Men | 3,276 | 50.0% | 3,614 | 47.4% |
| **Adult SEP, N (%)** | | | | |
| Adult housing tenure | | | | |
| Own outright | 4,556 | 69.5% | 2,036 | 26.7% |
| Own with mortgage | 942 | 14.4% | 4,062 | 53.2% |
| Other | 977 | 14.9% | 1,491 | 19.5% |
| *Missing* | *78* | *1.2%* | *40* | *0.5%* |
| Self-rated financial situation | | | | |
| Just about/quite/very difficult | 973 | 14.8% | 1,519 | 19.9% |
| Doing all right | 2,481 | 37.9% | 3,264 | 42.8% |
| Living comfortably | 3,059 | 46.7% | 2,804 | 36.8% |
| *Missing* | *40* | *0.6%* | *42* | *0.6%* |
| **Gender and adult SEP, N (%)** | | | | |
| Gender * adult housing tenure | | | | |

| | | | | |
|---|---|---|---|---|
| Women * own outright | 2,346 | 35.8% | 1,144 | 15.0% |
| Women * own with mortgage | 423 | 6.5% | 2,073 | 27.2% |
| Women * other | 475 | 7.2% | 776 | 10.2% |
| *Women * missing* | *33* | *0.5%* | *22* | *0.3%* |
| Men * own outright | 2,210 | 33.7% | 892 | 11.7% |
| Men * own with mortgage | 519 | 7.9% | 1,989 | 26.1% |
| Men * other | 502 | 7.7% | 715 | 9.4% |
| *Men * missing* | *45* | *0.7%* | *18* | *0.2%* |
| Gender * self-rated financial situation | | | | |
| Women * just about/quite/very difficult | 499 | 7.6% | 854 | 11.2% |
| Women * doing all right | 1,218 | 18.6% | 1,652 | 21.7% |
| Women * living comfortably | 1,537 | 23.5% | 1,474 | 19.3% |
| *Women * missing* | *23* | *0.4%* | *35* | *0.5%* |
| Men * just about/quite/very difficult | 474 | 7.2% | 665 | 8.7% |
| Men * doing all right | 1,263 | 19.3% | 1,612 | 21.1% |
| Men * living comfortably | 1,522 | 23.2% | 1,330 | 17.4% |
| *Men * missing* | *17* | *0.3%* | *7* | *0.1%* |

*Note.* BCS/70: 1970 British Cohort Study; N: frequency; NCDS/58: 1958 National Child Development Study; SD: standard deviation; SEP: socioeconomic position. Results based on unweighted data. Analytical samples exclude participants who took part in the latest main survey sweep prior to the COVID-19 pandemic onset (n=1,662 in NCDS/58 and n=116 in BCS/70) or within the period spanning the COVID-19 Surveys data collection (n=44 in BCS/70, interviewed between September-October 2020).

We augmented the latest sweep of the two birth cohorts with the UK Office for National Statistics (ONS) monthly Consumer Prices Index including owner occupiers' housing costs (CPIH) dataset [12], matching each individual's interview date with the CPIH three months before. CPIH is the most comprehensive measure of inflation available as it compiles a large sample of goods and services across multiple outlets in the UK, as well as owner occupiers' housing costs and Council Tax (a local tax on residential properties paid by occupiers regardless of ownership). The three-month lag was chosen upon inspection of the correlation between different CPIH lags and psychological distress in both birth cohorts, allowing a plausible yet not too protracted lag between exposure and outcome. The graph depicting the correlation between different CPIH lags and psychological distress scores (9-item Malaise Inventory) in NCDS/58 and BCS/70 are shown below:

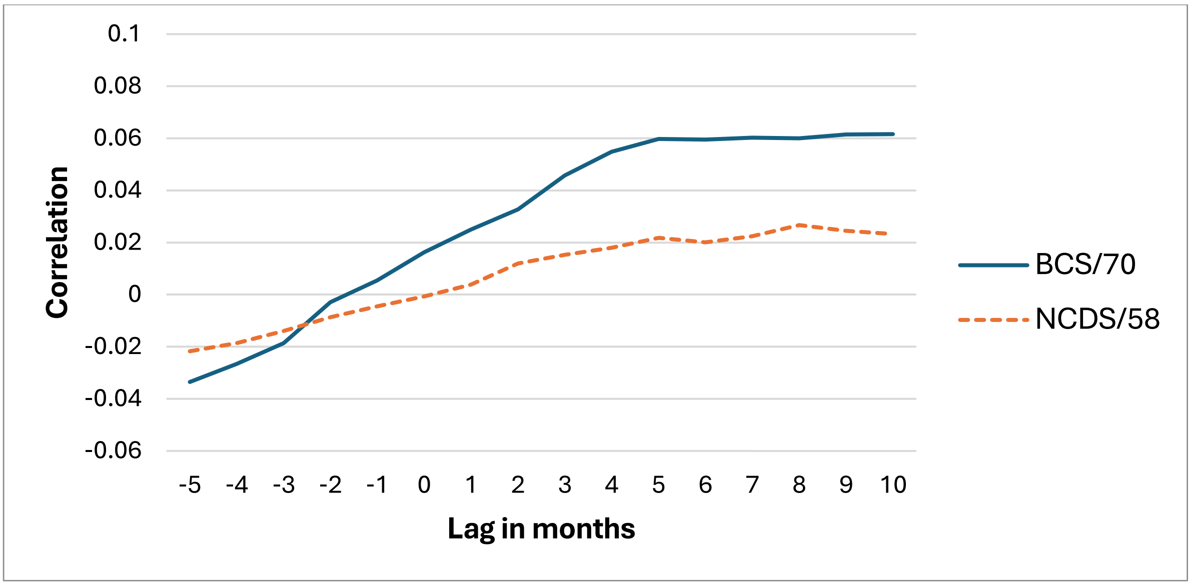

Note that a lag of zero (0) represents the correlation between the 9-item Malaise Inventory scores and the concurrent inflation level (as measured by the Consumer Prices Index including owner occupiers' housing costs, CPIH) of the same month in which the interview took place.

For the analyses during the post-lockdown period, potential confounders (including seasonality, childhood socioeconomic position, educational attainment, and youth and adulthood mental health, presence of long-standing illnesses, and living situation, among others) were identified based on previous literature [13, 14] and included in directed acyclic graphs (DAGs). A full list of the additional variables used in the study and the DAGs for these between-person post-lockdown analyses are included in **eAppendix 3** and **eAppendix 4 (Supplementary Material)**, respectively.

#### Statistical analysis

Negative binomial regression models were used to explore differences in mental ill-health by cost-of-living levels during the post-lockdown period, as well as variation in these differences by generation, gender, socioeconomic position, and their intersections.

In these analyses, we leveraged the variability in the dates of interviews in the latest main survey sweeps in both NCDS/58 and BCS/70. Two main sets of analyses were conducted to understand the potential between-person relationship between inflation and psychological distress. First, negative binomial regression models including the continuous CPIH level (lagged three months) as the main exposure and the Malaise inventory sum-score as outcome were estimated. Second, we assigned each participant to a group of 'unexposed' vs 'exposed' to 'very high inflation', based on whether they had been interviewed at a time when CPIH levels had been above 6 points for at least the three previous months (which was the case between June 2022 and December 2023). Due to this design, in addition to the participants excluded in the previous set of analysis, an additional 406 participants from NCDS/58 and 64 participants from BCS/70, whose interviews took place in January-April 2024, were excluded from the analyses with a binary CPIH operationalisation. Models were estimated separately for each birth cohort. First, general models for the overall cohort were estimated. Then, the potential variability in the relationship between inflation and psychological distress was explored by including the appropriate interaction terms between the exposure (i.e., the continuous CPIH or the binary 'very high inflation' variable) and the grouping variables (i.e., gender, each of the concurrent socioeconomic position indicators, or the intersection of between both). Unadjusted and adjusted models were estimated, based on literature-informed DAGs [13, 14] (see **eAppendix 4 (Supplementary Material)**.

In line with the main analyses (long-term trajectory analyses), both non-response weights and multiple imputation by chained equations (MICE) models were used to deal with missing data. In order to ensure congeniality between the imputation and analytical models, separate imputation models were used for each cohort and inequality under study (gender, tenure, self-rated financial position, gender * tenure, and gender * self-rated financial position).

The complete list of auxiliary variables included in the imputation models is available in **eAppendix 3 (Supplementary Material)**. Fifty imputed datasets were created, discarding the first 10 iterations of each chain. The analytical models were then conducted over the imputed datasets using Rubin's rules to pool the estimates and standard errors [15].

### eAppendix 3. Full list of variables used in the study

| Analysis | Variable | Name / dataset NCDS/58 | Name / dataset BCS/70 | Derivation / transformation |
|---|---|---|---|---|
| **Outcomes** | | | | |
| WP (all time-points) + BP (most recent time-point) | Sum-score of nine-item Malaise inventory | Derived from corresponding items at ages 23, 33, 42, 50, and 64.5 sweeps and COVID-19 survey waves. | Derived from corresponding items at ages 26, 29, 34, 42, 46, and 52.5 sweeps and COVID-19 survey waves | Sum score |
| **Exposures** | | | | |
| WP + BP | Gender | Sex assigned at birth (birth sweep). Complemented by additional sweeps if missing information. | Sex assigned at birth (birth sweep). Complemented by additional sweeps if missing information. | |
| WP | Parental social class during childhood | fclrg90 (CLOSER WP2, age 11) | fclrg90 (CLOSER WP2, age 10) | Recoded into 'Manual' and 'Non-manual' |
| WP | Housing tenure during childhood | tenure (CLOSER WP9, ages 7 & 11) | tenure (CLOSER WP9, ages 5 & 10) | |
| BP | Current self-rated financial situation | FINNOW (age 64.5 sweep) | b11finnow (age 52.5 sweep) | 'Just about getting by', 'Finding it quite difficult' and 'Finding it very difficult' combined into single group |
| BP | Current housing tenure | TENURE (age 64.5 sweep). Complemented by previous sweeps if unchanged. | b11ten (age 52.5 sweep). Complemented by previous sweeps if unchanged. | |
| BP | Inflation (CPIH) | Monthly CPIH (ONS dataset) | Monthly CPIH (ONS dataset) | |
| **Confounders** | | | | |
| BP | Seasonality | month of interview (age 64.5 sweep) | month of interview (age 52.5 sweep) | |
| BP | Gender | Sex assigned at birth (birth sweep). Complemented by additional sweeps if missing information. | Sex assigned at birth (birth sweep). Complemented by additional sweeps if missing information. | |
| BP | Parental social class | fclrg90 (CLOSER WP2, age 11) | fclrg90 (CLOSER WP2, age 10) | |

| | | | | |
|---|---|---|---|---|
| | during childhood | | | |
| BP | Cognitive ability (childhood) | n457 and n1840 (copying designs test and human figure drawing; age 7 sweep) | f119 and f121 (copying designs test and human figure drawing; age 5 sweep) | First principal component derived from principal component analysis |
| BP | Housing tenure during childhood | tenure (CLOSER WP9, ages 7 & 11) | tenure (CLOSER WP9, ages 5 & 10) | |
| BP | Youth mental health | intncdsz extncdsz (CLOSER WP9, age 16) | intbcsz extbcsz (CLOSER WP9, age 16) | |
| BP | Cohort member's social class in adulthood | clrg90 (CLOSER WP2, age 42) | clrg90 (CLOSER WP2, age 42) | |
| BP | Highest educational attainment | ND8HNVQ (age 50 sweep) | BD9HNVQ (age 42 sweep) | |
| BP | Cognitive ability (adulthood) | N8CFLISD N8CFANI and N8CFCOR (word list recall, timed letter search/cancellation, animal naming; age 50 sweep) | B10CFLISD B10CFANI and B10CFCOR (word list recall, timed letter search/cancellation, animal naming; age 46 sweep) | First principal component derived from principal component analysis |
| BP | Adult mental health | ND8MAL (age 50 sweep) | BD10MAL (age 46 sweep) | |
| BP | Self-rated financial situation before most recent main survey sweep | N9FINNOW (age 55 sweep) | B10FINNOW (age 46 sweep) | 'Just about getting by', 'Finding it quite difficult' and 'Finding it very difficult' combined into single group |
| BP | Housing tenure before most recent main survey sweep | Derived from N9TEN (age 55 sweep) | Derived from BD10TENURE (age 46 sweep) | Recoded into 'Owners/part-owners' and 'Not owners/part-owners' |
| BP | Long-standing illness | N9LOIL (age 55 sweep) | B10LOIL (age 46 sweep) | |
| BP | Economic activity | ND9ECACT (age 55 sweep) | BD10ECACT (age 46 sweep) | Recoded into 'Currently working' and 'Not currently working'. In |

| | | | | |
|---|---|---|---|---|
| | | | | NCDS/58, a third group including 'Retired' people was created and included in models wherever this did not affect model convergence |
| BP | Physical activity | N8EXERSE (age 50 sweep)<br>*Dichotomous variable reflecting whether the person reports exercising regularly or not.* | B10EXERSE (age 46 sweep)<br>*Number of days per week the person reports exercising.* | |
| BP | Living situation (living alone or not) | Derived from ND9HSIZE (age 55 sweep) | Derived from BD10HSIZE (age 46 sweep) | Recoded into 'Living alone' and 'Not living alone' |
| **Auxiliary variables** | | | | |
| WP | House overcrowding during childhood | Derived from crowd (CLOSER WP9; birth, age 7 & 11 sweeps) | Derived from crowd (CLOSER WP9; age 5 sweep) | Recoded into 'Up to 1 person per room' and 'Over 1 persons per room' |
| WP | Parental divorce during childhood | divorce (CLOSER WP9; up to age 16) | divorce (CLOSER WP9; up to age 16) | |
| WP | Low birth weight | lowbwt (CLOSER WP9, birth sweep) | lowbwt (CLOSER WP9, birth sweep) | |
| WP | Breastfed | brfed (CLOSER WP9, age 7 sweep) | brfed (CLOSER WP9, age 5 sweep) | |
| BP | Cognitive ability (cohort-specific information) | Derived from n914 and n917 (general ability; age 11 sweep) | Derived from i3617 i3618 i3619 i3620 i3621 i3622 i3623 i3624 i3625 i3626 i3627 i3628 i3629 i3630 i3631 i3632 i3633 i3634 i3635 i3636 i3637 i3638 i3639 i3640 i3641 i3642 i3643 and i3644 (BAS matrix tasks; age 10 sweep) | Sum |
| BP | Voted in last general elections | N9VOTE10 (age 55 sweep) | B9SCQ6 (age 42 sweep) | |

| | | | | |
|---|---|---|---|---|
| BP | Membership in organisations | Derived from N8ORGN01 N8ORGN02 N8ORGN03 N8ORGN04 N8ORGN05 N8ORGN06 N8ORGN07 N8ORGN08 N8ORGN09 N8ORGN10 N8ORGN11 N8ORGN12 N8ORGN13 N8ORGN14 N8ORGN15 and N8ORGN16 (current member of different organisations; age 42 sweep) | Derived from B9SCQ8A B9SCQ8B B9SCQ8C B9SCQ8D B9SCQ8E B9SCQ8F B9SCQ8G B9SCQ8H B9SCQ8I B9SCQ8J B9SCQ8K B9SCQ8L B9SCQ8M B9SCQ8N and B9SCQ8O (current member of different organisations; age 42 sweep) | Sum |
| BP | Provided consent for collection of biomarkers information | blcmea (age 44 sweep) | B10BSWILL (age 46 sweep) | |
| BP | Partnership status | Derived from ND9MS, ND9COHAB, and ND9PARTP (age 55 sweep) | Derived from BD10MS, BD10COHAB, and BD10PARTP (age 46 sweep) | Recoded into 'In a partnership (regardless of cohabitation)' and 'Not in a partnership' |
| BP | BMI | ND9BMI (age 55 sweep) | BD10BMI (age 46 sweep) | |
| BP | Self-rated health | N9HLTHGN (age 55 sweep) | B10HLTHGN (age 46 sweep) | |
| BP | Smoking status | Derived from N9SMOKIG (age 55 sweep) | Derived from B10SMOKIG (age 46 sweep) | Recoded into 'Never smoked', 'Ever smoked (not currently)', and 'Currently smokes' |
| BP | Social support | N8LISTEN (age 50 sweep) | B10LISTEN (age 46 sweep) | |
| BP | Number of non-responses to previous sweeps | Derived from OUTCME01 OUTCME02 OUTCME03 OUTCME04 OUTCME05 OUTCME06 | Derived from OUTCME01 OUTCME02 OUTCME03 OUTCME04 OUTCME05 OUTCME06 | Sum |

| | | | |
|---|---|---|---|
| | | OUTCME07<br>OUTCME08<br>OUTCME09 (response dataset) | OUTCME07<br>OUTCME08<br>OUTCME09<br>OUTCME10<br>(response dataset) |

*Note*. BCS/70: 1970 British Cohort Study; BP: between-person post-lockdown analyses; NCDS/58: 1958 National Child Development Study; WP: within-person long-term trajectory analyses. Due to convergence issues, some of the auxiliary variables for the between-person post-lockdown analyses were not included in some of the imputation models. In NCDS/58, these were: consent for biomarker collection and partnership status in overall models and models by gender; housing tenure in most recent pre-pandemic sweep, organisations membership, and non-responses to previous sweeps in models by tenure; non-responses to previous sweeps in models by financial situation; living situation, organisations membership, consent for biomarker collection, highest educational attainment, housing tenure in most recent pre-pandemic sweep, and non-responses to previous sweeps in models by the intersection of gender and tenure; and living situation, organisations membership, consent for biomarker collection, physical exercise, and non-responses to previous sweeps in models by the intersection of gender and financial situation. In BCS/70, these were: partnership status in models by tenure; partnership status and non-responses to previous sweeps in models by financial situation; housing tenure in most recent pre-pandemic sweep, economic activity, living situation, partnership status, and non-responses to previous sweeps in models by the intersection of gender and financial situation; and partnership status and non-responses to previous sweeps in models by gender and tenure.

**eAppendix 4. Directed Acyclic Graphs (DAGs) for the between-person post-lockdown analyses**

The following figures depict the DAGs for the between-person post-lockdown analyses. The main exposures appear in green shade, whereas the outcome appears in blue shade. Key pathways of interest are highlighted in **purple colour**, and the (groups of) variables that could confound the relationship are shaded in grey. The (groups of) variables that may not confound the relationship or may be on the pathway (and hence, adjusting for them could attenuate the relationship of interest) are included in white shade.

**Overall**

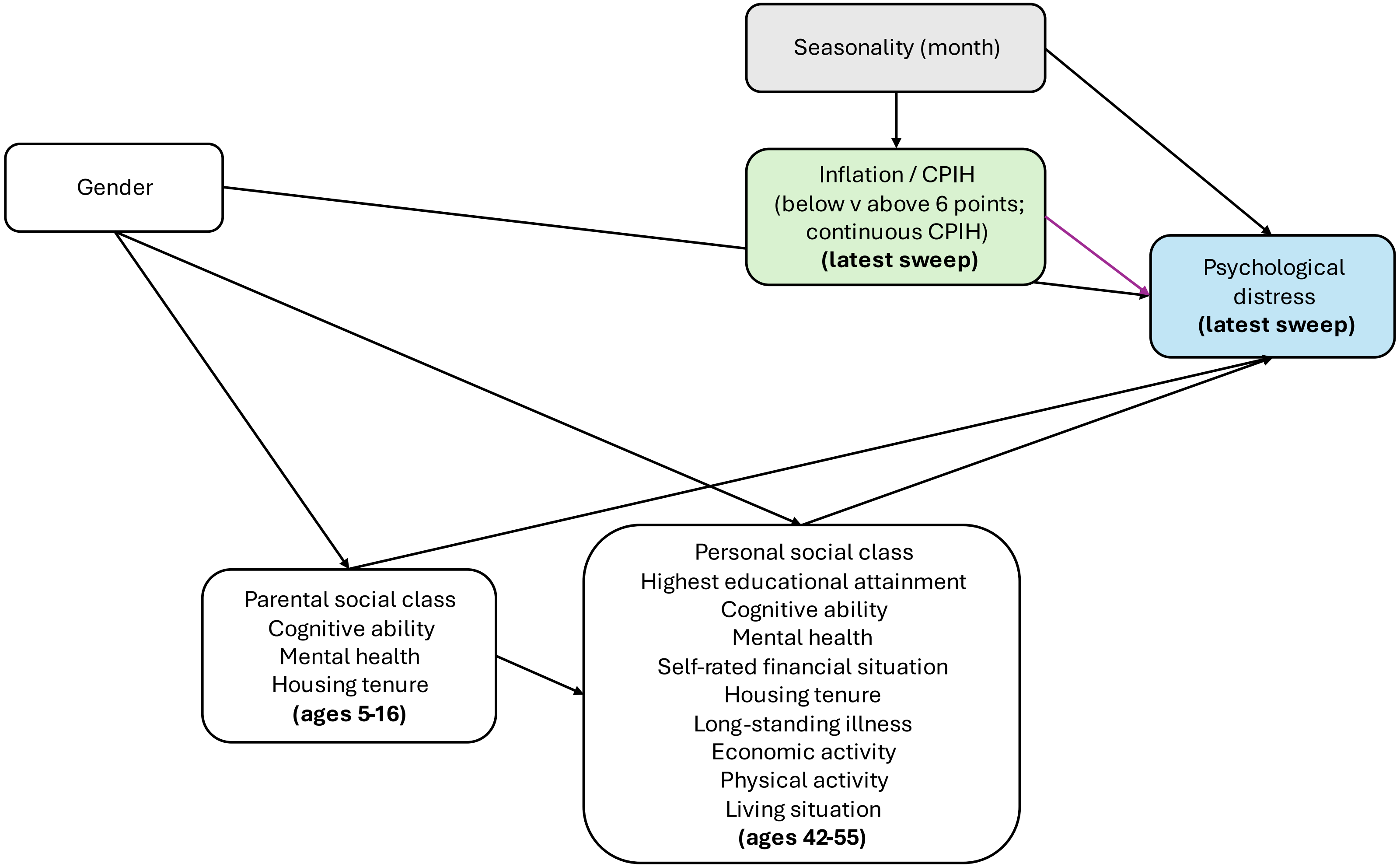

Seasonality (month)
Gender
Inflation / CPIH
(below v above 6 points;
continuous CPIH)
(latest sweep)
Psychological
distress
(latest sweep)
Parental social class
Cognitive ability
Mental health
Housing tenure
(ages 5-16)
Personal social class
Highest educational attainment
Cognitive ability
Mental health
Self-rated financial situation
Housing tenure
Long-standing illness
Economic activity
Physical activity
Living situation
(ages 42-55)

**By gender**

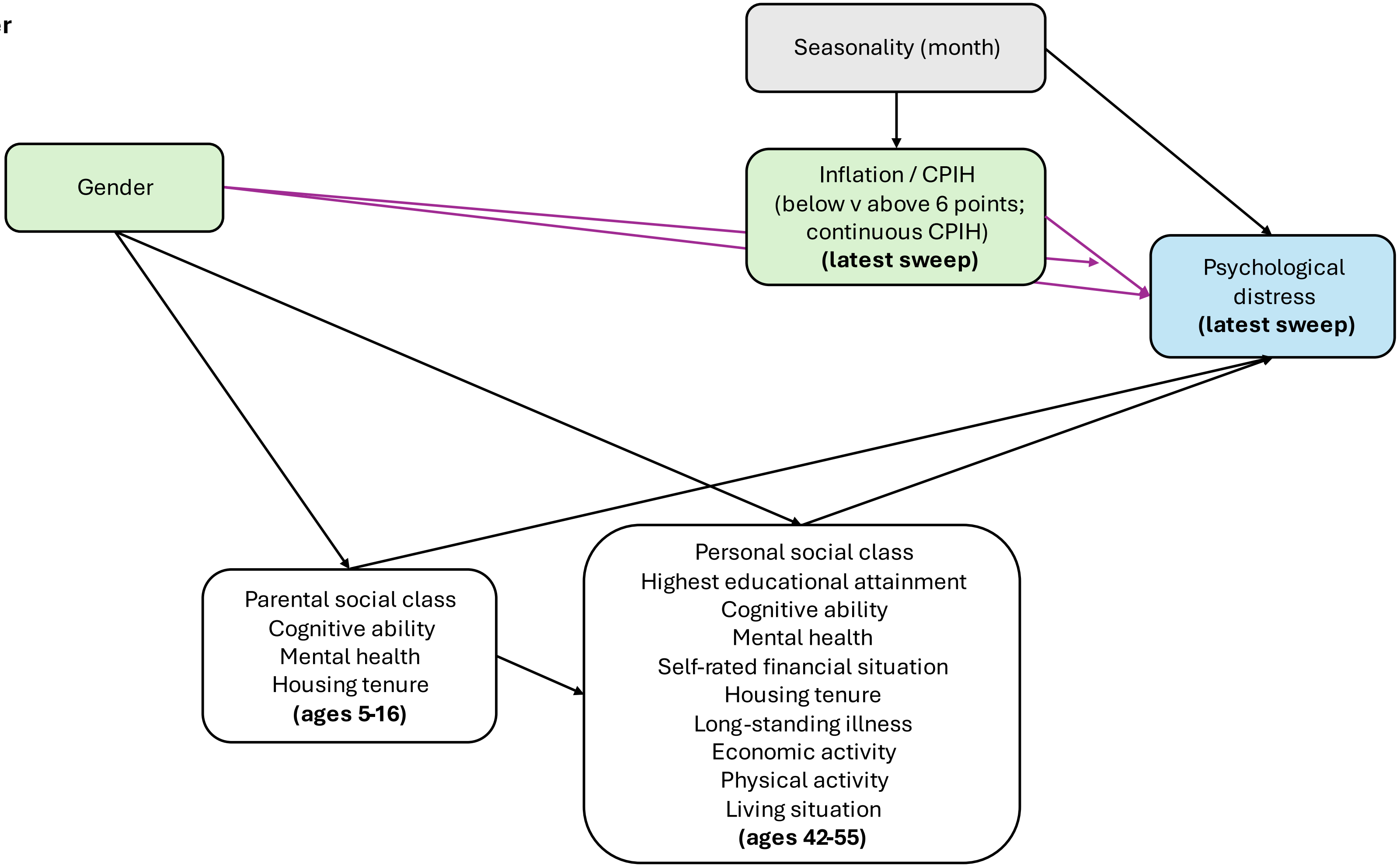

Seasonality (month)
Gender
Inflation / CPIH
(below v above 6 points;
continuous CPIH)
(latest sweep)
Psychological
distress
(latest sweep)
Parental social class
Cognitive ability
Mental health
Housing tenure
(ages 5-16)
Personal social class
Highest educational attainment
Cognitive ability
Mental health
Self-rated financial situation
Housing tenure
Long-standing illness
Economic activity
Physical activity
Living situation
(ages 42-55)

**By housing tenure**

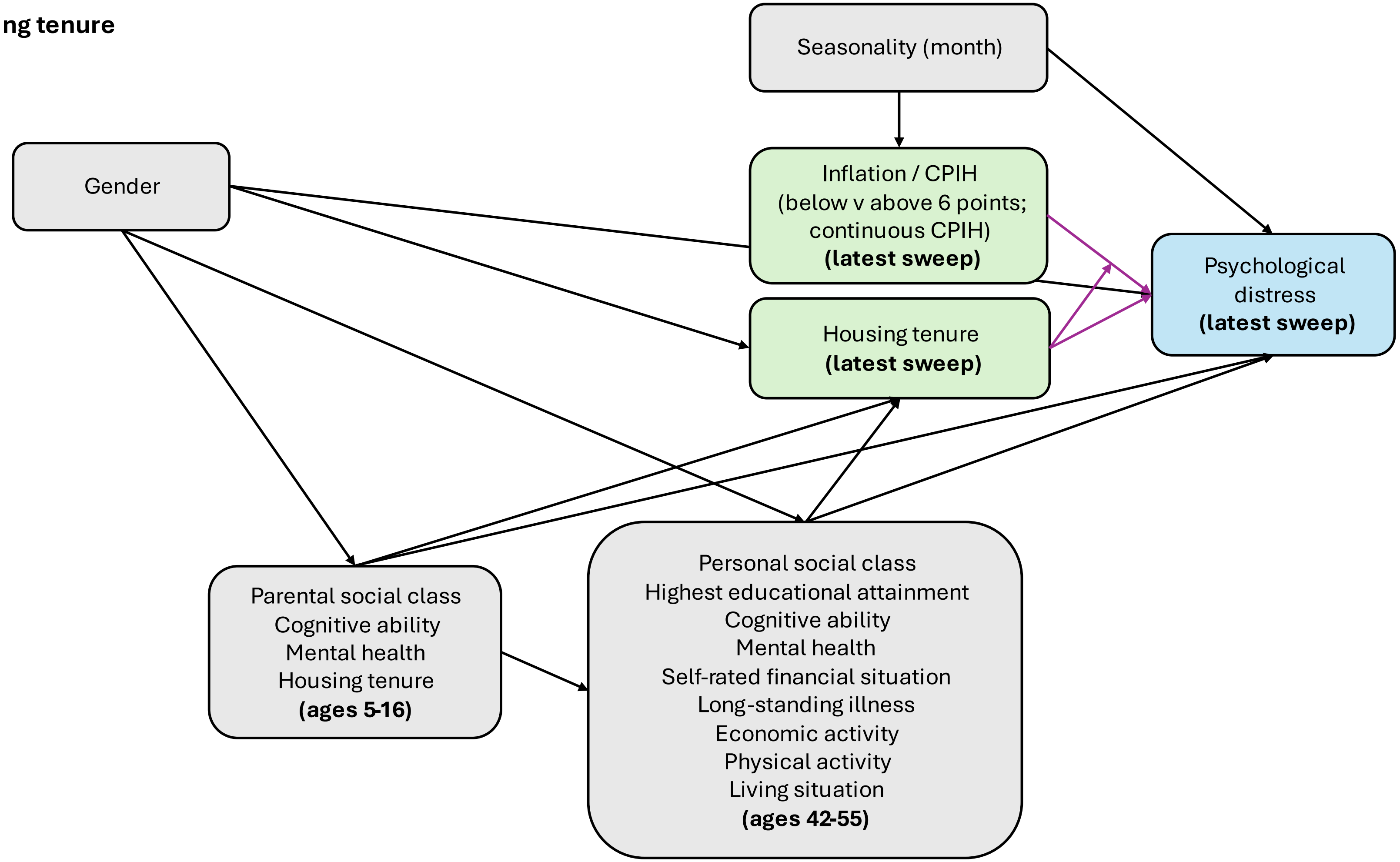

**By self-rated financial situation**

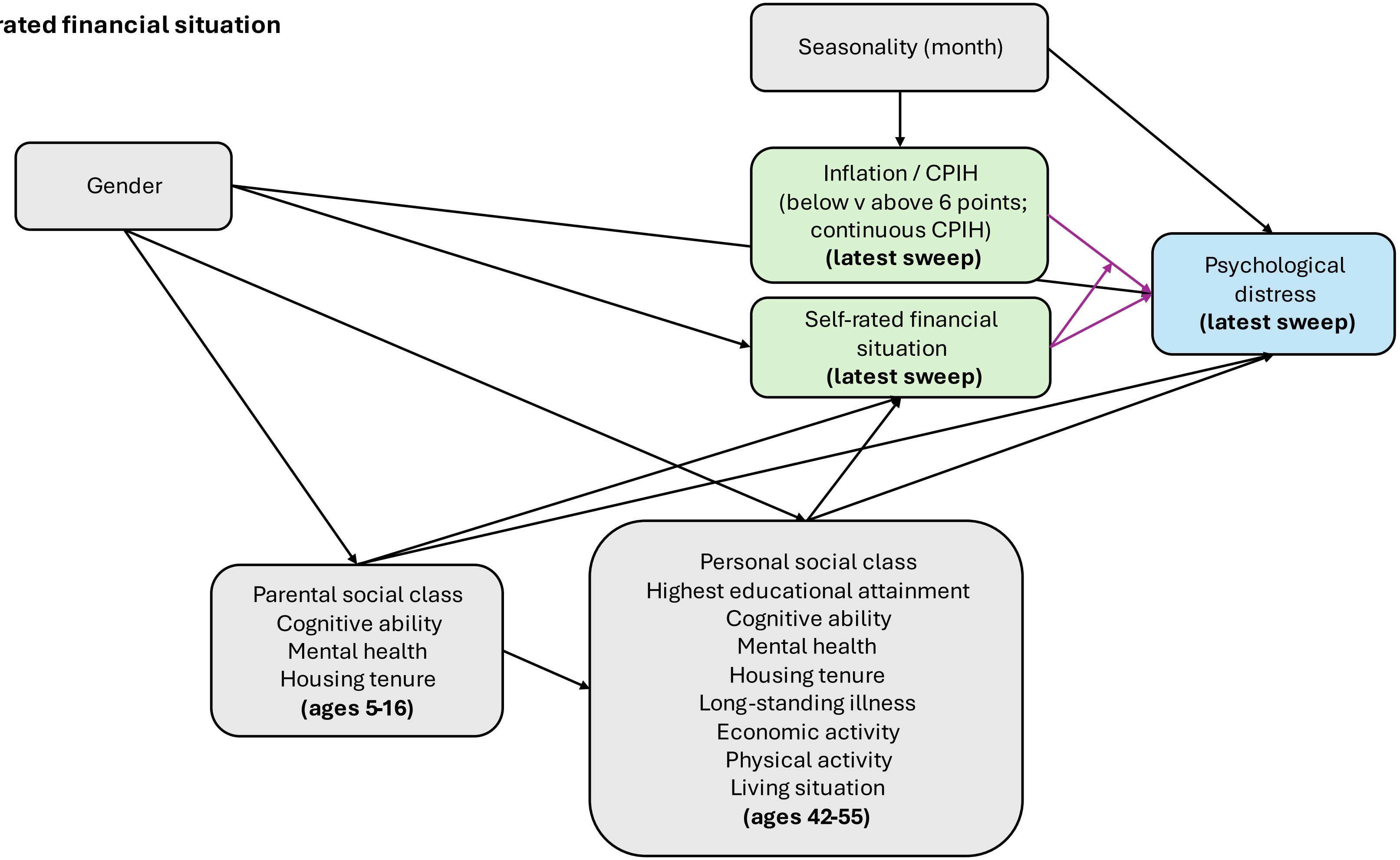

**By gender and self-rated financial situation**

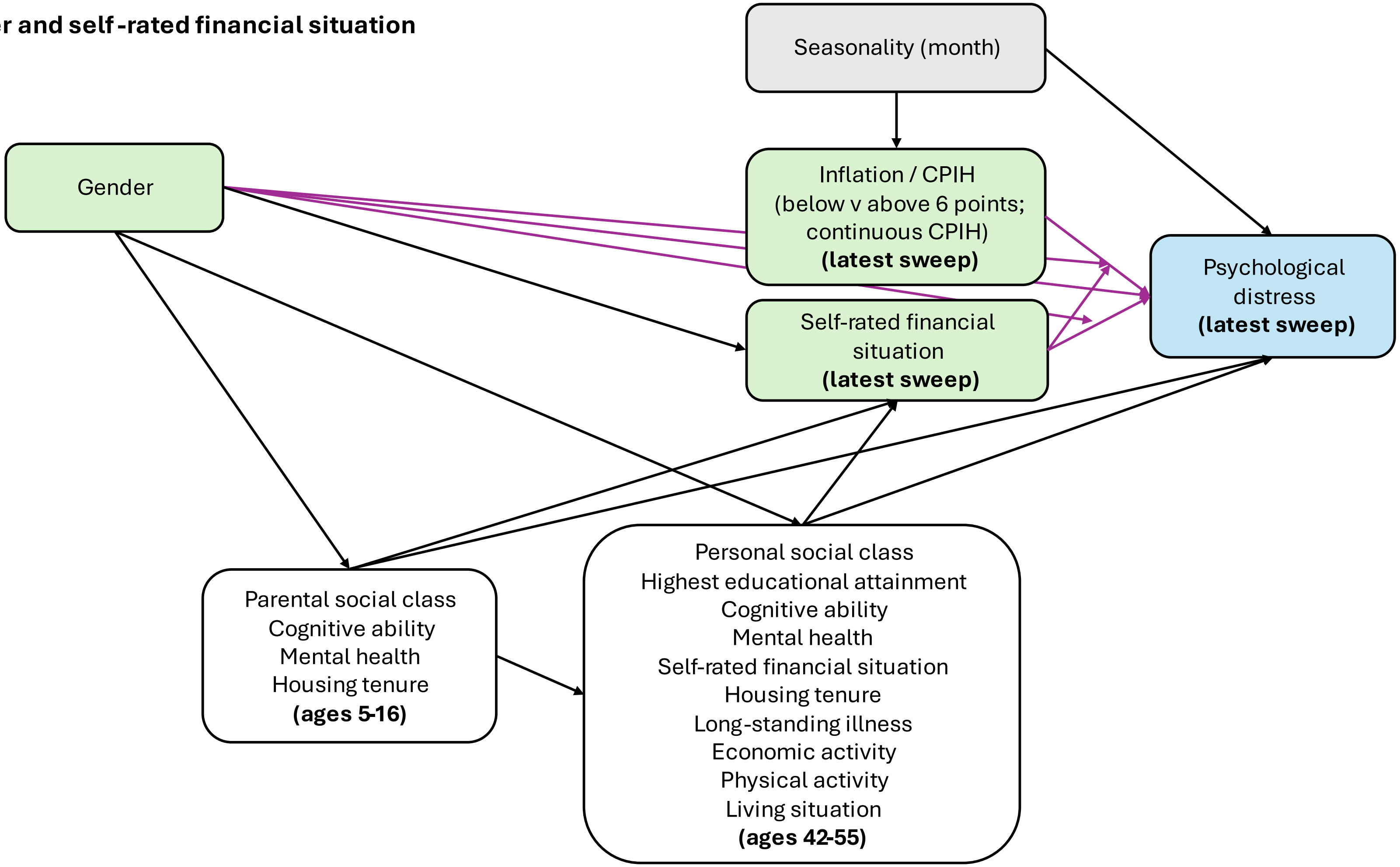

**By gender and housing tenure**

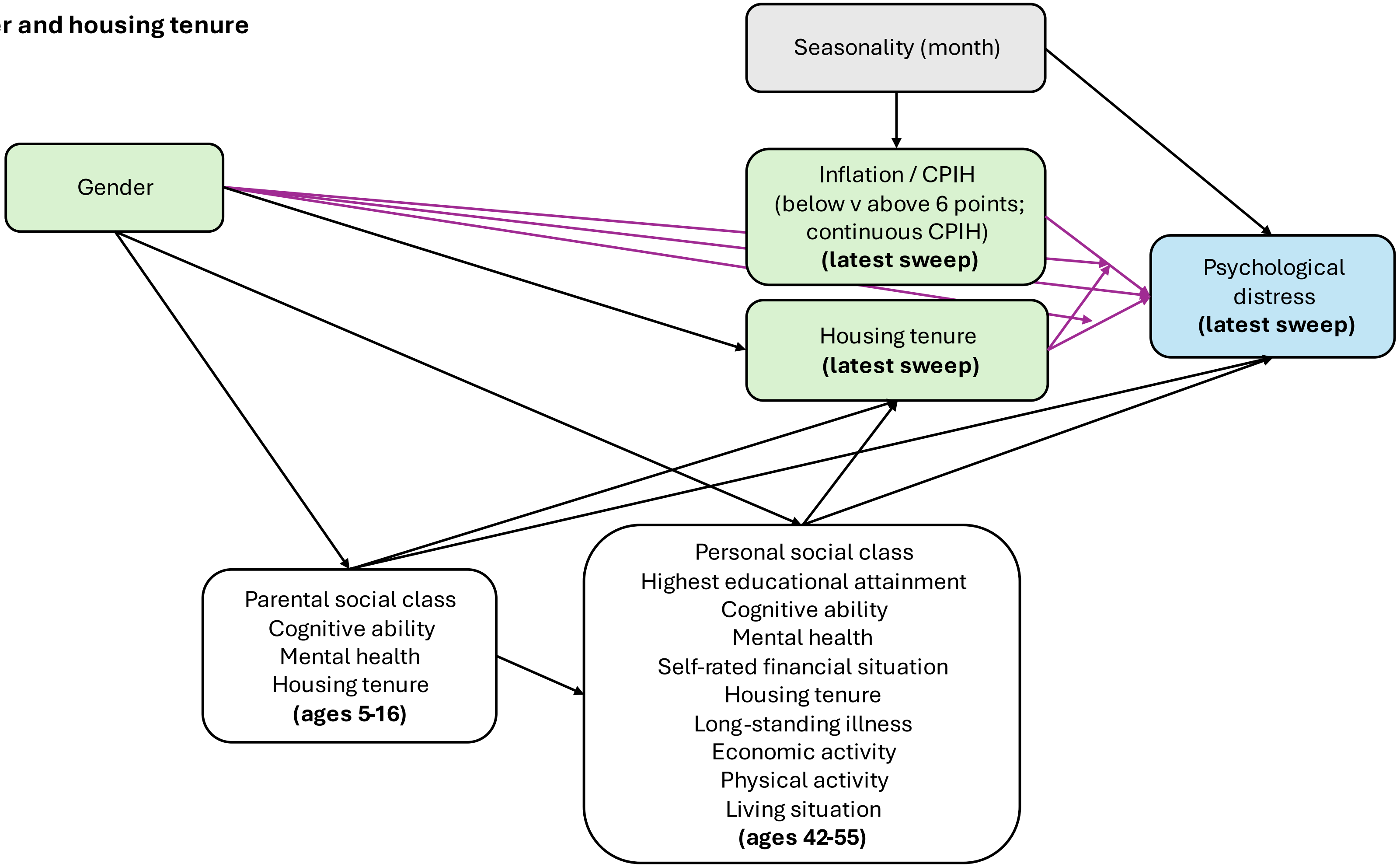

**eAppendix 5. Strengthening the Reporting of Observational Studies in Epidemiology (STROBE) checklist for cohort studies**

| | Item No | Recommendation | Section / paragraph |
|---|---|---|---|
| **Title and abstract** | 1 | (*a*) Indicate the study's design with a commonly used term in the title or the abstract | Title, abstract |
| | | (*b*) Provide in the abstract an informative and balanced summary of what was done and what was found | Abstract |
| **Introduction** | | | |
| Background/rationale | 2 | Explain the scientific background and rationale for the investigation being reported | Introduction / 1-4 |
| Objectives | 3 | State specific objectives, including any prespecified hypotheses | Introduction / 5 |
| **Methods** | | | |
| Study design | 4 | Present key elements of study design early in the paper | Methods / 1 |
| Setting | 5 | Describe the setting, locations, and relevant dates, including periods of recruitment, exposure, follow-up, and data collection | Methods / 1 |
| Participants | 6 | (*a*) Give the eligibility criteria, and the sources and methods of selection of participants. Describe methods of follow-up | Methods / 1 |
| | | (*b*) For matched studies, give matching criteria and number of exposed and unexposed | - |
| Variables | 7 | Clearly define all outcomes, exposures, predictors, potential confounders, and effect modifiers. Give diagnostic criteria, if applicable | Methods / 2-6<br>eAppendices 2-4 |
| Data sources/ measurement | 8* | For each variable of interest, give sources of data and details of methods of assessment (measurement). Describe comparability of assessment methods if there is more than one group | Methods / 2-6<br>eAppendix 1,2 |
| Bias | 9 | Describe any efforts to address potential sources of bias | Methods / 8-10<br>eAppendices 1, 2, 7, 9 |
| Study size | 10 | Explain how the study size was arrived at | Methods / 8<br>eAppendix 2 |
| Quantitative variables | 11 | Explain how quantitative variables were handled in the analyses. If applicable, describe which groupings were chosen and why | Methods / 2-6 |
| Statistical methods | 12 | (*a*) Describe all statistical methods, including those used to control for confounding | Methods / 7-12<br>eAppendices 2-4 |
| | | (*b*) Describe any methods used to examine subgroups and interactions | Methods / 9, 11 |

| | | | |
|---|---|---|---|
| | | | eAppendix 2 |
| | | (*c*) Explain how missing data were addressed | Methods / 13-15 |
| | | (*d*) If applicable, explain how loss to follow-up was addressed | Methods / 13-15 |
| | | (*e*) Describe any sensitivity analyses | Methods / 10 |
| **Results** | | | |
| Participants | 13* | (a) Report numbers of individuals at each stage of study—eg numbers potentially eligible, examined for eligibility, confirmed eligible, included in the study, completing follow-up, and analysed | Results / 1<br>Table 1 |
| | | (b) Give reasons for non-participation at each stage | Results / 1<br>Table 1 |
| | | (c) Consider use of a flow diagram | |
| Descriptive data | 14* | (a) Give characteristics of study participants (eg demographic, clinical, social) and information on exposures and potential confounders | Results / 1<br>Table 1 |
| | | (b) Indicate number of participants with missing data for each variable of interest | Table 1 |
| | | (c) Summarise follow-up time (eg, average and total amount) | Results / 1 |
| Outcome data | 15* | Report numbers of outcome events or summary measures over time | Results / 1<br>Table 1 |
| Main results | 16 | (*a*) Give unadjusted estimates and, if applicable, confounder-adjusted estimates and their precision (eg, 95% confidence interval). Make clear which confounders were adjusted for and why they were included | Results / 4-6, 10-12<br>Table 2<br>eAppendices 3-4, 8, 12 |
| | | (*b*) Report category boundaries when continuous variables were categorized | - |
| | | (*c*) If relevant, consider translating estimates of relative risk into absolute risk for a meaningful time period | - |
| Other analyses | 17 | Report other analyses done—eg analyses of subgroups and interactions, and sensitivity analyses | Results / 8, 13<br>eAppendices 9-13 |
| **Discussion** | | | |
| Key results | 18 | Summarise key results with reference to study objectives | Discussion / 1 |
| Limitations | 19 | Discuss limitations of the study, taking into account sources of potential bias or imprecision. Discuss both direction and magnitude of any potential bias | Discussion / 7 |

| | | | |
|---|---|---|---|
| Interpretation | 20 | Give a cautious overall interpretation of results considering objectives, limitations, multiplicity of analyses, results from similar studies, and other relevant evidence | Discussion / 2-5, 8 |
| Generalisability | 21 | Discuss the generalisability (external validity) of the study results | Discussion / 6-7 |
| **Other information** | | | |
| Funding | 22 | Give the source of funding and the role of the funders for the present study and, if applicable, for the original study on which the present article is based | Funding statement |

*Give information separately for exposed and unexposed groups.

*Note*. eAppendices are included in this Supplementary Material.
An Explanation and Elaboration article discusses each checklist item and gives methodological background and published examples of transparent reporting. The STROBE checklist is best used in conjunction with this article (freely available on the Web sites of PLoS Medicine at http://www.plosmedicine.org/, Annals of Internal Medicine at http://www.annals.org/, and Epidemiology at http://www.epidem.com/). Information on the STROBE Initiative is available at http://www.strobe-statement.org.

### eAppendix 6. Descriptive information on the analytical and overall samples

| | NCDS/58 | | | | BCS/70 | | | |
|---|---|---|---|---|---|---|---|---|
| | **Overall, N = 8,215** | | **Without pre-pandemic cases, N = 6,553** | | **Overall, N = 7,789** | | **Without pre- and during pandemic cases, N = 7,629** | |
| ***Social identities and positions*** | | | | | | | | |
| **Gender** | | | | | | | | |
| Women | 4,127 | 50.2% | 3,277 | 50.0% | 4,100 | 52.6% | 4,015 | 52.6% |
| Men | 4,088 | 49.8% | 3,276 | 50.0% | 3,689 | 47.4% | 3,614 | 47.4% |
| **Childhood socioeconomic position** | | | | | | | | |
| Parental social class | | | | | | | | |
| Manual | 4,211 | 51.3% | 3,394 | 51.8% | 3,694 | 47.4% | 3,618 | 47.4% |
| Non-manual | 2,693 | 32.8% | 2,116 | 32.3% | 2,921 | 37.5% | 2,864 | 37.5% |
| *Missing* | *1,311* | *16.0%* | *1,043* | *15.9%* | *1,174* | *15.1%* | *1,147* | *15.0%* |
| Childhood housing tenure | | | | | | | | |
| Owned at both time points | 2,824 | 34.4% | 2,250 | 34.3% | 3,502 | 45.0% | 3,418 | 44.8% |
| Owned at one time point | 516 | 6.3% | 414 | 6.3% | 572 | 7.3% | 565 | 7.4% |
| Rented at both time points | 3,025 | 36.8% | 2,419 | 36.9% | 1,694 | 21.7% | 1,667 | 21.9% |
| *Missing* | *1,850* | *22.5%* | *1,470* | *22.4%* | *2,021* | *25.9%* | *1,979* | *25.9%* |
| Gender * parental social class | | | | | | | | |
| Women * Manual | 2,104 | 25.6% | 1,699 | 25.9% | 1,939 | 24.9% | 1,898 | 24.9% |
| Women * Non-manual | 1,352 | 16.5% | 1,048 | 16.0% | 1,560 | 20.0% | 1,529 | 20.0% |
| *Women * missing* | *671* | *8.2%* | *530* | *8.1%* | *601* | *7.7%* | *588* | *7.7%* |
| Men * Manual | 2,107 | 25.6% | 1,695 | 25.9% | 1,755 | 22.5% | 1,720 | 22.5% |
| Men * Non-manual | 1,341 | 16.3% | 1,068 | 16.3% | 1,361 | 17.5% | 1,335 | 17.5% |
| *Men * missing* | *640* | *7.8%* | *513* | *7.8%* | *573* | *7.4%* | *559* | *7.3%* |
| Gender * childhood housing tenure | | | | | | | | |

| | | | | | | | | |
|---|---|---|---|---|---|---|---|---|
| Women * owned at both time points | 1,390 | 16.9% | 1,093 | 16.7% | 1,866 | 24.0% | 1,819 | 23.8% |
| Women * owned at one time point | 270 | 3.3% | 218 | 3.3% | 317 | 4.1% | 312 | 4.1% |
| Women * rented at both time points | 1,555 | 18.9% | 1,239 | 18.9% | 896 | 11.5% | 883 | 11.6% |
| *Women * missing* | *912* | *11.1%* | *727* | *11.1%* | *1,021* | *13.1%* | *1,001* | *13.1%* |
| Men * owned at both time points | 1,434 | 17.5% | 1,157 | 17.7% | 1,636 | 21.0% | 1,599 | 21.0% |
| Men * owned at one time point | 246 | 3.0% | 196 | 3.0% | 255 | 3.3% | 253 | 3.3% |
| Men * rented at both time points | 1,470 | 17.9% | 1,180 | 18.0% | 798 | 10.2% | 784 | 10.3% |
| *Men * missing* | *938* | *11.4%* | *743* | *11.3%* | *1,000* | *12.8%* | *978* | *12.8%* |
| **Adult socioeconomic position** | | | | | | | | |
| Adult housing tenure | | | | | | | | |
| Own outright | 5,586 | 68.0% | 4,556 | 69.5% | 2,081 | 26.7% | 2,036 | 26.7% |
| Own with mortgage | 1,247 | 15.2% | 942 | 14.4% | 4,152 | 53.3% | 4,062 | 53.2% |
| Other | 1,222 | 14.9% | 977 | 14.9% | 1,516 | 19.5% | 1,491 | 19.5% |
| *Missing* | *160* | *1.9%* | *78* | *1.2%* | *40* | *0.5%* | *40* | *0.5%* |
| Self-rated financial situation | | | | | | | | |
| Just about/quite/very difficult | 1,209 | 14.7% | 973 | 14.8% | 1,549 | 19.9% | 1,519 | 19.9% |
| Doing all right | 3,090 | 37.6% | 2,481 | 37.9% | 3,330 | 42.8% | 3,264 | 42.8% |
| Living comfortably | 3,870 | 47.1% | 3,059 | 46.7% | 2,868 | 36.8% | 2,804 | 36.8% |
| *Missing* | *46* | *0.6%* | *40* | *0.6%* | *42* | *0.5%* | *42* | *0.6%* |
| Gender * adult housing tenure | | | | | | | | |
| Women * own outright | 2,892 | 35.2% | 2,346 | 35.8% | 1,171 | 15.0% | 1,144 | 15.0% |
| Women * own with mortgage | 561 | 6.8% | 423 | 6.5% | 2,118 | 27.2% | 2,073 | 27.2% |
| Women * other | 605 | 7.4% | 475 | 7.2% | 789 | 10.1% | 776 | 10.2% |
| *Women * missing* | *69* | *0.8%* | *33* | *0.5%* | *22* | *0.3%* | *22* | *0.3%* |

| | | | | | | | | | |
|---|---|---|---|---|---|---|---|---|---|
| Men * own outright | 2,694 | 32.8% | 2,210 | 33.7% | | 910 | 11.7% | 892 | 11.7% |
| Men * own with mortgage | 686 | 8.4% | 519 | 7.9% | | 2,034 | 26.1% | 1,989 | 26.1% |
| Men * other | 617 | 7.5% | 502 | 7.7% | | 727 | 9.3% | 715 | 9.4% |
| *Men * missing* | *91* | *1.1%* | *45* | *0.7%* | | *18* | *0.2%* | *18* | *0.2%* |
| Gender * self-rated financial situation | | | | | | | | | |
| Women * just about/quite/very difficult | 622 | 7.6% | 499 | 7.6% | | 871 | 11.2% | 854 | 11.2% |
| Women * doing all right | 1,506 | 18.3% | 1,218 | 18.6% | | 1,685 | 21.6% | 1,652 | 21.7% |
| Women * living comfortably | 1,972 | 24.0% | 1,537 | 23.5% | | 1,509 | 19.4% | 1,474 | 19.3% |
| *Women * missing* | *27* | *0.3%* | *23* | *0.4%* | | *35* | *0.4%* | *35* | *0.5%* |
| Men * just about/quite/very difficult | 587 | 7.1% | 474 | 7.2% | | 678 | 8.7% | 665 | 8.7% |
| Men * doing all right | 1,584 | 19.3% | 1,263 | 19.3% | | 1,645 | 21.1% | 1,612 | 21.1% |
| Men * living comfortably | 1,898 | 23.1% | 1,522 | 23.2% | | 1,359 | 17.4% | 1,330 | 17.4% |
| *Men * missing* | *19* | *0.2%* | *17* | *0.3%* | | *7* | *0.1%* | *7* | *0.1%* |
| **Malaise inventory, M (SD)** | | | | | | | | | |
| *Age 23* | 1.12 | 1.46 | 1.11 | 1.45 | *Age 26* | 1.69 | 1.72 | 1.69 | 1.71 |
| *Age 33* | 0.92 | 1.44 | 0.90 | 1.42 | *Age 29* | 1.45 | 1.67 | 1.45 | 1.66 |
| *Age 42* | 1.43 | 1.68 | 1.42 | 1.68 | *Age 34* | 1.59 | 1.84 | 1.59 | 1.84 |
| *Age 50* | 1.40 | 1.85 | 1.40 | 1.87 | *Age 42* | 1.77 | 1.93 | 1.77 | 1.93 |
| *Age 62* | 1.22 | 1.69 | 1.22 | 1.69 | *Age 46* | 1.70 | 2.06 | 1.69 | 2.06 |
| *Age 62.5* | 1.50 | 1.88 | 1.50 | 1.88 | *Age 50* | 1.63 | 1.93 | 1.63 | 1.93 |
| *Age 63* | 1.43 | 1.84 | 1.43 | 1.85 | *Age 50.5* | 1.97 | 2.10 | 1.97 | 2.10 |
| *Age 64.5* | 1.40 | 1.80 | 1.38 | 1.78 | *Age 51* | 1.86 | 2.06 | 1.86 | 2.06 |
| | | | | | *Age 52.5* | 1.66 | 2.00 | 1.66 | 2.00 |
| **Malaise inventory missingness, N (%)** | | | | | | | | | |
| *Age 23* | *856* | *11.0%* | *624* | *10.2%* | *Age 26* | *2,562* | *32.0%* | *2,512* | *32.0%* |
| *Age 33* | *685* | *8.8%* | *478* | *7.8%* | *Age 29* | *1,273* | *15.9%* | *1,252* | *15.9%* |

| | | | | | | | | | |
|---|---|---|---|---|---|---|---|---|---|
| *Age 42* | *268* | *3.4%* | *142* | *2.3%* | *Age 34* | *1,666* | *20.8%* | *1,632* | *20.8%* |
| *Age 50* | *431* | *5.5%* | *272* | *4.4%* | *Age 42* | *1,718* | *21.4%* | *1,696* | *21.6%* |
| *Age 62* | *3,540* | *45.4%* | *2,783* | *45.3%* | *Age 46* | *1,619* | *20.2%* | *1,596* | *20.3%* |
| *Age 62.5* | *2,372* | *30.4%* | *1,797* | *29.3%* | *Age 50* | *4,589* | *57.2%* | *4,511* | *57.4%* |
| *Age 63* | *1,957* | *25.1%* | *1,463* | *23.8%* | *Age 50.5* | *3,613* | *45.1%* | *3,507* | *44.6%* |
| *Age 64.5* | *68* | *0.9%* | *22* | *0.4%* | *Age 51* | *3,215* | *40.1%* | *3,143* | *40.0%* |
| | | | | | *Age 52.5* | *828* | *10.3%* | *819* | *10.4%* |

*Note.* BCS/70: 1970 British Cohort Study; M: mean; N: frequency; NCDS/58: 1958 National Child Development Study; SD: standard deviation; SEP: socioeconomic position. Results based on unweighted data. Analytical samples exclude participants who took part in the latest main survey sweep prior to the COVID-19 pandemic onset (n=1,662 in NCDS/58 and n=116 in BCS/70) or within the period spanning the COVID-19 Surveys data collection (n=44 in BCS/70, interviewed between September-October 2020).

**eAppendix 7. Selection of optimal functional form for multilevel growth curve models**

**Visualisation of imputed and weighted life-course data**

The graphs below show the connected 95% confidence intervals (CI) of the mean Malaise Inventory score estimated independently at each time-point, based on weighted and imputed data, for the overall samples, by gender, each of the childhood SEP indicators and, finally, by both gender and childhood SEP. The different 95% CIs have been connected for visualisation purposes only.

***Overall***

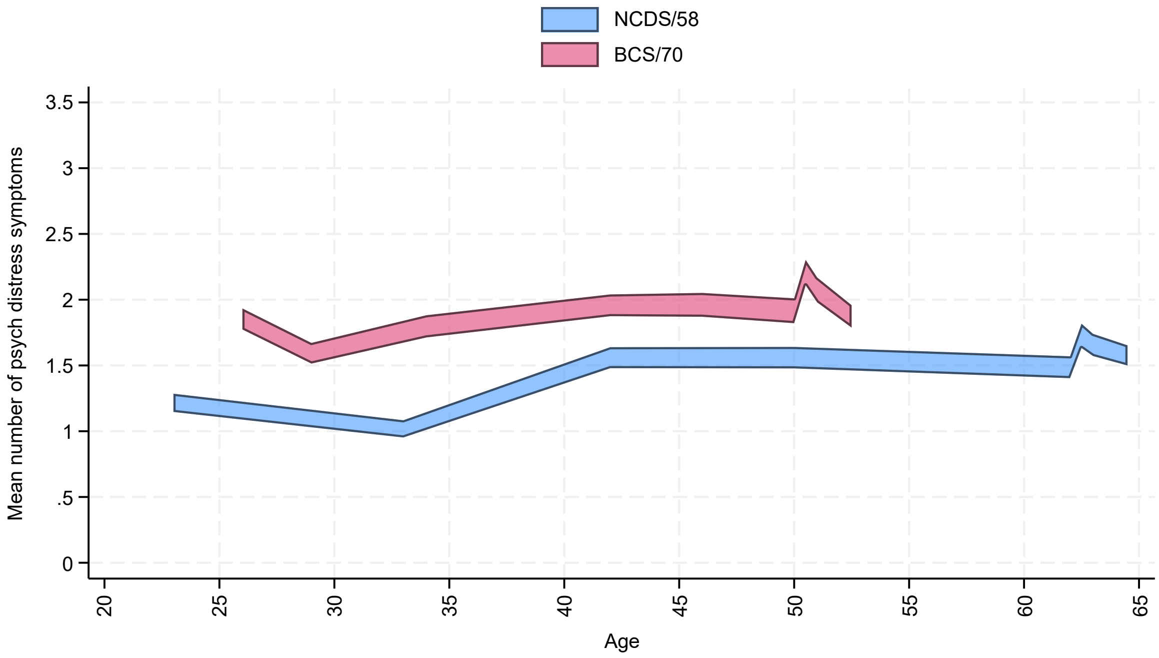

***By gender***

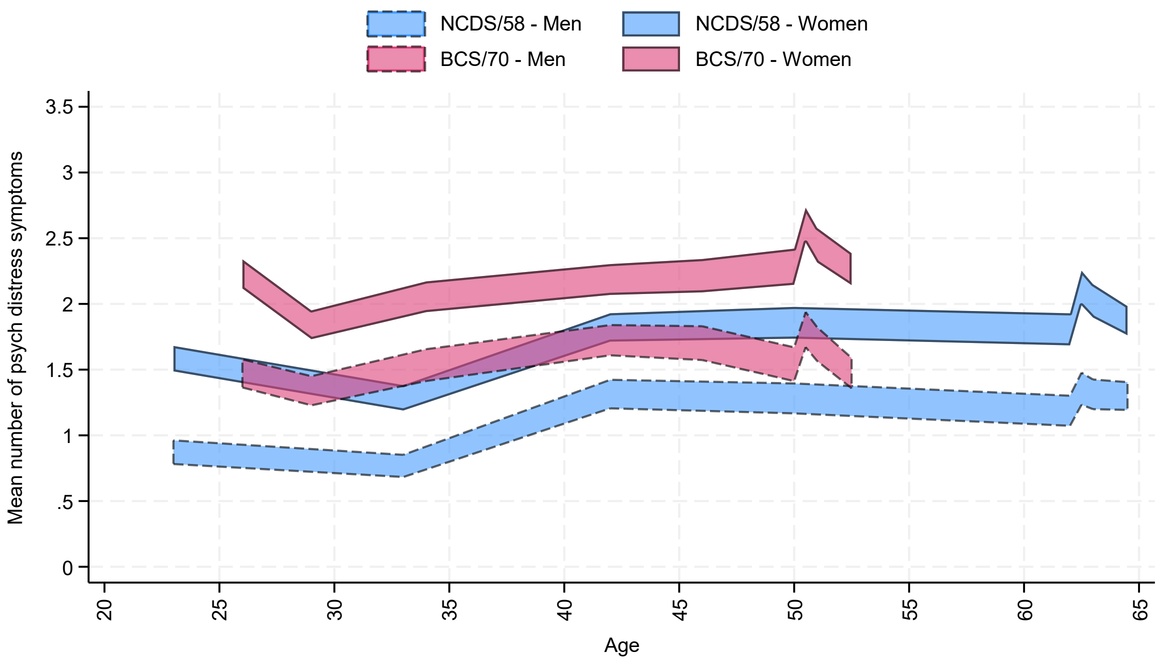

***By parental social class***

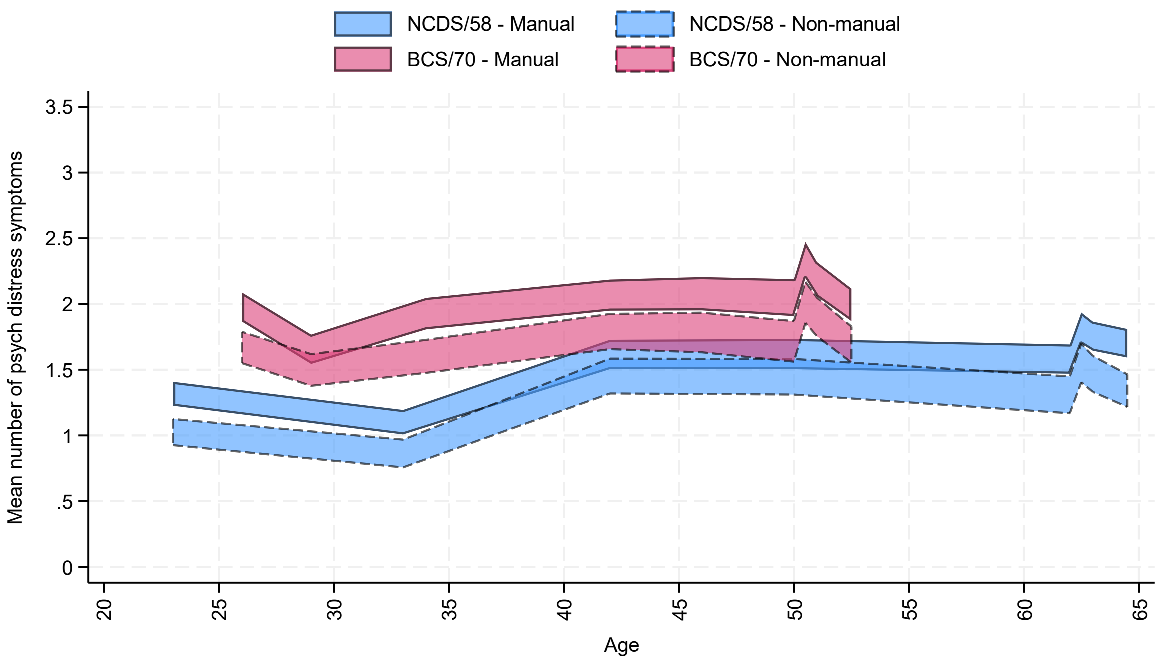

***By childhood housing tenure***

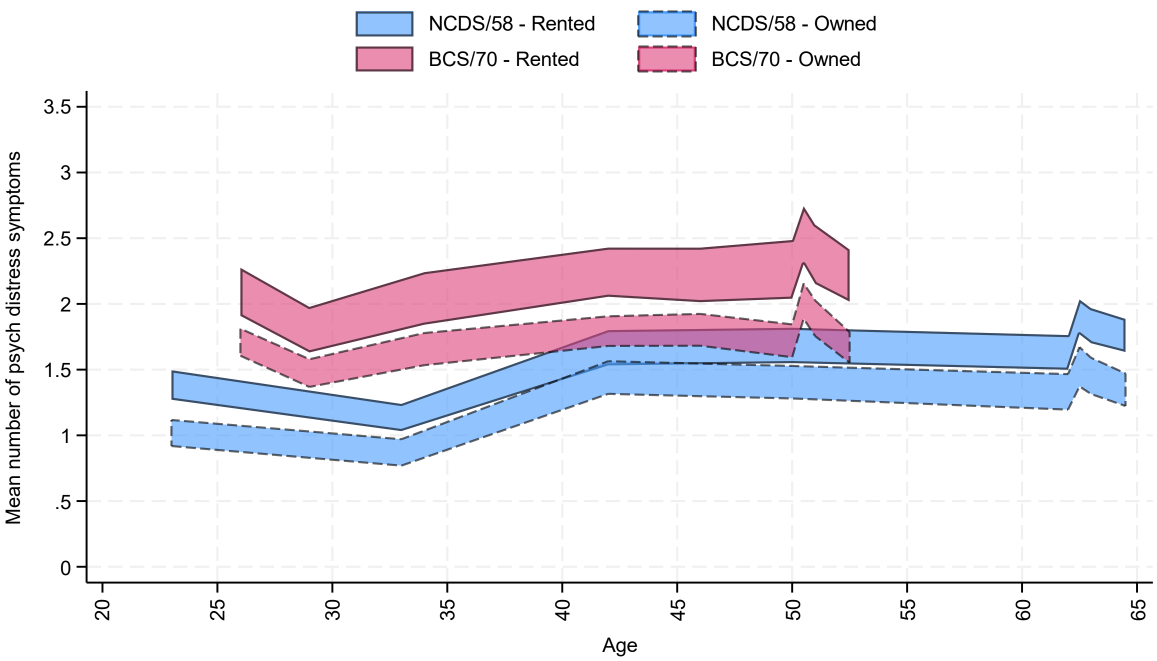

***By gender (M: men; W: women) and parental social class (M: manual; NM: non-manual)***

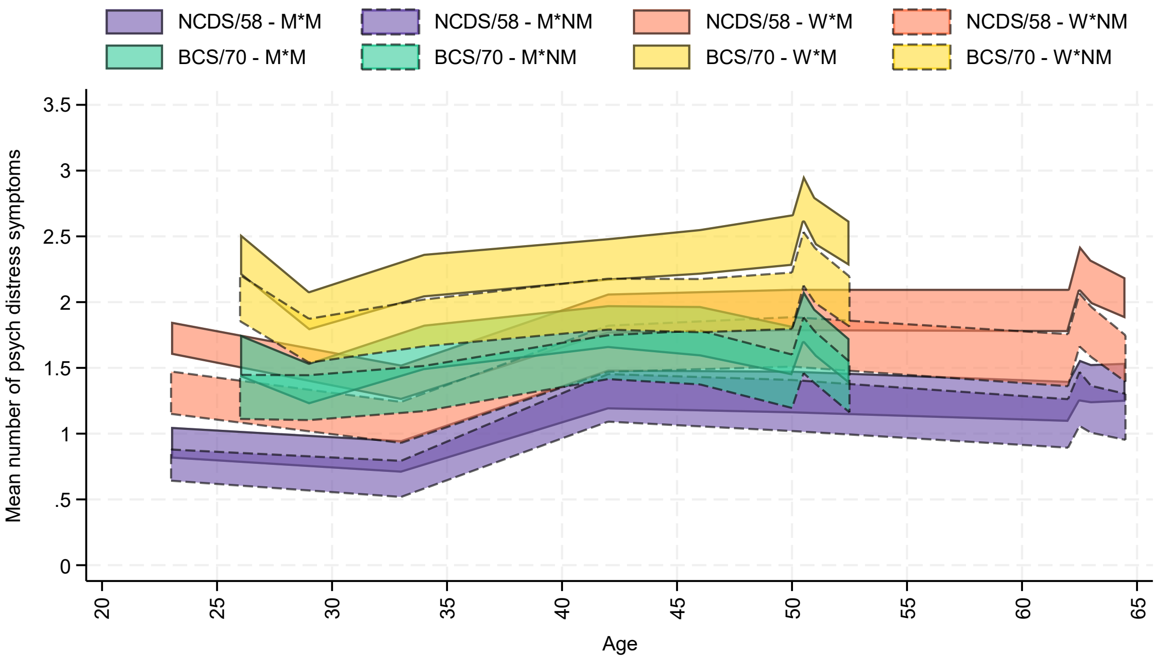

***By gender (M: men; W: women) and childhood housing tenure***

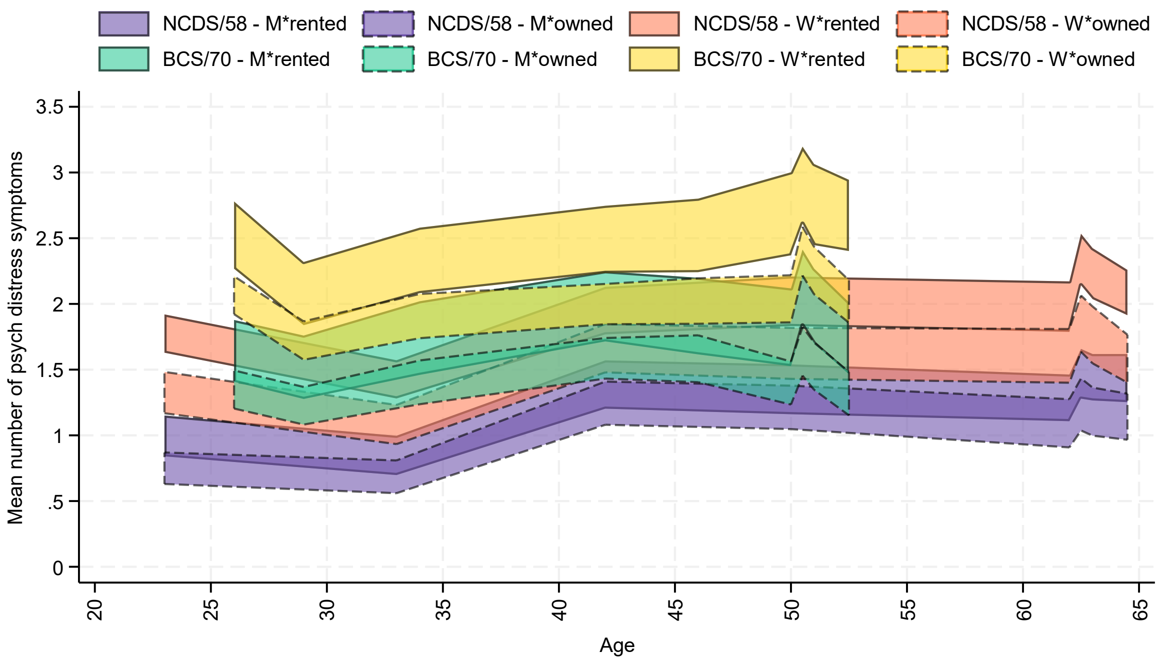

### Candidate model specifications and model comparison strategy

The visual exploration of the weighted and imputed mean values of the outcome measure across the different time-points in both birth cohorts suggested the appropriateness of exploring three main functional forms for the long-term trajectories:

1. A cubic polynomial trajectory, which would allow to model the initial decrease, later increase, and further plateau or reversal of the increase in both cohorts, although without much ability to capture the observed increases during the COVID-19 pandemic.
2. A cubic polynomial trajectory with a time-specific factor capturing the step-increase during the COVID-19 pandemic, which would allow to, in addition to the previous case (#1), accommodate a transient increase during that period).
3. A piecewise model with a cubic and a quadratic spline with a knot at the latest pre-pandemic assessment, which, unlike #2, would allow to model the changes during and after the pandemic with more granularity than as a step-increase, but rather as an initial increase and posterior decrease (without necessarily returning to the overall cubic trajectory).

Models were estimated using a complete-case approach, separately for each birth cohort and for each condition (overall and then including an interaction between the growth parameters and gender, each of the childhood socioeconomic position indicators, and the gender*childhood SEP intersection). Models were estimated with and without random slopes for the linear growth parameters (linear random slopes were chosen to accommodate variability in the change over time across individuals without introducing the model complexity that random slopes for the polynomial terms would have introduced). Random intercepts were included as default to accommodate individual variability in the starting points.

The Akaike Information Criterion (AIC) and Bayesian Information Criterion (BIC) were obtained, representing the ability of the model to reproduce the data, with lower AIC and BIC suggesting better fit. Models with the best fit to the data were selected. Where different models were supported by the AIC and BIC indices, models with more consistent functional forms across the different scenarios were favoured.

Results of the model comparison strategy can be found in the table below.

### Model comparison results

| Interaction with growth parameters | Model | NCDS/58 | | | | | BCS/70 | | | | |
|---|---|---|---|---|---|---|---|---|---|---|---|
| | | N obs | Log-likelihood | df | AIC | BIC | N obs | Log-likelihood | df | AIC | BIC |
| Overall (none) | Cubic | 41,539 | -59,309.4 | 6 | 118,630.7 | 118,682.5 | 50,036 | -74,486.1 | 6 | 148,984.1 | 149,037.1 |
| | Cubic with random linear slopes | 41,539 | -58,219.8 | 7 | 116,453.6 | 116,514.0 | 50,036 | -73,469.4 | 7 | 146,952.7 | 147,014.5 |
| | Cubic with pandemic factor | 41,539 | -59,308.1 | 7 | 118,630.2 | 118,690.7 | 50,036 | -74,463.5 | 7 | 148,941.0 | 149,002.8 |
| | Cubic with pandemic factor and random linear slopes | 41,539 | -58,219.5 | 8 | 116,454.9 | 116,524.0 | 50,036 | -73,431.7 | 8 | 146,879.4 | 146,950.0 |
| | Piecewise cubic + quadratic | 41,539 | -59,286.5 | 8 | 118,589.1 | 118,658.1 | 50,036 | -74,448.8 | 8 | 148,913.6 | 148,984.1 |
| | Piecewise cubic + quadratic with random linear slopes | 41,539 | -58,167.0 | 10 | **116,353.9** | **116,440.2** | 50,036 | -73,325.7 | 10 | **146,671.4** | **146,759.6** |
| Gender | Cubic | 41,539 | -59,118.2 | 10 | 118,256.4 | 118,342.8 | 50,036 | -74,301.5 | 10 | 148,623.0 | 148,711.2 |
| | Cubic with random linear slopes | 41,539 | -58,019.6 | 11 | 116,061.2 | 116,156.2 | 50,036 | -73,290.8 | 11 | 146,603.5 | 146,700.6 |
| | Cubic with pandemic factor | 41,539 | -59,114.3 | 12 | 118,252.6 | 118,356.2 | 50,036 | -74,282.8 | 12 | 148,589.6 | 148,695.4 |
| | Cubic with pandemic factor and random linear slopes | 41,539 | -58,016.6 | 13 | 116,059.2 | 116,171.4 | 50,036 | -73,254.9 | 13 | 146,535.8 | 146,650.5 |
| | Piecewise cubic + quadratic | 41,539 | -59,091.2 | 14 | 118,210.5 | 118,331.3 | 50,036 | -74,267.0 | 14 | 148,562.0 | 148,685.5 |
| | Piecewise cubic + quadratic with random linear slopes | 41,539 | -57,961.3 | 16 | **115,954.7** | **116,092.8** | 50,036 | -73,145.9 | 16 | **146,323.8** | **146,465.0** |
| Parental social class | Cubic | 35,377 | -48,541.4 | 10 | 97,102.7 | 97,187.5 | 43,087 | -61,527.2 | 10 | 123,074.5 | 123,161.2 |
| | Cubic with random linear slopes | 35,377 | -47,763.2 | 11 | 95,548.3 | 95,641.5 | 43,087 | -60,819.3 | 11 | 121,660.5 | 121,755.9 |
| | Cubic with pandemic factor | 35,377 | -48,540.6 | 12 | 97,105.1 | 97,206.8 | 43,087 | -61,502.4 | 12 | 123,028.7 | 123,132.8 |
| | Cubic with pandemic factor and random linear slopes | 35,377 | -47,762.8 | 13 | 95,551.7 | 95,661.8 | 43,087 | -60,785.3 | 13 | 121,596.5 | 121,709.3 |
| | Piecewise cubic + quadratic | 35,377 | -48,519.7 | 14 | 97,067.3 | 97,186.0 | 43,087 | -61,489.5 | 14 | 123,006.9 | 123,128.3 |
| | Piecewise cubic + quadratic with random linear slopes | 35,377 | -47,736.2 | 16 | **95,504.3** | **95,639.9** | 43,087 | -60,745.3 | 16 | **121,522.6** | **121,661.3** |
| | Cubic | 32,723 | -44,461.7 | 14 | 88,951.4 | 89,068.9 | 37,716 | -52,003.9 | 14 | 104,035.8 | 104,155.3 |

| | | | | | | | | | | | |
|---|---|---|---|---|---|---|---|---|---|---|---|
| **Childhood housing tenure** | Cubic with random linear slopes | 32,723 | -43,792.5 | 15 | 87,615.0 | **87,740.9** | 37,716 | -51,470.7 | 15 | 102,971.5 | 103,099.5 |
| | Cubic with pandemic factor | 32,723 | -44,459.8 | 17 | 88,953.6 | 89,096.3 | 37,716 | -51,982.2 | 17 | 103,998.5 | 104,143.6 |
| | Cubic with pandemic factor and random linear slopes | 32,723 | -43,790.5 | 18 | 87,616.9 | 87,768.1 | 37,716 | -51,442.5 | 18 | 102,921.1 | **103,074.7** |
| | Piecewise cubic + quadratic | 32,723 | -44,442.0 | 20 | 88,924.0 | 89,091.9 | 37,716 | -51,969.2 | 20 | 103,978.3 | 104,149.1 |
| | Piecewise cubic + quadratic with random linear slopes | 32,723 | -43,768.1 | 22 | **87,580.2** | 87,764.9 | 37,716 | -51,423.2 | 22 | **102,890.3** | 103,078.2 |
| **Gender and parental social class** | Cubic | 35,377 | -48,367.5 | 18 | 96,771.0 | 96,923.5 | 43,087 | -61,354.0 | 18 | 122,743.9 | 122,900.0 |
| | Cubic with random linear slopes | 35,377 | -47,579.4 | 19 | 95,196.7 | **95,357.7** | 43,087 | -60,653.0 | 19 | 121,343.9 | 121,508.6 |
| | Cubic with pandemic factor | 35,377 | -48,359.8 | 22 | 96,763.6 | 96,950.0 | 43,087 | -61,332.7 | 22 | 122,709.5 | 122,900.2 |
| | Cubic with pandemic factor and random linear slopes | 35,377 | -47,572.6 | 23 | 95,191.2 | 95,386.1 | 43,087 | -60,620.4 | 23 | 121,286.8 | 121,486.3 |
| | Piecewise cubic + quadratic | 35,377 | -48,337.2 | 26 | 96,726.3 | 96,946.6 | 43,087 | -61,318.3 | 26 | 122,688.6 | 122,914.1 |
| | Piecewise cubic + quadratic with random linear slopes | 35,377 | -47,544.0 | 28 | **95,144.0** | 95,381.3 | 43,087 | -60,578.6 | 28 | **121,213.1** | **121,455.9** |
| **Gender and childhood housing tenure** | Cubic | 32,723 | -44,293.6 | 26 | 88,639.1 | 88,857.4 | 37,716 | -51,851.7 | 26 | 103,755.4 | 103,977.4 |
| | Cubic with random linear slopes | 32,723 | -43,618.3 | 27 | 87,290.7 | **87,517.4** | 37,716 | -51,318.4 | 27 | 102,690.9 | **102,921.4** |
| | Cubic with pandemic factor | 32,723 | -44,289.0 | 32 | 88,642.0 | 88,910.7 | 37,716 | -51,831.8 | 32 | 103,727.5 | 104,000.7 |
| | Cubic with pandemic factor and random linear slopes | 32,723 | -43,613.4 | 33 | 87,292.8 | 87,569.8 | 37,716 | -51,290.2 | 33 | 102,646.4 | 102,928.2 |
| | Piecewise cubic + quadratic | 32,723 | -44,270.2 | 38 | 88,616.4 | 88,935.4 | 37,716 | -51,816.9 | 38 | 103,709.7 | 104,034.2 |
| | Piecewise cubic + quadratic with random linear slopes | 32,723 | -43,589.7 | 40 | **87,259.4** | 87,595.2 | 37,716 | -51,268.4 | 40 | **102,616.8** | 102,958.3 |

*Note.* AIC: Akaike Information Criterion; BCS/70: 1970 British Cohort Study; BIC: Bayesian Information Criterion; df: degrees of freedom; NCDS/58: 1958 National Child Development Study. Best fit indices within each condition are highlighted in boldface.

### eAppendix 8. Results from the multilevel growth curve models

| | NCDS/58 (n=6,553) | | BCS/70 (n=7,629) | |
|---|---|---|---|---|
| | B (95% CI) | p | B (95% CI) | p |
| **Overall** | | | | |
| Intercept | 1.11 (1.06, 1.16) | <0.001 | 1.68 (1.62, 1.73) | <0.001 |
| Spline 1, linear term | -0.05 (-0.06, -0.03) | <0.001 | -0.05 (-0.08, -0.03) | <0.001 |
| Spline 1, quadratic term | 0.004 (0.004, 0.005) | <0.001 | 0.006 (0.004, 0.009) | <0.001 |
| Spline 1, cubic term | -0.0001 (-0.0001, -0.0001) | <0.001 | -0.0002 (-0.0002, -0.0001) | <0.001 |
| Spline 2, linear term | 0.33 (0.21, 0.44) | <0.001 | 0.37 (0.24, 0.50) | <0.001 |
| Spline 2, quadratic term | -0.12 (-0.17, -0.08) | <0.001 | -0.16 (-0.21, -0.11) | <0.001 |
| Spline 1, linear term (SD) | 0.03 (0.03, 0.03) | | 0.05 (0.05, 0.06) | |
| Spline 2, linear term (SD) | 0.23 (0.20, 0.27) | | 0.27 (0.24, 0.30) | |
| Intercept (SD) | 1.02 (0.98, 1.07) | | 1.28 (1.24, 1.32) | |
| Residual (SD) | 1.09 (1.06, 1.12) | | 1.19 (1.16, 1.21) | |
| **Gender (ref.: Men)** | | | | |
| Intercept | 0.75 (0.69, 0.82) | <0.001 | 1.30 (1.23, 1.38) | <0.001 |
| Spline 1, linear term | -0.03 (-0.05, -0.01) | 0.004 | -0.03 (-0.06, 0.01) | 0.114 |
| Spline 1, quadratic term | 0.004 (0.002, 0.005) | <0.001 | 0.006 (0.002, 0.009) | 0.002 |
| Spline 1, cubic term | -0.0001 (-0.0001, 0.0000) | <0.001 | -0.0002 (-0.0003, -0.0001) | <0.001 |
| Spline 2, linear term | 0.23 (0.08, 0.39) | 0.004 | 0.35 (0.19, 0.51) | <0.001 |
| Spline 2, quadratic term | -0.08 (-0.14, -0.02) | 0.006 | -0.16 (-0.22, -0.10) | <0.001 |
| Intercept * Women | 0.72 (0.62, 0.82) | <0.001 | 0.73 (0.62, 0.83) | <0.001 |
| Spline 1, linear term * Women | -0.04 (-0.07, -0.01) | 0.020 | -0.05 (-0.10, 0.00) | 0.069 |
| Spline 1, quadratic term * Women | 0.002 (0.000, 0.004) | 0.126 | 0.002 (-0.004, 0.007) | 0.568 |
| Spline 1, cubic term * Women | 0.0000 (-0.0001, 0.0000) | 0.295 | 0.0000 (-0.0001, 0.0002) | 0.810 |
| Spline 2, linear term * Women | 0.20 (-0.05, 0.44) | 0.111 | 0.04 (-0.20, 0.28) | 0.747 |
| Spline 2, quadratic term * Women | -0.09 (-0.18, 0.00) | 0.052 | -0.01 (-0.10, 0.08) | 0.828 |
| Spline 1, linear term (SD) | 0.03 (0.03, 0.03) | | 0.05 (0.05, 0.05) | |
| Spline 2, linear term (SD) | 0.23 (0.20, 0.27) | | 0.27 (0.23, 0.30) | |

| | | | | |
|---|---|---|---|---|
| Intercept (SD) | 0.98 (0.94, 1.03) | | 1.25 (1.21, 1.29) | |
| Residual (SD) | 1.09 (1.06, 1.12) | | 1.19 (1.16, 1.21) | |
| **Parental social class (ref.: Manual)** | | | | |
| Intercept | 1.20 (1.13, 1.27) | <0.001 | 1.78 (1.70, 1.86) | <0.001 |
| Spline 1, linear term | -0.05 (-0.07, -0.03) | <0.001 | -0.05 (-0.09, -0.02) | 0.002 |
| Spline 1, quadratic term | 0.004 (0.003, 0.006) | <0.001 | 0.007 (0.003, 0.010) | <0.001 |
| Spline 1, cubic term | -0.0001 (-0.0001, -0.0001) | <0.001 | -0.0002 (-0.0003, -0.0001) | <0.001 |
| Spline 2, linear term | 0.33 (0.18, 0.48) | <0.001 | 0.35 (0.17, 0.53) | <0.001 |
| Spline 2, quadratic term | -0.12 (-0.18, -0.06) | <0.001 | -0.16 (-0.22, -0.09) | <0.001 |
| Intercept * Non-manual | -0.24 (-0.35, -0.14) | <0.001 | -0.23 (-0.35, -0.12) | <0.001 |
| Spline 1, linear term * Non-manual | 0.00 (-0.03, 0.04) | 0.808 | 0.01 (-0.04, 0.05) | 0.789 |
| Spline 1, quadratic term * Non-manual | 0.000 (-0.002, 0.003) | 0.742 | -0.001 (-0.005, 0.004) | 0.798 |
| Spline 1, cubic term * Non-manual | 0.0000 (0.0000, 0.0000) | 0.557 | 0.0000 (-0.0001, 0.0001) | 0.857 |
| Spline 2, linear term * Non-manual | -0.01 (-0.25, 0.23) | 0.946 | 0.05 (-0.21, 0.30) | 0.709 |
| Spline 2, quadratic term * Non-manual | -0.01 (-0.10, 0.08) | 0.823 | -0.02 (-0.11, 0.08) | 0.749 |
| Spline 1, linear term (SD) | 0.03 (0.03, 0.03) | | 0.05 (0.05, 0.06) | |
| Spline 2, linear term (SD) | 0.23 (0.20, 0.27) | | 0.27 (0.24, 0.30) | |
| Intercept (SD) | 1.02 (0.98, 1.06) | | 1.27 (1.24, 1.31) | |
| Residual (SD) | 1.09 (1.06, 1.12) | | 1.19 (1.16, 1.21) | |
| **Childhood housing tenure (ref.: Rented at both time-points)** | | | | |
| Intercept | 1.27 (1.19, 1.35) | <0.001 | 1.87 (1.75, 1.99) | <0.001 |
| Spline 1, linear term | -0.06 (-0.08, -0.03) | <0.001 | -0.05 (-0.10, 0.00) | 0.070 |
| Spline 1, quadratic term | 0.005 (0.003, 0.007) | <0.001 | 0.006 (0.001, 0.011) | 0.024 |
| Spline 1, cubic term | -0.0001 (-0.0001, -0.0001) | <0.001 | -0.0002 (-0.0003, 0.0000) | 0.031 |
| Spline 2, linear term | 0.38 (0.20, 0.56) | <0.001 | 0.31 (0.03, 0.59) | 0.031 |
| Spline 2, quadratic term | -0.14 (-0.21, -0.07) | <0.001 | -0.14 (-0.24, -0.04) | 0.008 |
| Intercept * Owned at one time-point | -0.16 (-0.37, 0.05) | 0.128 | -0.13 (-0.36, 0.10) | 0.265 |
| Intercept * Owned at both time-points | -0.34 (-0.46, -0.22) | <0.001 | -0.30 (-0.45, -0.15) | <0.001 |
| Spline 1, linear term * Owned at one time-point | 0.01 (-0.05, 0.07) | 0.747 | -0.02 (-0.13, 0.08) | 0.670 |
| Spline 1, linear term * Owned at both time-points | 0.02 (-0.01, 0.06) | 0.241 | 0.00 (-0.07, 0.07) | 0.981 |

| | | | | |
|---|---|---|---|---|
| Spline 1, quadratic term * Owned at one time-point | 0.000 (-0.005, 0.004) | 0.843 | 0.002 (-0.008, 0.013) | 0.638 |
| Spline 1, quadratic term * Owned at both time-points | -0.001 (-0.003, 0.002) | 0.491 | 0.000 (-0.006, 0.007) | 0.957 |
| Spline 1, cubic term * Owned at one time-point | 0.0000 (-0.0001, 0.0001) | 0.931 | -0.0001 (-0.0003, 0.0002) | 0.599 |
| Spline 1, cubic term * Owned at both time-points | 0.0000 (0.0000, 0.0001) | 0.674 | 0.0000 (-0.0002, 0.0002) | 0.859 |
| Spline 2, linear term * Owned at one time-point | -0.05 (-0.48, 0.38) | 0.814 | 0.14 (-0.34, 0.63) | 0.566 |
| Spline 2, linear term * Owned at both time-points | -0.12 (-0.38, 0.14) | 0.373 | 0.08 (-0.24, 0.40) | 0.613 |
| Spline 2, quadratic term * Owned at one time-point | 0.03 (-0.13, 0.19) | 0.725 | -0.05 (-0.23, 0.13) | 0.602 |
| Spline 2, quadratic term * Owned at both time-points | 0.03 (-0.06, 0.13) | 0.486 | -0.03 (-0.15, 0.09) | 0.594 |
| Spline 1, linear term (SD) | 0.03 (0.03, 0.03) | | 0.05 (0.05, 0.06) | |
| Spline 2, linear term (SD) | 0.23 (0.20, 0.27) | | 0.27 (0.24, 0.30) | |
| Intercept (SD) | 1.02 (0.97, 1.06) | | 1.27 (1.23, 1.31) | |
| Residual (SD) | 1.09 (1.06, 1.12) | | 1.19 (1.16, 1.21) | |
| **Gender (ref.: Men) and parental social class (ref.: Manual)** | | | | |
| Intercept | 0.80 (0.72, 0.89) | <0.001 | 1.41 (1.30, 1.52) | <0.001 |
| Spline 1, linear term | -0.03 (-0.06, 0.00) | 0.028 | -0.03 (-0.08, 0.02) | 0.206 |
| Spline 1, quadratic term | 0.003 (0.002, 0.005) | <0.001 | 0.006 (0.001, 0.011) | 0.021 |
| Spline 1, cubic term | -0.0001 (-0.0001, 0.0000) | <0.001 | -0.0002 (-0.0003, -0.0001) | 0.008 |
| Spline 2, linear term | 0.23 (0.02, 0.45) | 0.033 | 0.34 (0.11, 0.57) | 0.004 |
| Spline 2, quadratic term | -0.07 (-0.15, 0.00) | 0.064 | -0.15 (-0.24, -0.07) | 0.001 |
| Intercept * Women | 0.80 (0.66, 0.93) | <0.001 | 0.73 (0.57, 0.89) | <0.001 |
| Spline 1, linear term * Women | -0.04 (-0.08, 0.00) | 0.050 | -0.05 (-0.12, 0.02) | 0.204 |
| Spline 1, quadratic term * Women | 0.002 (-0.001, 0.005) | 0.202 | 0.001 (-0.006, 0.008) | 0.694 |
| Spline 1, cubic term * Women | 0.0000 (-0.0001, 0.0000) | 0.398 | 0.0000 (-0.0002, 0.0002) | 0.805 |
| Spline 2, linear term * Women | 0.20 (-0.13, 0.53) | 0.226 | 0.02 (-0.31, 0.36) | 0.901 |
| Spline 2, quadratic term * Women | -0.10 (-0.22, 0.02) | 0.118 | 0.00 (-0.13, 0.12) | 0.952 |
| Intercept * Non-manual | -0.13 (-0.27, 0.00) | 0.052 | -0.24 (-0.41, -0.07) | 0.005 |
| Spline 1, linear term * Non-manual | 0.00 (-0.04, 0.04) | 0.915 | 0.01 (-0.06, 0.08) | 0.781 |
| Spline 1, quadratic term * Non-manual | 0.001 (-0.002, 0.003) | 0.687 | -0.001 (-0.008, 0.007) | 0.831 |
| Spline 1, cubic term * Non-manual | 0.0000 (-0.0001, 0.0000) | 0.613 | 0.0000 (-0.0002, 0.0002) | 0.840 |
| Spline 2, linear term * Non-manual | 0.00 (-0.33, 0.33) | 0.989 | 0.03 (-0.30, 0.35) | 0.872 |

| | | | | |
|---|---|---|---|---|
| Spline 2, quadratic term * Non-manual | -0.02 (-0.14, 0.10) | 0.759 | -0.01 (-0.13, 0.11) | 0.897 |
| Intercept * Women * Non-manual | -0.21 (-0.42, 0.00) | 0.048 | 0.01 (-0.25, 0.27) | 0.964 |
| Spline 1, linear term * Women * Non-manual | 0.01 (-0.05, 0.07) | 0.689 | 0.00 (-0.12, 0.11) | 0.931 |
| Spline 1, quadratic term * Women * Non-manual | 0.000 (-0.005, 0.004) | 0.833 | 0.000 (-0.011, 0.012) | 0.961 |
| Spline 1, cubic term * Women * Non-manual | 0.0000 (-0.0001, 0.0001) | 0.933 | 0.0000 (-0.0003, 0.0003) | 0.910 |
| Spline 2, linear term * Women * Non-manual | -0.01 (-0.48, 0.46) | 0.967 | 0.04 (-0.46, 0.54) | 0.866 |
| Spline 2, quadratic term * Women * Non-manual | 0.02 (-0.16, 0.19) | 0.848 | -0.01 (-0.19, 0.17) | 0.882 |
| Spline 1, linear term (SD) | 0.03 (0.03, 0.03) | | 0.05 (0.05, 0.05) | |
| Spline 2, linear term (SD) | 0.23 (0.20, 0.27) | | 0.27 (0.23, 0.30) | |
| Intercept (SD) | 0.98 (0.94, 1.02) | | 1.24 (1.20, 1.28) | |
| Residual (SD) | 1.09 (1.06, 1.12) | | 1.19 (1.16, 1.21) | |
| **Gender (ref.: Men) and childhood housing tenure (ref.: Rented at both time-points)** | | | | |
| Intercept | 0.85 (0.74, 0.96) | <0.001 | 1.45 (1.29, 1.61) | <0.001 |
| Spline 1, linear term | -0.03 (-0.07, 0.00) | 0.042 | -0.02 (-0.09, 0.06) | 0.646 |
| Spline 1, quadratic term | 0.004 (0.001, 0.006) | 0.001 | 0.005 (-0.003, 0.012) | 0.214 |
| Spline 1, cubic term | -0.0001 (-0.0001, 0.0000) | 0.001 | -0.0002 (-0.0003, 0.0000) | 0.132 |
| Spline 2, linear term | 0.27 (0.02, 0.51) | 0.032 | 0.40 (0.05, 0.74) | 0.027 |
| Spline 2, quadratic term | -0.09 (-0.18, 0.00) | 0.052 | -0.18 (-0.31, -0.05) | 0.006 |
| Intercept * Women | 0.83 (0.68, 0.99) | <0.001 | 0.82 (0.58, 1.06) | <0.001 |
| Spline 1, linear term * Women | -0.05 (-0.09, 0.00) | 0.055 | -0.06 (-0.17, 0.04) | 0.240 |
| Spline 1, quadratic term * Women | 0.002 (-0.001, 0.006) | 0.161 | 0.003 (-0.008, 0.014) | 0.618 |
| Spline 1, cubic term * Women | 0.0000 (-0.0001, 0.0000) | 0.287 | 0.0000 (-0.0003, 0.0003) | 0.971 |
| Spline 2, linear term * Women | 0.23 (-0.15, 0.60) | 0.232 | -0.17 (-0.69, 0.35) | 0.518 |
| Spline 2, quadratic term * Women | -0.10 (-0.24, 0.03) | 0.138 | 0.08 (-0.11, 0.26) | 0.415 |
| Intercept * Owned at one time-point | -0.14 (-0.43, 0.15) | 0.332 | -0.05 (-0.38, 0.28) | 0.761 |
| Intercept * Owned at both time-points | -0.19 (-0.33, -0.04) | 0.014 | -0.23 (-0.44, -0.02) | 0.031 |
| Spline 1, linear term * Owned at one time-point | 0.01 (-0.09, 0.11) | 0.810 | -0.03 (-0.16, 0.11) | 0.716 |
| Spline 1, linear term * Owned at both time-points | 0.01 (-0.04, 0.05) | 0.730 | -0.01 (-0.11, 0.08) | 0.761 |
| Spline 1, quadratic term * Owned at one time-point | 0.000 (-0.007, 0.006) | 0.892 | 0.002 (-0.012, 0.015) | 0.806 |
| Spline 1, quadratic term * Owned at both time-points | 0.000 (-0.003, 0.003) | 0.999 | 0.001 (-0.008, 0.010) | 0.754 |

| | | | | |
|---|---|---|---|---|
| Spline 1, cubic term * Owned at one time-point | 0.0000 (-0.0001, 0.0001) | 0.945 | 0.0000 (-0.0004, 0.0003) | 0.797 |
| Spline 1, cubic term * Owned at both time-points | 0.0000 (-0.0001, 0.0001) | 0.875 | 0.0000 (-0.0003, 0.0002) | 0.743 |
| Spline 2, linear term * Owned at one time-point | -0.06 (-0.69, 0.56) | 0.842 | 0.06 (-0.64, 0.75) | 0.871 |
| Spline 2, linear term * Owned at both time-points | -0.08 (-0.40, 0.25) | 0.652 | -0.09 (-0.49, 0.31) | 0.650 |
| Spline 2, quadratic term * Owned at one time-point | 0.03 (-0.20, 0.26) | 0.823 | 0.00 (-0.26, 0.26) | 0.984 |
| Spline 2, quadratic term * Owned at both time-points | 0.02 (-0.11, 0.14) | 0.785 | 0.04 (-0.11, 0.18) | 0.615 |
| Intercept * Women * Owned at one time-point | -0.05 (-0.45, 0.35) | 0.822 | -0.18 (-0.63, 0.28) | 0.450 |
| Intercept * Women * Owned at both time-points | -0.27 (-0.49, -0.05) | 0.014 | -0.13 (-0.43, 0.18) | 0.405 |
| Spline 1, linear term * Women * Owned at one time-point | 0.00 (-0.12, 0.11) | 0.943 | 0.01 (-0.19, 0.20) | 0.960 |
| Spline 1, linear term * Women * Owned at both time-points | 0.03 (-0.04, 0.09) | 0.420 | 0.03 (-0.10, 0.16) | 0.664 |
| Spline 1, quadratic term * Women * Owned at one time-point | 0.000 (-0.008, 0.008) | 0.983 | 0.001 (-0.019, 0.021) | 0.890 |
| Spline 1, quadratic term * Women * Owned at both time-points | -0.002 (-0.006, 0.003) | 0.444 | -0.002 (-0.016, 0.011) | 0.715 |
| Spline 1, cubic term * Women * Owned at one time-point | 0.0000 (-0.0001, 0.0001) | 0.977 | 0.0000 (-0.0006, 0.0005) | 0.866 |
| Spline 1, cubic term * Women * Owned at both time-points | 0.0000 (0.0000, 0.0001) | 0.482 | 0.0000 (-0.0003, 0.0004) | 0.791 |
| Spline 2, linear term * Women * Owned at one time-point | 0.03 (-0.80, 0.86) | 0.947 | 0.17 (-0.82, 1.15) | 0.739 |
| Spline 2, linear term * Women * Owned at both time-points | -0.08 (-0.59, 0.42) | 0.750 | 0.34 (-0.26, 0.95) | 0.265 |
| Spline 2, quadratic term * Women * Owned at one time-point | 0.00 (-0.31, 0.31) | 0.989 | -0.10 (-0.46, 0.27) | 0.590 |
| Spline 2, quadratic term * Women * Owned at both time-points | 0.03 (-0.16, 0.22) | 0.735 | -0.14 (-0.35, 0.08) | 0.218 |
| Spline 1, linear term (SD) | 0.03 (0.03, 0.03) | | 0.05 (0.05, 0.05) | |
| Spline 2, linear term (SD) | 0.23 (0.20, 0.27) | | 0.27 (0.23, 0.30) | |
| Intercept (SD) | 0.98 (0.93, 1.02) | | 1.24 (1.20, 1.28) | |
| Residual (SD) | 1.09 (1.06, 1.12) | | 1.19 (1.16, 1.21) | |

*Note*. BCS/70: 1970 British Cohort Study; B: coefficient; CI: confidence interval; NCDS/58: 1958 National Child Development Study; p: significance level; SD: standard deviation (standardised random effects: intercepts, slopes, and residual). Results based on weighted and imputed data. The intercept is located at the first time-point in each of the trajectories, corresponding to age 23 in NCDS/58 and age 26 in BCS/70.

**eAppendix 9. Results from sensitivity check models including continuous age in most recent main survey data collection time-point**

| | NCDS/58 (n=8,215) | | BCS/70 (n=7,789) | |
|---|---|---|---|---|
| **Overall** | B (95% CI) | p | B (95% CI) | p |
| Intercept | 1.12 (1.07, 1.16) | <0.001 | 1.68 (1.63, 1.73) | <0.001 |
| Spline 1, linear term | -0.04 (-0.05, -0.03) | <0.001 | -0.05 (-0.07, -0.02) | <0.001 |
| Spline 1, quadratic term | 0.004 (0.003, 0.005) | <0.001 | 0.006 (0.003, 0.008) | <0.001 |
| Spline 1, cubic term | -0.0001 (-0.0001, -0.0001) | <0.001 | -0.0002 (-0.0002, -0.0001) | <0.001 |
| Spline 2, linear term | 0.18 (0.10, 0.25) | <0.001 | 0.15 (0.06, 0.24) | 0.002 |
| Spline 2, quadratic term | -0.06 (-0.08, -0.03) | <0.001 | -0.07 (-0.10, -0.04) | <0.001 |
| Spline 1, linear term (SD) | 0.03 (0.03, 0.03) | | 0.05 (0.05, 0.06) | |
| Spline 2, linear term (SD) | 0.21 (0.18, 0.25) | | 0.26 (0.23, 0.29) | |
| Intercept (SD) | 1.02 (0.99, 1.06) | | 1.28 (1.24, 1.32) | |
| Residual (SD) | 1.09 (1.07, 1.12) | | 1.19 (1.17, 1.22) | |

*Note*. BCS/70: 1970 British Cohort Study; B: coefficient; CI: confidence interval; NCDS/58: 1958 National Child Development Study; p: significance level; SD: standard deviation (standardised random effects: intercepts, slopes, and residual). Results based on weighted and imputed data. The intercept is located at the first time-point in each of the trajectories, corresponding to age 23 in NCDS/58 and age 26 in BCS/70.

**eAppendix 10. Plot of marginal predicted levels for each time-point and group (long-term trajectory plots) from sensitivity check models including continuous age in most recent main survey data collection time-point (NCDS/58, n=8,215; BCS/70, n=7,789)**

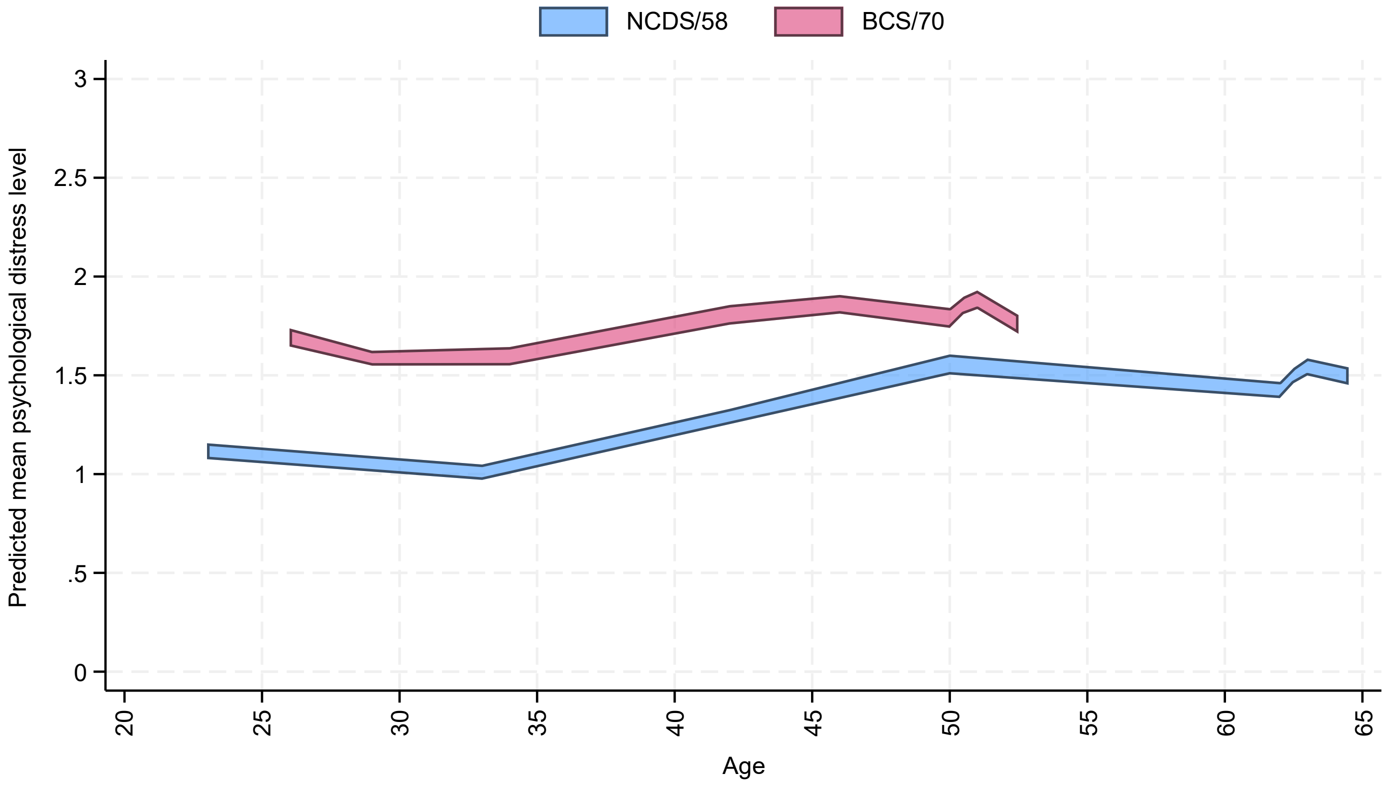

*Note.* 95% confidence intervals for the marginal predicted mean psychological distress levels from the multilevel growth curve models. Results based on weighted and imputed data.

**eAppendix 11. Estimates and 95% confidence intervals (CIs) from pooled analyses comparing psychological distress levels and gaps between most recent main survey sweeps and earliest time-point or point of highest psychological distress during COVID-19 pandemic.**

| | Pooled samples (n=14,182) | | | | | |
|---|---|---|---|---|---|---|
| | First vs last | | Pre-pandemic vs last | | Pandemic vs last | |
| | B (95% CI) | *p* | B (95% CI) | *p* | B (95% CI) | *p* |
| **Overall** | | | | | | |
| Time | 0.35 (0.26, 0.45) | <0.001 | 0.01 (-0.07, 0.09) | 0.791 | -0.16 (-0.23, -0.08) | <0.001 |
| Cohort (ref.: NCDS/58): BCS/70 | 0.61 (0.53, 0.69) | <0.001 | 0.36 (0.27, 0.44) | <0.001 | 0.45 (0.36, 0.54) | <0.001 |
| Time * BCS/70 | -0.33 (-0.45, -0.21) | <0.001 | -0.09 (-0.20, 0.01) | 0.087 | -0.18 (-0.29, -0.07) | 0.002 |
| **Gender (ref.: Men)** | | | | | | |
| Time | 0.41 (0.28, 0.54) | <0.001 | 0.00 (-0.13, 0.12) | 0.974 | -0.06 (-0.17, 0.04) | 0.248 |
| Cohort (ref.: NCDS/58): BCS/70 | 0.59 (0.48, 0.69) | <0.001 | 0.39 (0.28, 0.51) | <0.001 | 0.45 (0.33, 0.57) | <0.001 |
| Women | 0.72 (0.61, 0.83) | <0.001 | 0.58 (0.45, 0.70) | <0.001 | 0.79 (0.67, 0.91) | <0.001 |
| Time * BCS/70 | -0.41 (-0.58, -0.25) | <0.001 | -0.22 (-0.38, -0.06) | 0.006 | -0.27 (-0.42, -0.12) | 0.001 |
| Time * Women | -0.12 (-0.30, 0.06) | 0.208 | 0.03 (-0.15, 0.21) | 0.765 | -0.19 (-0.35, -0.04) | 0.016 |
| BCS/70 * Women | 0.01 (-0.14, 0.17) | 0.860 | -0.10 (-0.27, 0.07) | 0.254 | -0.04 (-0.22, 0.14) | 0.653 |
| Time * BCS/70 * Women | 0.16 (-0.09, 0.41) | 0.203 | 0.25 (0.02, 0.48) | 0.036 | 0.19 (-0.02, 0.41) | 0.082 |
| **Parental social class (ref.: Manual)** | | | | | | |
| Time | 0.38 (0.25, 0.50) | <0.001 | 0.08 (-0.04, 0.20) | 0.179 | -0.13 (-0.23, -0.02) | 0.019 |
| Cohort (ref.: NCDS/58): BCS/70 | 0.64 (0.53, 0.75) | <0.001 | 0.42 (0.31, 0.54) | <0.001 | 0.48 (0.36, 0.61) | <0.001 |
| Non-manual | -0.24 (-0.35, -0.12) | <0.001 | -0.09 (-0.23, 0.04) | 0.172 | -0.21 (-0.34, -0.08) | 0.002 |
| Time * BCS/70 | -0.36 (-0.53, -0.19) | <0.001 | -0.16 (-0.31, -0.01) | 0.038 | -0.21 (-0.37, -0.06) | 0.008 |
| Time * Non-manual | -0.07 (-0.26, 0.13) | 0.505 | -0.20 (-0.39, -0.01) | 0.039 | -0.08 (-0.24, 0.07) | 0.287 |
| BCS/70 * Non-manual | -0.04 (-0.20, 0.13) | 0.674 | -0.15 (-0.33, 0.03) | 0.112 | -0.06 (-0.24, 0.13) | 0.558 |
| Time * BCS/70 * Non-manual | 0.07 (-0.21, 0.35) | 0.608 | 0.19 (-0.05, 0.43) | 0.122 | 0.10 (-0.11, 0.31) | 0.361 |
| **Gender (ref.: Men) and parental social class (ref.: Manual)** | | | | | | |
| Time | 0.45 (0.28, 0.63) | <0.001 | 0.07 (-0.11, 0.24) | 0.452 | -0.01 (-0.15, 0.13) | 0.882 |

| | | | | | | |
|---|---|---|---|---|---|---|
| Cohort (ref.: NCDS/58): BCS/70 | 0.67 (0.52, 0.82) | <0.001 | 0.45 (0.28, 0.61) | <0.001 | 0.50 (0.32, 0.68) | <0.001 |
| Women | 0.81 (0.66, 0.95) | <0.001 | 0.62 (0.45, 0.79) | <0.001 | 0.90 (0.73, 1.06) | <0.001 |
| Non-manual | -0.11 (-0.26, 0.05) | 0.184 | -0.03 (-0.22, 0.16) | 0.756 | -0.06 (-0.24, 0.12) | 0.527 |
| Time * BCS/70 | -0.50 (-0.74, -0.27) | <0.001 | -0.29 (-0.50, -0.07) | 0.009 | -0.33 (-0.55, -0.11) | 0.004 |
| Time * Women | -0.16 (-0.41, 0.09) | 0.210 | 0.03 (-0.22, 0.27) | 0.823 | -0.24 (-0.45, -0.03) | 0.026 |
| Time * Non-manual | -0.13 (-0.41, 0.16) | 0.377 | -0.20 (-0.47, 0.08) | 0.162 | -0.15 (-0.37, 0.07) | 0.176 |
| BCS/70 * Women | -0.08 (-0.31, 0.14) | 0.472 | -0.07 (-0.31, 0.17) | 0.578 | -0.06 (-0.31, 0.19) | 0.638 |
| BCS/70 * Non-manual | -0.19 (-0.42, 0.04) | 0.104 | -0.13 (-0.37, 0.12) | 0.320 | -0.11 (-0.39, 0.16) | 0.414 |
| Women * Non-manual | -0.25 (-0.50, -0.01) | 0.043 | -0.12 (-0.39, 0.15) | 0.393 | -0.29 (-0.56, -0.02) | 0.035 |
| Time * BCS/70 * Women | 0.30 (-0.05, 0.65) | 0.096 | 0.26 (-0.05, 0.57) | 0.103 | 0.24 (-0.05, 0.53) | 0.105 |
| Time * BCS/70 * Non-manual | 0.25 (-0.13, 0.64) | 0.199 | 0.19 (-0.13, 0.52) | 0.245 | 0.17 (-0.17, 0.50) | 0.325 |
| Time * Women * Non-manual | 0.13 (-0.31, 0.56) | 0.563 | 0.00 (-0.38, 0.38) | 0.983 | 0.14 (-0.20, 0.47) | 0.417 |
| BCS/70 * Women * Non-manual | 0.28 (-0.08, 0.64) | 0.121 | -0.05 (-0.41, 0.31) | 0.785 | 0.10 (-0.30, 0.50) | 0.621 |
| Time * BCS/70 * Women * Non-manual | -0.36 (-0.94, 0.22) | 0.225 | -0.02 (-0.50, 0.45) | 0.922 | -0.14 (-0.63, 0.35) | 0.570 |
| **Childhood housing tenure (ref.: Rented at both time-points)** | | | | | | |
| Time | 0.35 (0.22, 0.49) | <0.001 | 0.05 (-0.07, 0.18) | 0.401 | -0.17 (-0.28, -0.05) | 0.007 |
| Cohort (ref.: NCDS/58): BCS/70 | 0.66 (0.51, 0.82) | <0.001 | 0.42 (0.25, 0.59) | <0.001 | 0.51 (0.33, 0.70) | <0.001 |
| Owned at both time-points | -0.35 (-0.47, -0.23) | <0.001 | -0.23 (-0.38, -0.09) | 0.001 | -0.35 (-0.50, -0.21) | <0.001 |
| Time * BCS/70 | -0.27 (-0.50, -0.05) | 0.019 | -0.05 (-0.25, 0.15) | 0.619 | -0.16 (-0.35, 0.04) | 0.120 |
| Time * Owned at both time-points | 0.00 (-0.19, 0.20) | 0.977 | -0.10 (-0.30, 0.09) | 0.294 | 0.00 (-0.17, 0.16) | 0.961 |
| BCS/70 * Owned at both time-points | 0.04 (-0.16, 0.23) | 0.693 | -0.05 (-0.26, 0.16) | 0.638 | -0.01 (-0.24, 0.22) | 0.938 |
| Time * BCS/70 * Owned at both time-points | -0.15 (-0.43, 0.13) | 0.293 | -0.03 (-0.29, 0.23) | 0.824 | -0.04 (-0.29, 0.22) | 0.786 |
| **Gender (ref.: Men) and childhood housing tenure (ref.: Rented at both time-points)** | | | | | | |
| Time | 0.36 (0.18, 0.53) | <0.001 | 0.02 (-0.18, 0.22) | 0.846 | -0.07 (-0.23, 0.08) | 0.363 |
| Cohort (ref.: NCDS/58): BCS/70 | 0.64 (0.44, 0.84) | <0.001 | 0.53 (0.30, 0.75) | <0.001 | 0.65 (0.42, 0.89) | <0.001 |
| Women | 0.83 (0.66, 0.99) | <0.001 | 0.76 (0.55, 0.96) | <0.001 | 0.99 (0.79, 1.20) | <0.001 |

| | | | | | | |
|---|---|---|---|---|---|---|
| Owned at both time-points | -0.19 (-0.34, -0.05) | 0.008 | -0.05 (-0.24, 0.13) | 0.556 | -0.16 (-0.34, 0.03) | 0.101 |
| Time * BCS/70 | -0.29 (-0.58, 0.01) | 0.058 | -0.19 (-0.47, 0.08) | 0.172 | -0.34 (-0.60, -0.08) | 0.011 |
| Time * Women | 0.00 (-0.27, 0.26) | 0.980 | 0.07 (-0.21, 0.34) | 0.627 | -0.18 (-0.43, 0.06) | 0.144 |
| Time * Owned at both time-points | 0.03 (-0.21, 0.28) | 0.781 | -0.10 (-0.36, 0.16) | 0.463 | -0.02 (-0.24, 0.20) | 0.882 |
| BCS/70 * Women | 0.04 (-0.27, 0.34) | 0.806 | -0.21 (-0.54, 0.11) | 0.199 | -0.29 (-0.63, 0.04) | 0.085 |
| BCS/70 * Owned at both time-points | -0.05 (-0.30, 0.20) | 0.686 | -0.19 (-0.47, 0.09) | 0.186 | -0.24 (-0.54, 0.06) | 0.122 |
| Women * Owned at both time-points | -0.27 (-0.50, -0.04) | 0.021 | -0.33 (-0.60, -0.06) | 0.017 | -0.35 (-0.64, -0.07) | 0.016 |
| Time * BCS/70 * Women | 0.03 (-0.42, 0.48) | 0.901 | 0.28 (-0.12, 0.67) | 0.173 | 0.35 (-0.02, 0.72) | 0.060 |
| Time * BCS/70 * Owned at both time-points | -0.16 (-0.53, 0.21) | 0.401 | 0.02 (-0.33, 0.37) | 0.912 | 0.09 (-0.24, 0.43) | 0.588 |
| Time * Women * Owned at both time-points | -0.07 (-0.44, 0.31) | 0.733 | -0.01 (-0.38, 0.36) | 0.968 | 0.01 (-0.32, 0.35) | 0.932 |
| BCS/70 * Women * Owned at both time-points | 0.15 (-0.22, 0.52) | 0.436 | 0.24 (-0.17, 0.66) | 0.253 | 0.41 (-0.02, 0.83) | 0.063 |
| Time * BCS/70 * Women * Owned at both time-points | 0.02 (-0.53, 0.57) | 0.943 | -0.09 (-0.62, 0.43) | 0.726 | -0.24 (-0.72, 0.24) | 0.329 |

*Note*. BCS/70: 1970 British Cohort Study; B: coefficient; CI: confidence interval; NCDS/58: 1958 National Child Development Study; p: significance level. Results based on weighted and imputed data. Analyses including childhood housing tenure exclude cohort members who lived in a rented or owned house at only one of the time-points; sample size in these analyses is $n_{NCDS/58}$=4,409 and $n_{BCS/70}$=5,246. Age at first time-point, most recent pre-pandemic time-point, during-pandemic time-point, and last time-point are 23, 50, 62.5, and 64.5 in NCDS/58, and 26, 46, 50.5, and 52.5 in BCS/70.

**eAppendix 12. Results from between-person post-lockdown analyses on the relationship between inflation and psychological distress.**

The analyses focused on the post-lockdown period in the overall samples showed significant differences in the incidence rate of psychological distress symptoms by inflation, both as a continuous or binary measure, in both cohorts. Differences attenuated after adjustment for seasonality. Women (particularly in BCS/70) and those in a disadvantaged (concurrent) socioeconomic position had significantly higher overall incidence rates than men and those in an advantaged (concurrent) socioeconomic position. Gender differences in BCS/70 remained significant after adjustment for seasonality, and this was also the case in most instances for the differences across self-rated financial situations, where the confounder set was more extensive. Differences by concurrent housing tenure were smaller than by self-rated financial situation and often attenuated to non-significance or close to non-significance after adjustment. We did not find evidence of overall gaps in the incidence rates at the intersection of gender and concurrent socioeconomic position. We did not find evidence suggesting a different relationship between inflation (either continuous or binary) and psychological distress by gender, socioeconomic position, or their intersection. The only exception was for the unadjusted interaction between continuous CPIH and the 'living comfortably' self-rated financial situation in BCS/70 ($IRR_{CPIHcont*comfortably\_BCS/70}$= 0.95 [0.91, 1.00], $p$ = .044), with the coefficient becoming non-significant after confounder adjustment.

Detailed results from these analyses are provided in the table below, whereas the visual depiction of the marginal predicted number of distress symptoms before and after CPIH had been ≥6% for three months are available in **eAppendix 13 (Supplementary Material)**.

| | NCDS/58 | | | | | | | |
|---|---|---|---|---|---|---|---|---|
| | Continuous CPIH | | | | Binary (high inflation) | | | |
| | Unadjusted | | Adjusted | | Unadjusted | | Adjusted | |
| | IRR (95% CI) | *p* | IRR (95% CI) | *p* | IRR (95% CI) | *p* | IRR (95% CI) | *p* |
| **Overall, n=6,553 [6,147] **** | | | | | | | | |
| Inflation | 1.03 (1.01, 1.06) | 0.006 | 1.02 (0.99, 1.06) | 0.146 | 1.23 (1.09, 1.38) | 0.001 | 1.19 (1.00, 1.42) | 0.050 |
| **Gender (ref.: Men), n=6,553 [6,147] **** | | | | | | | | |
| Inflation | 1.03 (0.99, 1.07) | 0.106 | 1.02 (0.98, 1.06) | 0.405 | 1.38 (1.13, 1.68) | 0.001 | 1.39 (1.15, 1.67) | 0.001 |
| Women | 1.40 (0.99, 1.98) | 0.058 | 1.41 (1.00, 1.99) | 0.052 | 1.19 (0.98, 1.44) | 0.081 | 1.13 (0.89, 1.44) | 0.302 |
| Inflation * Women | 1.01 (0.96, 1.06) | 0.794 | 1.01 (0.96, 1.06) | 0.750 | 1.08 (0.85, 1.37) | 0.523 | 1.09 (0.86, 1.37) | 0.481 |
| **Housing tenure (ref.: Own outright), n=6,475 [6,076] **** | | | | | | | | |
| Inflation | 1.02 (0.99, 1.04) | 0.185 | 0.99 (0.97, 1.02) | 0.534 | 1.13 (1.00, 1.29) | 0.050 | 0.98 (0.85, 1.14) | 0.822 |
| Own with mortgage | 1.16 (0.76, 1.76) | 0.494 | 1.05 (0.70, 1.56) | 0.816 | 1.20 (0.96, 1.51) | 0.112 | 1.12 (0.90, 1.38) | 0.314 |
| Other | 1.52 (0.96, 2.39) | 0.071 | 1.21 (0.79, 1.84) | 0.383 | 1.83 (1.41, 2.38) | <0.001 | 1.24 (1.00, 1.54) | 0.055 |
| Inflation * Own with mortgage | 1.00 (0.94, 1.06) | 0.961 | 1.00 (0.95, 1.06) | 0.947 | 0.95 (0.70, 1.29) | 0.757 | 0.94 (0.72, 1.22) | 0.635 |
| Inflation * Other | 1.03 (0.97, 1.09) | 0.387 | 0.99 (0.93, 1.05) | 0.712 | 1.04 (0.77, 1.41) | 0.814 | 0.88 (0.67, 1.17) | 0.384 |
| **Self-rated financial situation (ref.: Difficulties), n=6,513 [6,123] **** | | | | | | | | |
| Inflation | 1.03 (0.99, 1.08) | 0.143 | 0.98 (0.94, 1.03) | 0.491 | 1.16 (0.94, 1.44) | 0.171 | 0.91 (0.71, 1.15) | 0.414 |
| Doing all right | 0.66 (0.43, 1.01) | 0.058 | 0.83 (0.55, 1.26) | 0.388 | 0.66 (0.52, 0.84) | 0.001 | 0.84 (0.68, 1.04) | 0.113 |
| Living comfortably | 0.46 (0.31, 0.68) | <0.001 | 0.65 (0.44, 0.96) | 0.031 | 0.40 (0.33, 0.49) | <0.001 | 0.63 (0.52, 0.77) | <0.001 |
| Inflation * Doing all right | 0.99 (0.93, 1.05) | 0.656 | 1.00 (0.94, 1.06) | 0.943 | 0.89 (0.67, 1.19) | 0.435 | 0.97 (0.75, 1.27) | 0.847 |
| Inflation * Living comfortably | 0.98 (0.93, 1.03) | 0.443 | 1.00 (0.95, 1.06) | 0.937 | 0.98 (0.75, 1.28) | 0.868 | 1.06 (0.82, 1.37) | 0.649 |
| **Gender (ref.: Men) * housing tenure (ref.: Own outright), n=6,475 [6,076] **** | | | | | | | | |
| Inflation | 1.01 (0.97, 1.05) | 0.521 | 1.00 (0.96, 1.04) | 0.884 | 1.11 (0.91, 1.35) | 0.323 | 1.07 (0.86, 1.33) | 0.520 |
| Women | 1.48 (1.02, 2.15) | 0.039 | 1.48 (1.02, 2.15) | 0.040 | 1.51 (1.24, 1.84) | <0.001 | 1.51 (1.24, 1.84) | <0.001 |
| Own with mortgage | 1.58 (0.94, 2.64) | 0.084 | 1.54 (0.92, 2.58) | 0.097 | 1.35 (1.02, 1.79) | 0.036 | 1.34 (1.02, 1.77) | 0.039 |
| Other | 1.60 (0.75, 3.42) | 0.223 | 1.57 (0.76, 3.22) | 0.221 | 2.22 (1.44, 3.41) | <0.001 | 2.18 (1.46, 3.25) | <0.001 |
| Inflation * Women | 1.01 (0.96, 1.06) | 0.678 | 1.01 (0.96, 1.06) | 0.660 | 1.08 (0.84, 1.38) | 0.543 | 1.08 (0.84, 1.38) | 0.542 |
| Inflation * Own with mortgage | 0.95 (0.88, 1.03) | 0.205 | 0.96 (0.89, 1.03) | 0.230 | 0.79 (0.52, 1.19) | 0.262 | 0.80 (0.53, 1.21) | 0.288 |

| | | | | | | | | |
|---|---|---|---|---|---|---|---|---|
| Inflation * Other | 1.04 (0.94, 1.15) | 0.478 | 1.04 (0.94, 1.15) | 0.445 | 0.97 (0.59, 1.60) | 0.897 | 0.98 (0.61, 1.56) | 0.931 |
| Women * Own with mortgage | 0.64 (0.30, 1.37) | 0.253 | 0.65 (0.30, 1.38) | 0.263 | 0.86 (0.57, 1.30) | 0.481 | 0.87 (0.58, 1.31) | 0.515 |
| Women * Other | 0.96 (0.38, 2.41) | 0.934 | 1.00 (0.41, 2.44) | 0.998 | 0.75 (0.44, 1.27) | 0.288 | 0.76 (0.46, 1.26) | 0.285 |
| Inflation * Women * Own with mortgage | 1.08 (0.97, 1.20) | 0.182 | 1.07 (0.96, 1.20) | 0.191 | 1.38 (0.78, 2.42) | 0.266 | 1.35 (0.77, 2.37) | 0.293 |
| Inflation * Women * Other | 0.98 (0.87, 1.11) | 0.772 | 0.98 (0.86, 1.11) | 0.725 | 1.13 (0.60, 2.10) | 0.710 | 1.12 (0.62, 2.03) | 0.711 |
| **Gender (ref.: Men) * self-rated financial situation (ref.: Difficulties), n=6,513 [6,123] **** | | | | | | | | |
| Inflation | 1.03 (0.96, 1.11) | 0.384 | 1.02 (0.94, 1.10) | 0.635 | 1.17 (0.81, 1.68) | 0.413 | 1.11 (0.75, 1.62) | 0.604 |
| Women | 1.30 (0.69, 2.45) | 0.422 | 1.35 (0.72, 2.53) | 0.355 | 1.34 (0.95, 1.91) | 0.098 | 1.36 (0.96, 1.93) | 0.088 |
| Doing all right | 0.67 (0.33, 1.39) | 0.282 | 0.67 (0.33, 1.36) | 0.271 | 0.68 (0.45, 1.03) | 0.066 | 0.67 (0.45, 1.00) | 0.050 |
| Living comfortably | 0.45 (0.24, 0.85) | 0.014 | 0.45 (0.24, 0.84) | 0.011 | 0.37 (0.26, 0.53) | <0.001 | 0.36 (0.26, 0.52) | <0.001 |
| Inflation * Women | 1.00 (0.92, 1.10) | 0.924 | 1.00 (0.91, 1.09) | 0.990 | 1.00 (0.65, 1.55) | 0.983 | 0.99 (0.64, 1.52) | 0.968 |
| Inflation * Doing all right | 0.97 (0.88, 1.07) | 0.543 | 0.97 (0.88, 1.07) | 0.544 | 0.76 (0.46, 1.25) | 0.276 | 0.77 (0.48, 1.24) | 0.281 |
| Inflation * Living comfortably | 0.97 (0.89, 1.06) | 0.470 | 0.97 (0.89, 1.06) | 0.492 | 1.00 (0.63, 1.59) | 0.986 | 1.01 (0.65, 1.58) | 0.957 |
| Women * Doing all right | 0.93 (0.39, 2.22) | 0.873 | 0.89 (0.38, 2.09) | 0.782 | 0.92 (0.56, 1.51) | 0.749 | 0.91 (0.56, 1.47) | 0.705 |
| Women * Living comfortably | 1.10 (0.50, 2.43) | 0.814 | 1.07 (0.49, 2.34) | 0.873 | 1.16 (0.75, 1.78) | 0.503 | 1.15 (0.75, 1.77) | 0.511 |
| Inflation * Women * Doing all right | 1.03 (0.92, 1.17) | 0.587 | 1.04 (0.92, 1.17) | 0.504 | 1.37 (0.76, 2.48) | 0.292 | 1.40 (0.79, 2.50) | 0.253 |
| Inflation * Women * Living comfortably | 1.01 (0.90, 1.13) | 0.909 | 1.01 (0.91, 1.13) | 0.827 | 0.95 (0.55, 1.65) | 0.849 | 0.96 (0.56, 1.65) | 0.880 |

| | BCS/70 | | | | | | | |
|---|---|---|---|---|---|---|---|---|
| | Continuous CPIH | | | | Binary (high inflation) | | | |
| | Unadjusted | | Adjusted | | Unadjusted | | Adjusted | |
| | IRR (95% CI) | *p* | IRR (95% CI) | *p* | IRR (95% CI) | *p* | IRR (95% CI) | *p* |
| **Overall, n=7,629 [7,561] **** | | | | | | | | |
| Inflation | 1.05 (1.03, 1.07) | <0.001 | 1.04 (1.01, 1.06) | 0.001 | 1.24 (1.13, 1.36) | <0.001 | 1.17 (1.04, 1.32) | 0.009 |
| **Gender (ref.: Men), n=7,629 [7,561] **** | | | | | | | | |
| Inflation | 1.04 (1.01, 1.08) | 0.015 | 1.04 (1.00, 1.07) | 0.065 | 1.24 (1.04, 1.49) | 0.019 | 1.19 (0.98, 1.45) | 0.085 |
| Women | 1.49 (1.13, 1.96) | 0.005 | 1.50 (1.14, 1.97) | 0.003 | 1.56 (1.31, 1.85) | <0.001 | 1.57 (1.34, 1.84) | <0.001 |
| Inflation * Women | 1.01 (0.97, 1.05) | 0.752 | 1.01 (0.97, 1.05) | 0.777 | 1.00 (0.81, 1.24) | 0.970 | 1.00 (0.82, 1.22) | 0.965 |
| **Housing tenure (ref.: Own outright), n=7,589 [7,521] **** | | | | | | | | |
| Inflation | 1.03 (0.99, 1.07) | 0.124 | 1.00 (0.97, 1.04) | 0.873 | 1.14 (0.98, 1.32) | 0.101 | 0.95 (0.82, 1.11) | 0.533 |
| Own with mortgage | 1.03 (0.79, 1.33) | 0.853 | 1.04 (0.82, 1.32) | 0.718 | 1.00 (0.88, 1.13) | 0.989 | 1.00 (0.89, 1.12) | 0.972 |
| Other | 1.50 (1.05, 2.14) | 0.025 | 1.13 (0.82, 1.57) | 0.445 | 1.57 (1.26, 1.96) | <0.001 | 1.01 (0.84, 1.21) | 0.938 |
| Inflation * Own with mortgage | 0.99 (0.95, 1.04) | 0.758 | 1.00 (0.96, 1.03) | 0.831 | 0.97 (0.81, 1.18) | 0.786 | 1.02 (0.87, 1.21) | 0.776 |
| Inflation * Other | 1.01 (0.96, 1.07) | 0.664 | 0.99 (0.94, 1.04) | 0.631 | 1.05 (0.80, 1.38) | 0.727 | 1.04 (0.82, 1.30) | 0.755 |
| **Self-rated financial situation (ref.: Difficulties), n=7,587 [7,524] **** | | | | | | | | |
| Inflation | 1.04 (1.00, 1.08) | 0.032 | 0.99 (0.96, 1.03) | 0.770 | 1.17 (0.97, 1.41) | 0.094 | 0.97 (0.80, 1.18) | 0.788 |
| Doing all right | 0.64 (0.47, 0.88) | 0.006 | 0.71 (0.52, 0.96) | 0.029 | 0.58 (0.48, 0.71) | <0.001 | 0.73 (0.62, 0.86) | <0.001 |
| Living comfortably | 0.51 (0.37, 0.70) | <0.001 | 0.65 (0.48, 0.89) | 0.008 | 0.42 (0.34, 0.50) | <0.001 | 0.63 (0.53, 0.75) | <0.001 |
| Inflation * Doing all right | 0.97 (0.93, 1.02) | 0.225 | 1.00 (0.96, 1.05) | 0.968 | 0.88 (0.70, 1.10) | 0.263 | 0.98 (0.80, 1.20) | 0.848 |
| Inflation * Living comfortably | 0.95 (0.91, 1.00) | 0.044 | 0.98 (0.94, 1.03) | 0.496 | 0.83 (0.66, 1.05) | 0.112 | 0.91 (0.73, 1.12) | 0.356 |
| **Gender (ref.: Men) * housing tenure (ref.: Own outright), n=7,589 [7,521] **** | | | | | | | | |
| Inflation | 1.02 (0.95, 1.09) | 0.621 | 1.01 (0.94, 1.08) | 0.818 | 1.23 (0.91, 1.64) | 0.172 | 1.15 (0.86, 1.54) | 0.341 |
| Women | 1.52 (0.93, 2.48) | 0.092 | 1.48 (0.92, 2.40) | 0.108 | 1.80 (1.45, 2.24) | <0.001 | 1.79 (1.44, 2.23) | <0.001 |
| Own with mortgage | 0.96 (0.58, 1.57) | 0.858 | 0.94 (0.57, 1.53) | 0.800 | 1.09 (0.87, 1.37) | 0.453 | 1.08 (0.86, 1.35) | 0.516 |
| Other | 1.74 (0.89, 3.38) | 0.104 | 1.66 (0.87, 3.20) | 0.127 | 1.94 (1.26, 3.00) | 0.003 | 1.89 (1.26, 2.82) | 0.002 |
| Inflation * Women | 1.01 (0.94, 1.10) | 0.720 | 1.02 (0.94, 1.10) | 0.610 | 0.90 (0.64, 1.26) | 0.541 | 0.92 (0.66, 1.28) | 0.616 |
| Inflation * Own with mortgage | 1.01 (0.94, 1.10) | 0.741 | 1.02 (0.94, 1.10) | 0.662 | 0.95 (0.67, 1.34) | 0.760 | 0.97 (0.69, 1.36) | 0.860 |

| | | | | | | | | |
|---|---|---|---|---|---|---|---|---|
| Inflation * Other | 1.00 (0.91, 1.10) | 0.956 | 1.01 (0.92, 1.11) | 0.850 | 0.87 (0.52, 1.45) | 0.581 | 0.90 (0.55, 1.46) | 0.664 |
| Women * Own with mortgage | 1.11 (0.62, 1.98) | 0.723 | 1.14 (0.65, 2.02) | 0.649 | 0.90 (0.69, 1.18) | 0.448 | 0.91 (0.69, 1.19) | 0.475 |
| Women * Other | 0.74 (0.34, 1.61) | 0.449 | 0.77 (0.36, 1.66) | 0.510 | 0.72 (0.44, 1.18) | 0.189 | 0.74 (0.46, 1.17) | 0.199 |
| Inflation * Women * Own with mortgage | 0.97 (0.89, 1.07) | 0.568 | 0.97 (0.88, 1.06) | 0.476 | 1.04 (0.69, 1.55) | 0.868 | 1.01 (0.68, 1.51) | 0.955 |
| Inflation * Women * Other | 1.03 (0.92, 1.15) | 0.611 | 1.02 (0.91, 1.14) | 0.711 | 1.42 (0.79, 2.57) | 0.246 | 1.36 (0.77, 2.39) | 0.292 |
| **Gender (ref.: Men) * self-rated financial situation (ref.: Difficulties), n=7,587 [7,524] **** | | | | | | | | |
| Inflation | 1.02 (0.95, 1.09) | 0.631 | 1.01 (0.95, 1.08) | 0.706 | 1.03 (0.70, 1.50) | 0.897 | 0.99 (0.68, 1.43) | 0.947 |
| Women | 1.02 (0.58, 1.79) | 0.951 | 1.03 (0.59, 1.79) | 0.910 | 1.14 (0.78, 1.67) | 0.487 | 1.16 (0.81, 1.66) | 0.405 |
| Doing all right | 0.46 (0.25, 0.83) | 0.010 | 0.47 (0.26, 0.84) | 0.011 | 0.46 (0.31, 0.68) | <0.001 | 0.47 (0.33, 0.67) | <0.001 |
| Living comfortably | 0.44 (0.24, 0.80) | 0.007 | 0.45 (0.25, 0.80) | 0.007 | 0.34 (0.23, 0.50) | <0.001 | 0.34 (0.24, 0.49) | <0.001 |
| Inflation * Women | 1.04 (0.97, 1.12) | 0.297 | 1.04 (0.97, 1.12) | 0.314 | 1.26 (0.84, 1.90) | 0.269 | 1.24 (0.84, 1.82) | 0.284 |
| Inflation * Doing all right | 1.01 (0.93, 1.09) | 0.842 | 1.01 (0.93, 1.09) | 0.873 | 1.09 (0.71, 1.68) | 0.691 | 1.08 (0.72, 1.62) | 0.713 |
| Inflation * Living comfortably | 0.94 (0.87, 1.02) | 0.147 | 0.94 (0.87, 1.02) | 0.138 | 0.83 (0.54, 1.30) | 0.417 | 0.82 (0.54, 1.26) | 0.369 |
| Women * Doing all right | 1.77 (0.89, 3.51) | 0.103 | 1.75 (0.89, 3.42) | 0.104 | 1.50 (0.98, 2.29) | 0.064 | 1.48 (0.99, 2.21) | 0.058 |
| Women * Living comfortably | 1.27 (0.65, 2.48) | 0.484 | 1.25 (0.65, 2.42) | 0.504 | 1.44 (0.94, 2.21) | 0.091 | 1.42 (0.95, 2.13) | 0.088 |
| Inflation * Women * Doing all right | 0.94 (0.86, 1.04) | 0.233 | 0.95 (0.86, 1.04) | 0.243 | 0.70 (0.43, 1.15) | 0.159 | 0.72 (0.45, 1.14) | 0.164 |
| Inflation * Women * Living comfortably | 1.02 (0.93, 1.12) | 0.702 | 1.02 (0.93, 1.12) | 0.669 | 0.98 (0.60, 1.61) | 0.936 | 1.00 (0.62, 1.62) | 0.997 |

*Note*. BCS/70: 1970 British Cohort Study; CI: confidence interval; CPIH: Consumer Prices Index including owner occupiers' housing costs; IRR: incidence rate ratio; NCDS/58: 1958 National Child Development Study; p: significance level. Results based on weighted and imputed data. ** Binary analyses exclude participants whose interview took place in 2024 (406 participants in NCDS/58; 64 participants in BCS/70); sample size for these analyses is included in brackets.

**eAppendix 13. Visual depiction of marginal mean predicted number of psychological distress symptoms before and after high inflation (Consumer Prices Index including owner occupiers' housing costs, CPIH ≥ 6%), adjusted results**

The graphs below show the marginal predicted number of symptoms before and after the Consumer Prices Index including owner occupiers' housing costs (CPIH) had been equal or higher than 6% for at least three months. Results are based weighted and imputed data and exclude participants whose interview took place in 2024 (406 participants in NCDS/58; 64 participants in BCS/70).

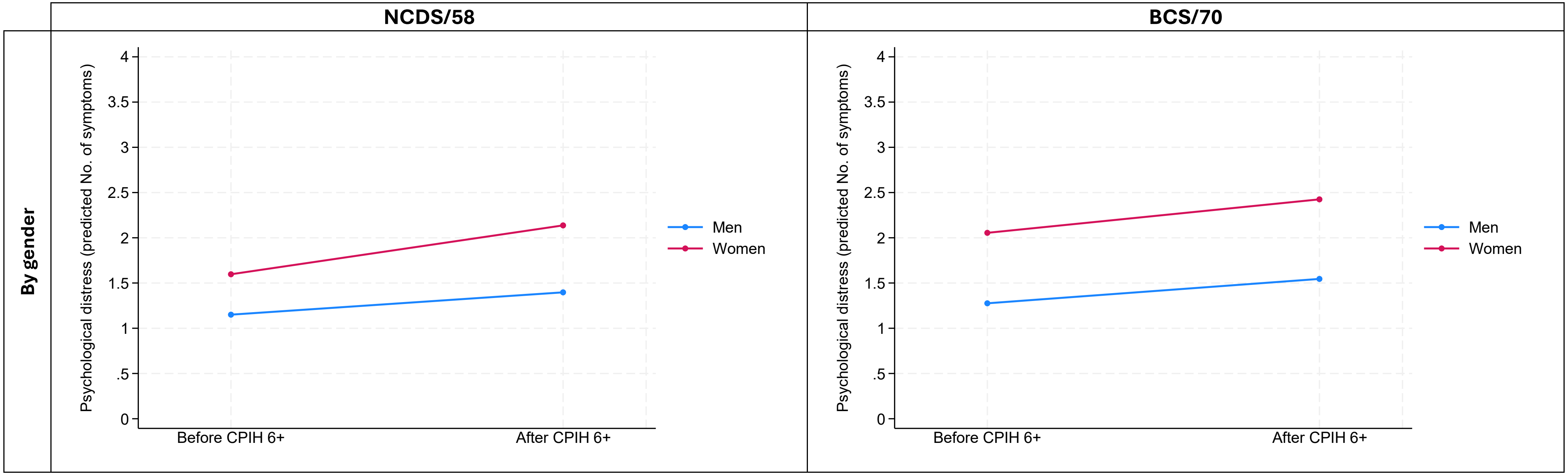

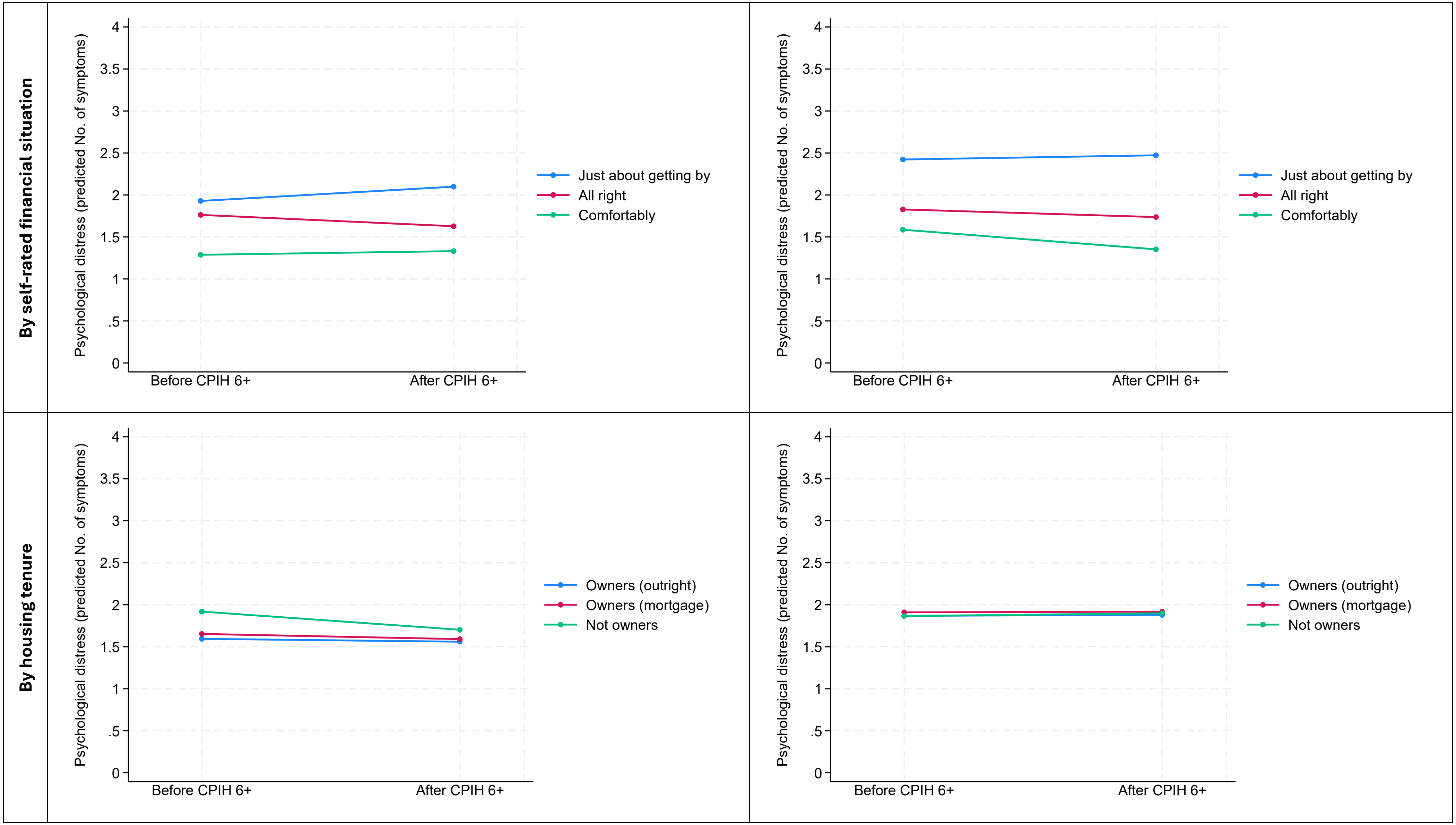
By self-rated financial situation
By housing tenure
Psychological distress (predicted No. of symptoms)
0
.5
1
1.5
2
2.5
3
3.5
4
Before CPIH 6+
After CPIH 6+
Just about getting by
All right
Comfortably
Owners (outright)
Owners (mortgage)
Not owners

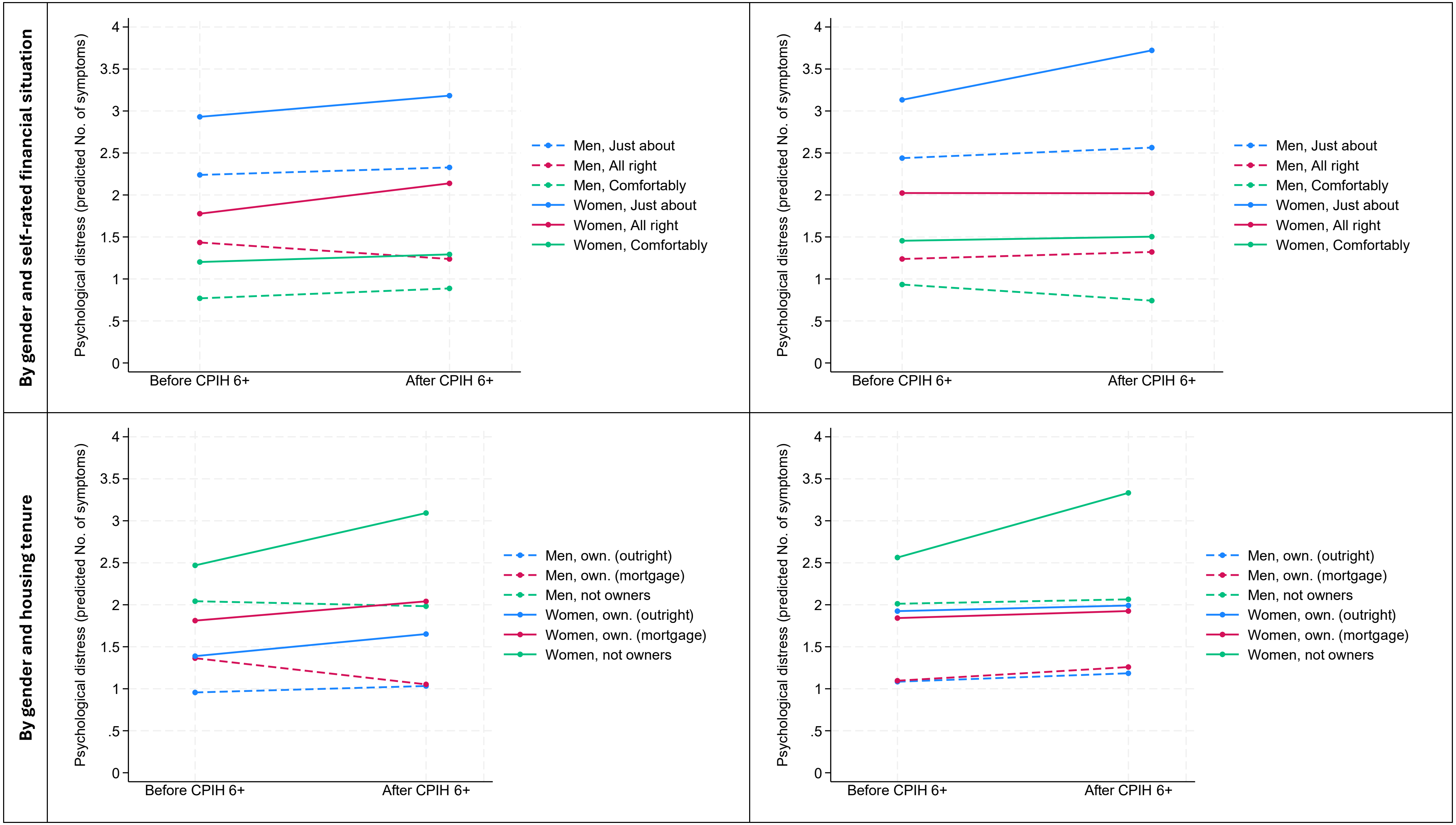
By gender and self-rated financial situation
By gender and housing tenure
Psychological distress (predicted No. of symptoms)
0
.5
1
1.5
2
2.5
3
3.5
4
Before CPIH 6+
After CPIH 6+
Men, Just about
Men, All right
Men, Comfortably
Women, Just about
Women, All right
Women, Comfortably
Men, own. (outright)
Men, own. (mortgage)
Men, not owners
Women, own. (outright)
Women, own. (mortgage)
Women, not owners
